# What is the forecasted prevalence and incidence of long-term conditions in Wales: a rapid evidence map

**DOI:** 10.1101/2023.06.23.23291814

**Authors:** Deborah Edwards, Judit Csontos, Elizabeth Gillen, Grace Hutchinson, Abubakar Sha’aban, Judith Carrier, Ruth Lewis, Rhiannon Tudor Edwards, Jacob Davies, Brendan Collins, Alison Cooper, Adrian Edwards

## Abstract

It is becoming apparent that the NHS will face many issues in years to come due to the growth of ageing population in relation to the working age population alongside the increase in multimorbidity and persistent health inequalities, particularly for preventable illness. This has implications to how healthcare and health systems are delivered, and how the NHS will need to adapt to meet the increasing demand that this places on healthcare services.

This rapid evidence map reports forecasted prevalence and incidence data across a range of long -term conditions in Wales to support planning about how best to organise and finance care for the increasing population with long-term conditions over the next 10 years. The findings by conditions include: atrial fibrillation, cancer, cardiovascular diseases, peripheral vascular disease, stroke, dementia, diabetes, heart failure, hypertension, mental illness, and multi-morbidities. Three risk factors for long term conditions were also included, i.e., poor diet/nutrition, obesity, and smoking. The review included evidence from 2012 to March 2023

**Implications for policy and practice:** Results show which long-term conditions are projected to increase over 10 years or more. Further preventive interventions through behavioural science approaches, with increased investment, should be considered to mitigate the rising prevalence of several preventable conditions. Smoking, excessive drinking and obesity are candidates for targeted preventive work, especially in areas of deprivation, to lessen health inequalities. Further research is needed for some conditions, and to provide a more comprehensive understanding of the burden of these conditions in Wales. Earlier diagnosis by genetic and genomic technologies and enabling lifestyle changes or by more cost-effective home care could reduce NHS costs of some long-term conditions.

## 1. BACKGROUND

### 1.1 Who is this Rapid Evidence Map for?

This Rapid Evidence Map was conducted as part of the Health and Care Research Wales Evidence Centre Work Programme. The above question was requested by the Science and Evidence Advice (SEA) Division, Welsh Government.

### 1.2 Purpose of this Rapid Evidence Map

It is becoming apparent that the NHS will face many issues in years to come due to the growth of ageing population in relation to the working age population alongside the increase in multimorbidity and persistent health inequalities, particularly for preventable illness (McKee et al. 2021). The growing number of the older population will result in an increase in age related conditions and a subsequent inflation in older people with care needs, estimated to be around 25% (Guzman-Castillo et al. 2017). This has implications to how healthcare and health systems are delivered, and the NHS will need to adapt to meet the increasing demand that this places on healthcare services.

The Global Burden of Disease (GBD) project sought to produce comparable estimates of ill health and injury on a global level. Data from first important paper applying GBD methodology for England highlighted that life expectancy had improved but that morbidity, especially in multiple conditions had increased (Steel et al. 2018). Public Health England have observed this pattern across other countries and attributes mostly to a slowing in the rate of improvement in cardiovascular mortality and to some extent cancer (Public Health England 2022).

In Wales, cancer and cardiovascular disease contribute to 61% of years of life lost and 36% of disability-adjusted life years (Public Health Wales Observatory 2017). The GBD report from 2016 for Wales illustrates cancer as the greatest cause of disease burden over and above cardiovascular disease (Public Health Wales Observatory 2018). The biggest risk factor contributing to current burden of disease being past smoking (Public Health Wales Observatory 2018). In England, ischaemic heart disease, lung cancer, stroke and COPD are the most common cause of disease burden with rates seen to have stabilised over recent years. However, deaths from dementia, pancreatic and colon cancer constitute a higher proportion of all deaths as absolute rates have increased (Public Health England 2022).

As the population in Wales live longer, chronic conditions such as heart disease are predicted to rise, and the likelihood of additional illnesses will also increase with age (Public Health Wales Observatory 2017). Research in the UK estimates that around 23% of the population meet current criteria for multimorbidity, increasing with age, and attention to early diagnosis, so that the figure is around two-thirds in those over 65, with nearly half having three or more conditions (Barnett et al. 2012). By 2035, the prevalence of multi-morbidity is estimated to increase (Kingston et al. 2018). In Wales as reported in 2017, 60% of those over 75 years have two or more illnesses and this is projected to significantly increase by 2035. The greatest increases across all adults over 18 years are likely to be in stroke, heart conditions and neurological conditions including dementia (Public Health Wales Observatory 2017).

The projected rise in the number of older people has raised concern about how best to plan for and finance care for this increasing population with chronic conditions. Such planning requires accurate projections for future numbers of people with chronic conditions (including multi morbidity) and future costs of providing quality of care for them. This rapid evidence map will report forecasted prevalence and incidence data across a range of chronic and long -term conditions in Wales.

## 2. FINDINGS

### 2.1 Overview of the available evidence

This section is organised structured by according to the included long-term conditions considered by the review, which included atrial fibrillation (AF), cancer (breast, colorectal and lung, prostate), cardiovascular diseases (CVD) (coronary heart disease (CHD), peripheral vascular disease (PVD); stroke and transient ischaemic attacks (TIA)), dementia, diabetes, heart failure (HF), hypertension, mental illness (anxiety, bipolar disorder, depression, psychosis, schizophrenia) and multi-morbidities. Three risk factors for long term conditions were also included, namely poor diet/nutrition, obesity and smoking.

The included studies and their projection data are summarised in Table 1 and presented according to the underlying condition and the country within which the studies were conducted. An evidence map, which outlines the research gaps is presented in Figure 1.

**Figure 1:**
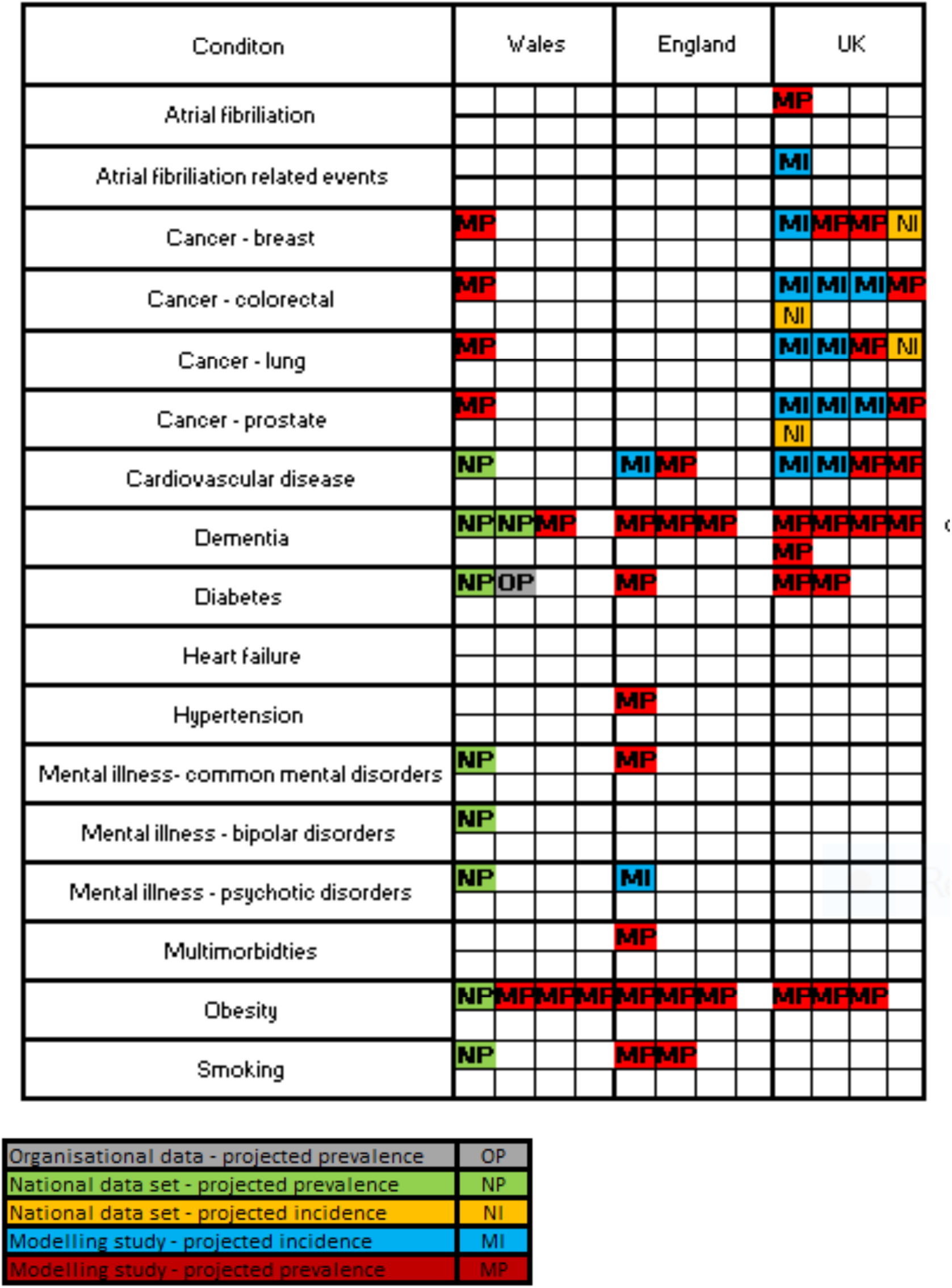
Evidence gap map: Different types of projection data (colour) by condition (rows) against country (Wales, England and UK) (columns)

**Table 1:**
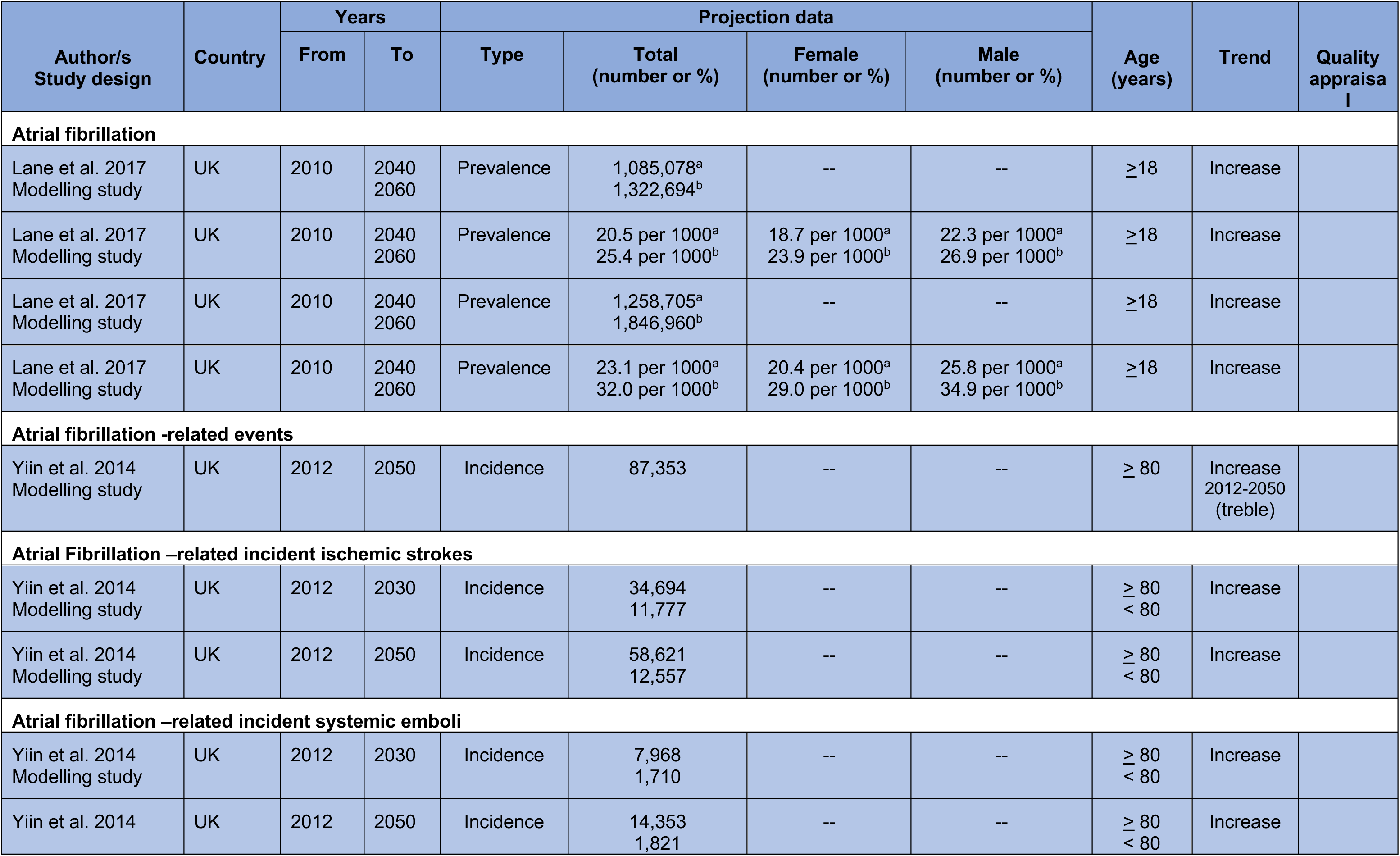
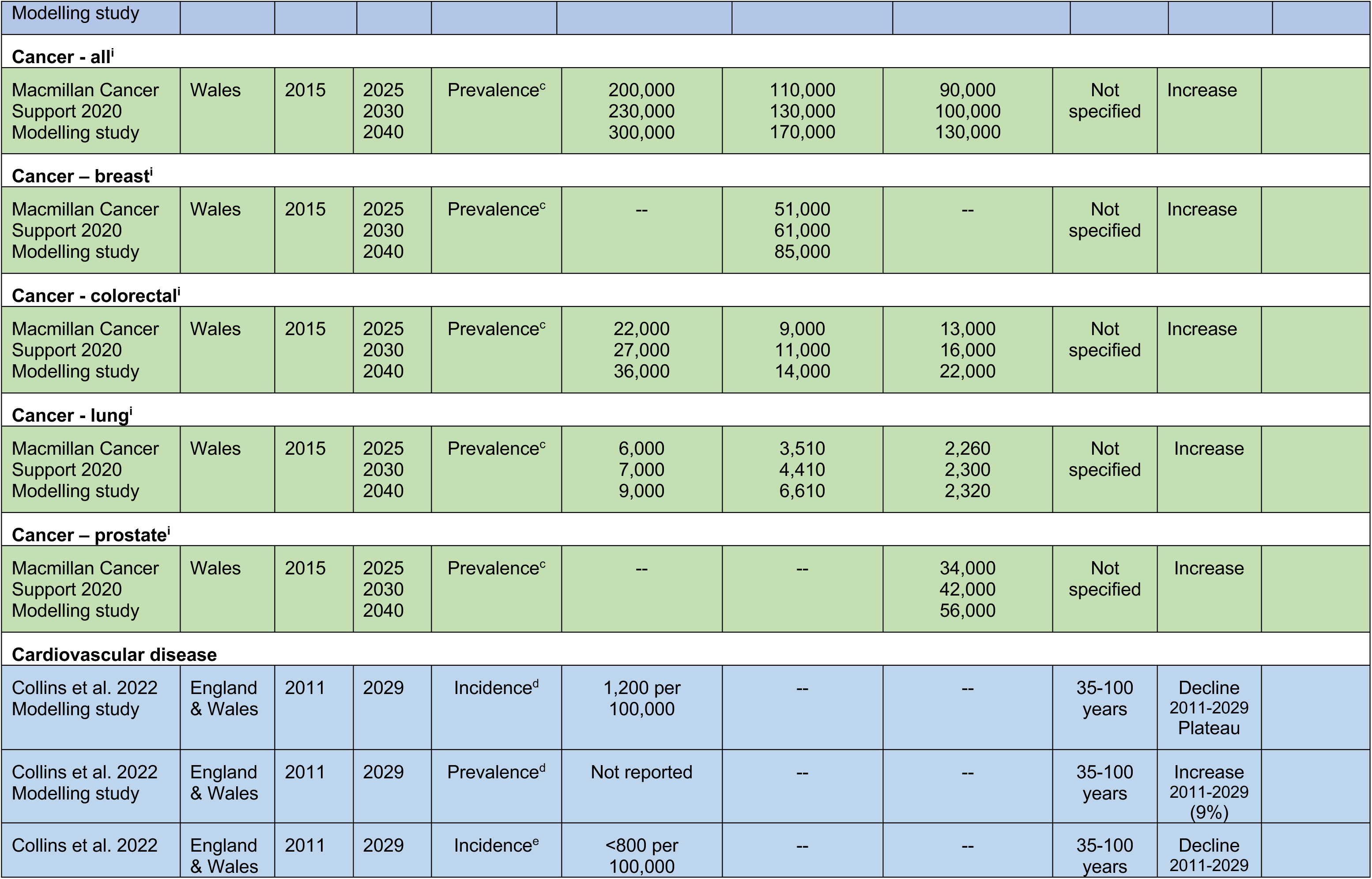
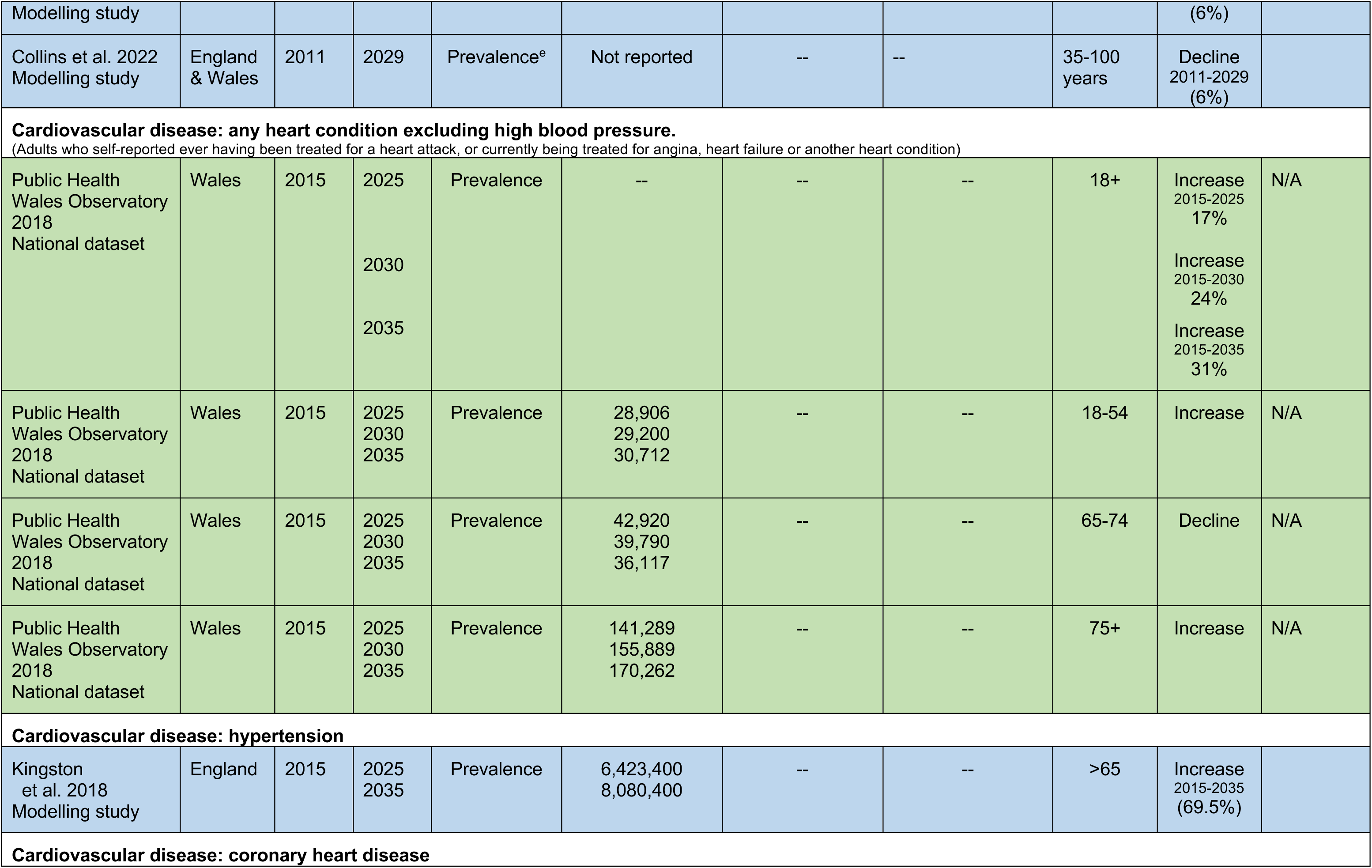
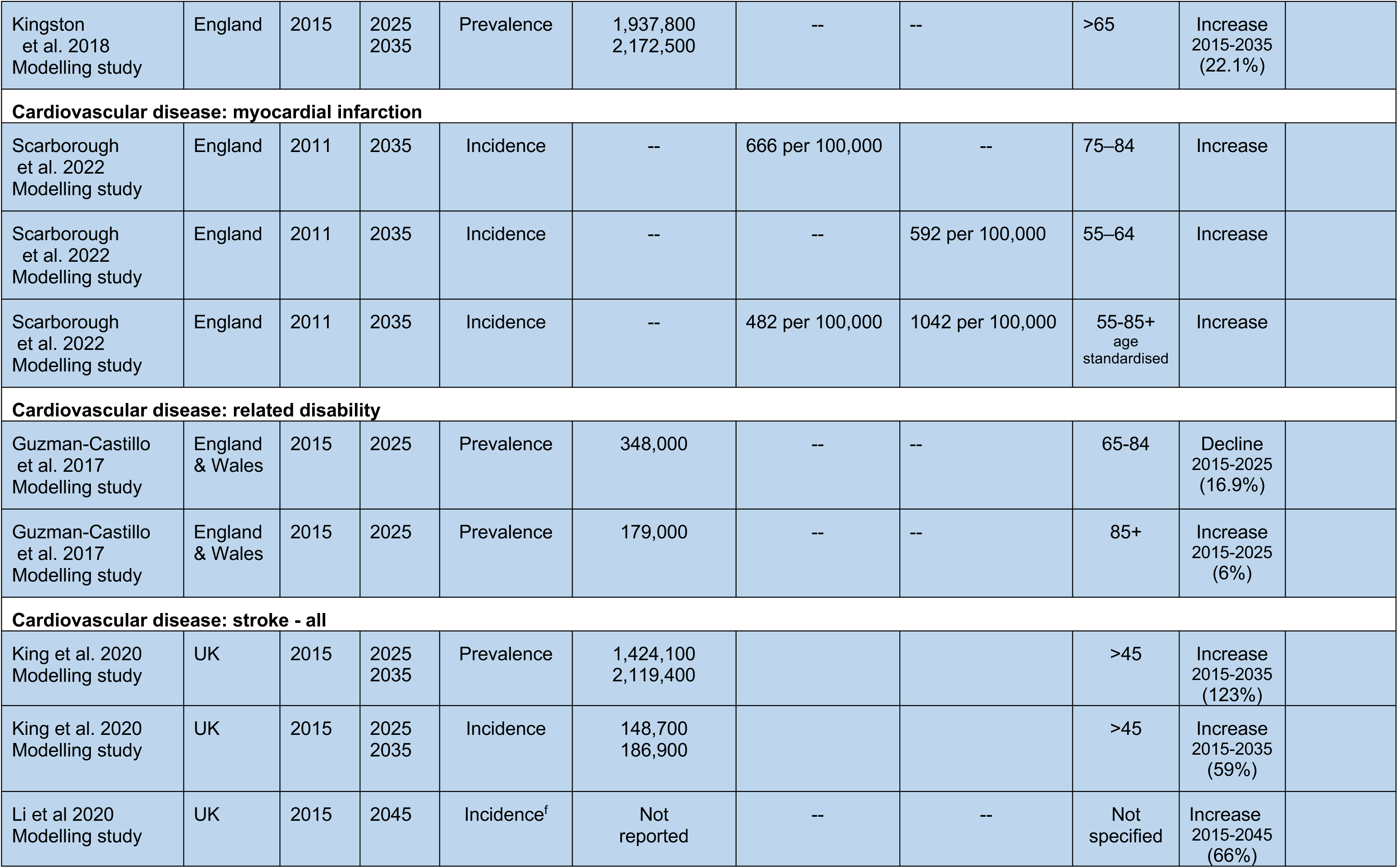
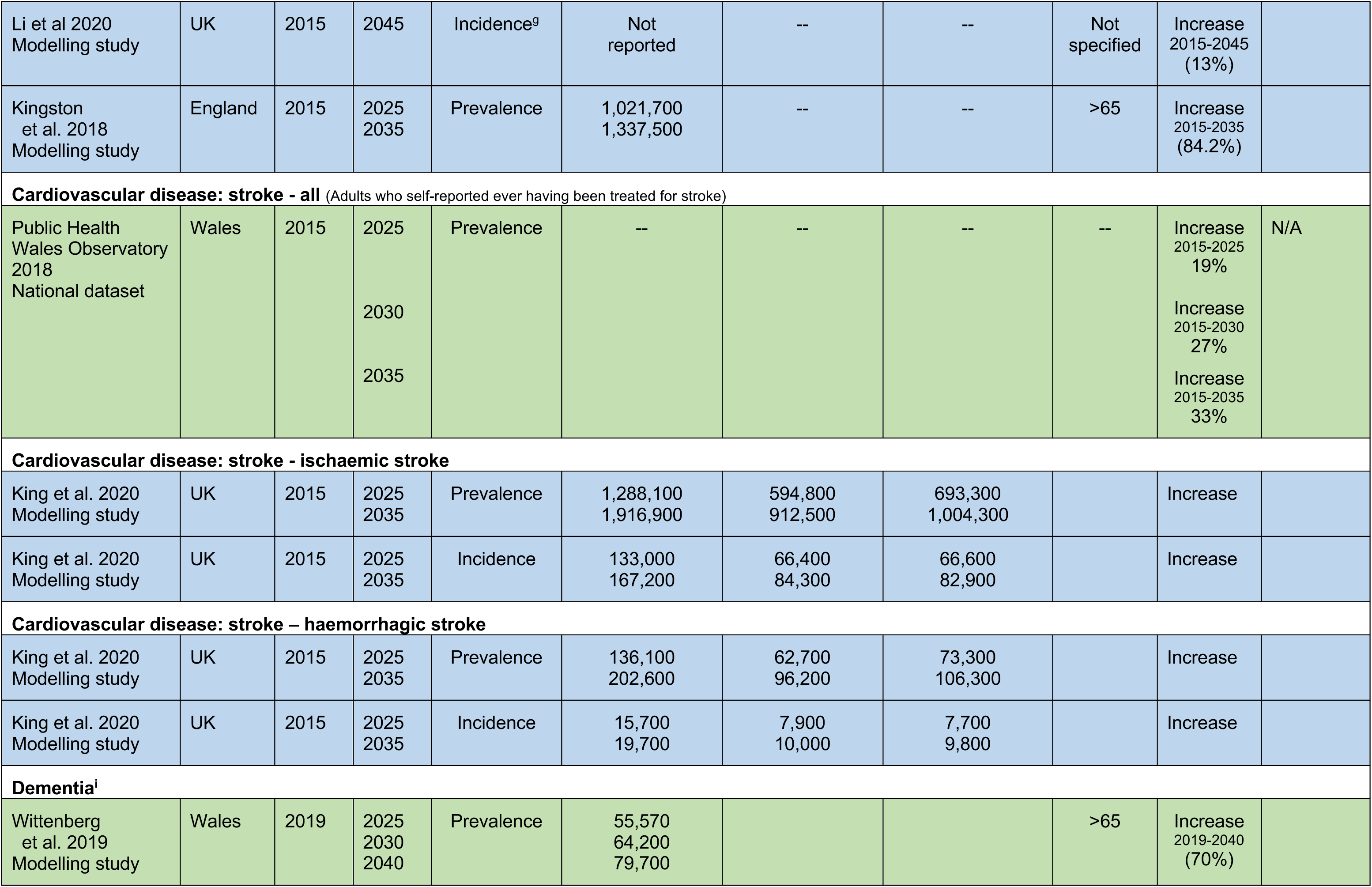
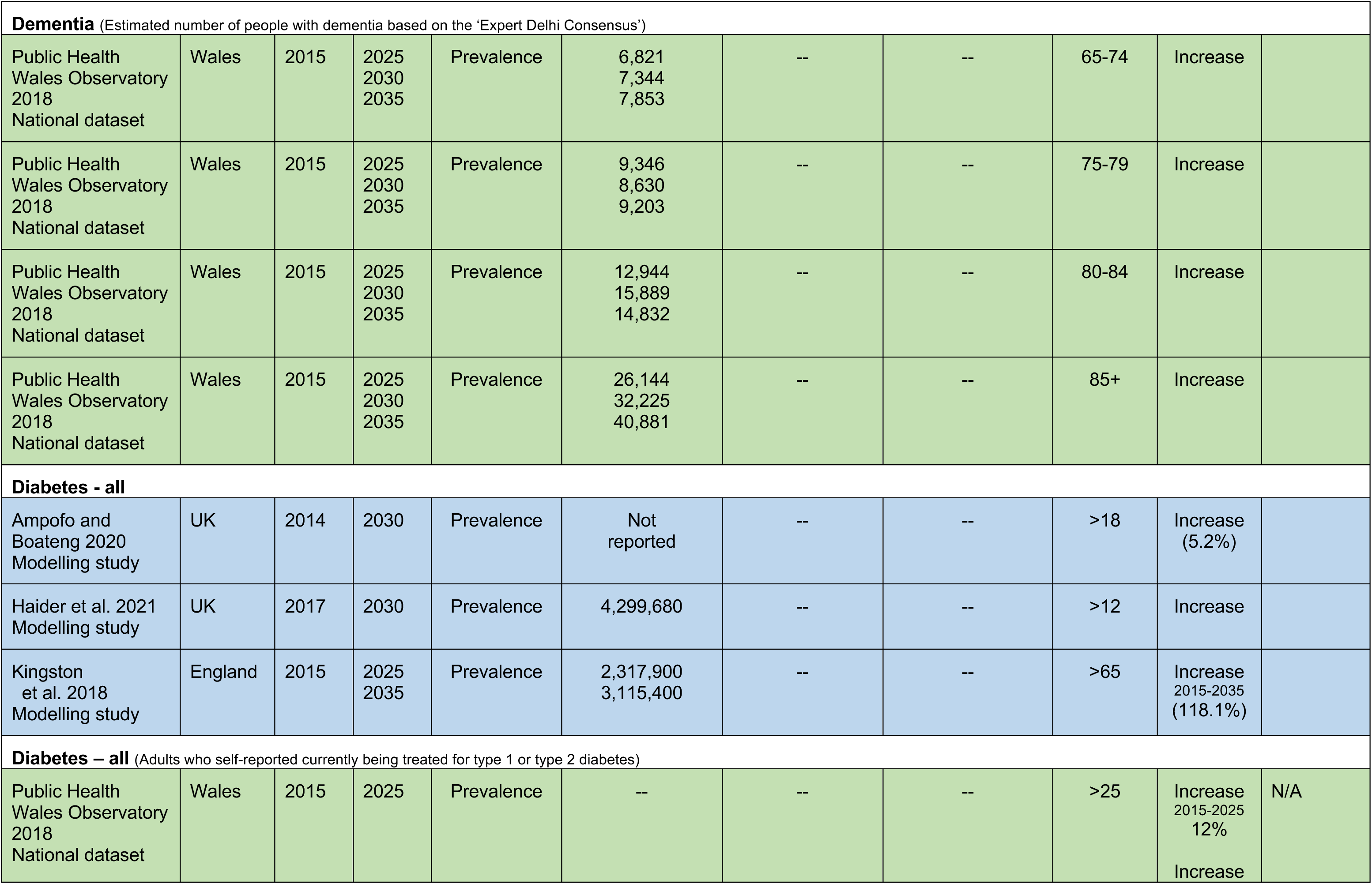
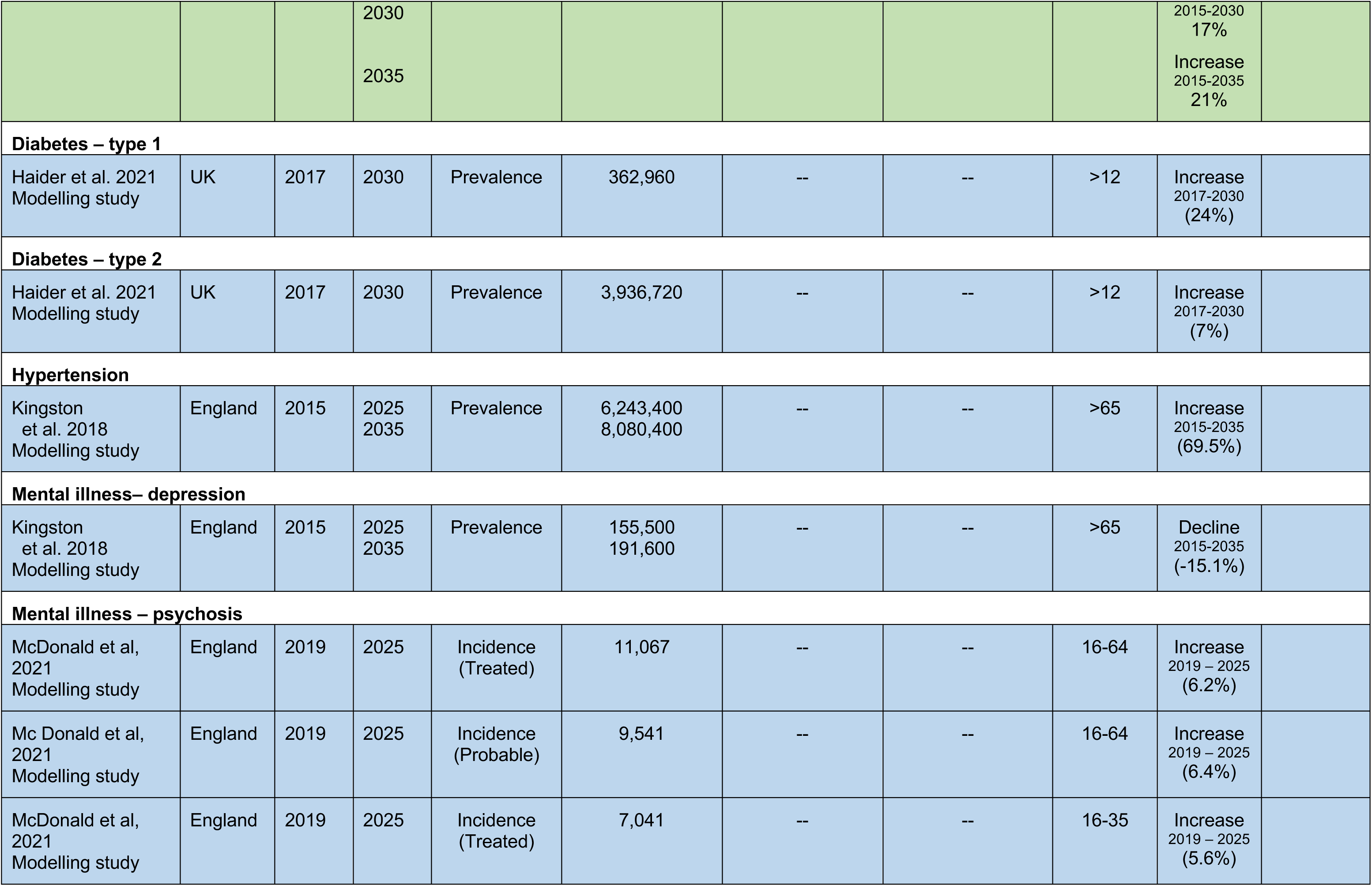
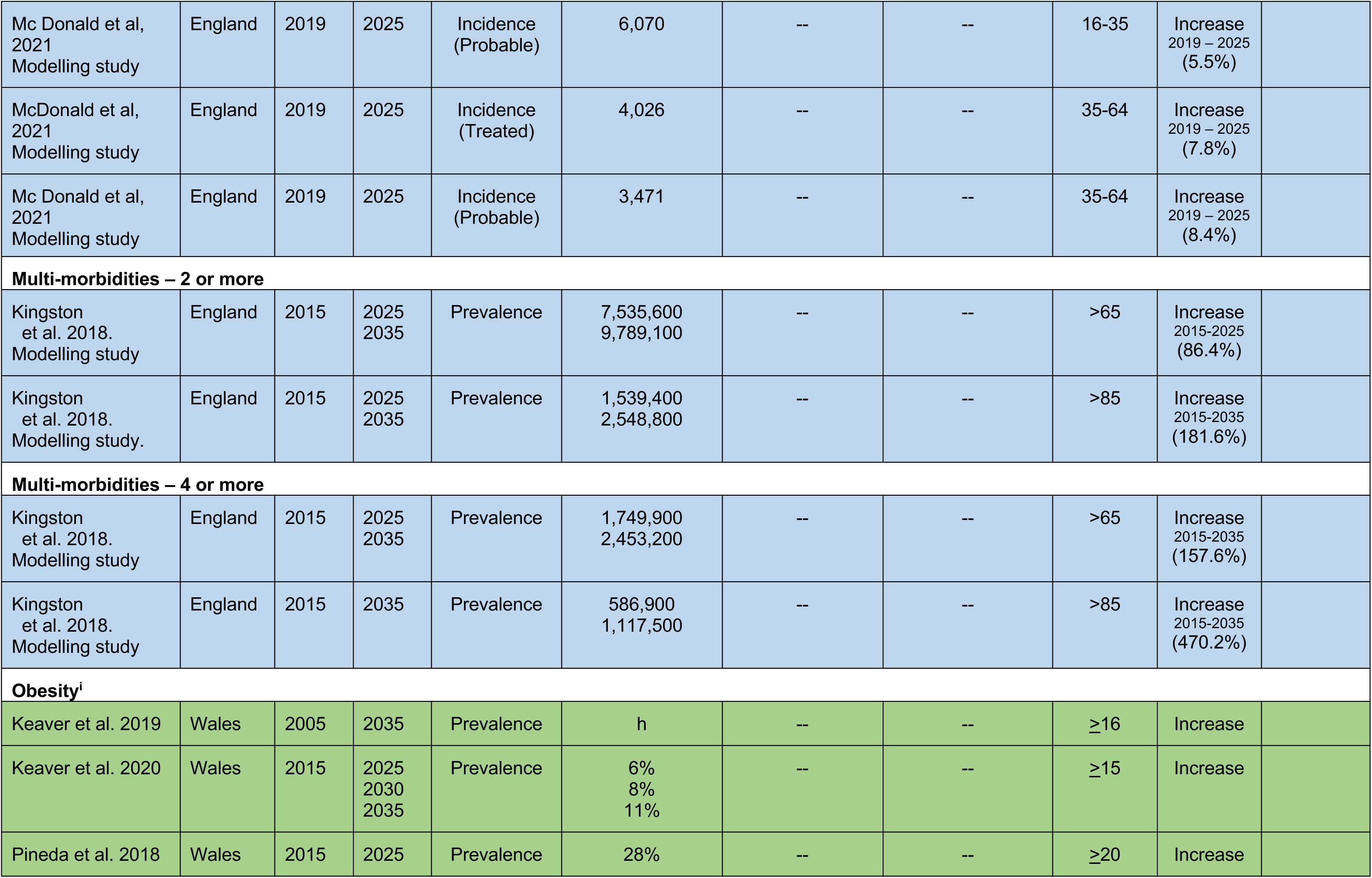
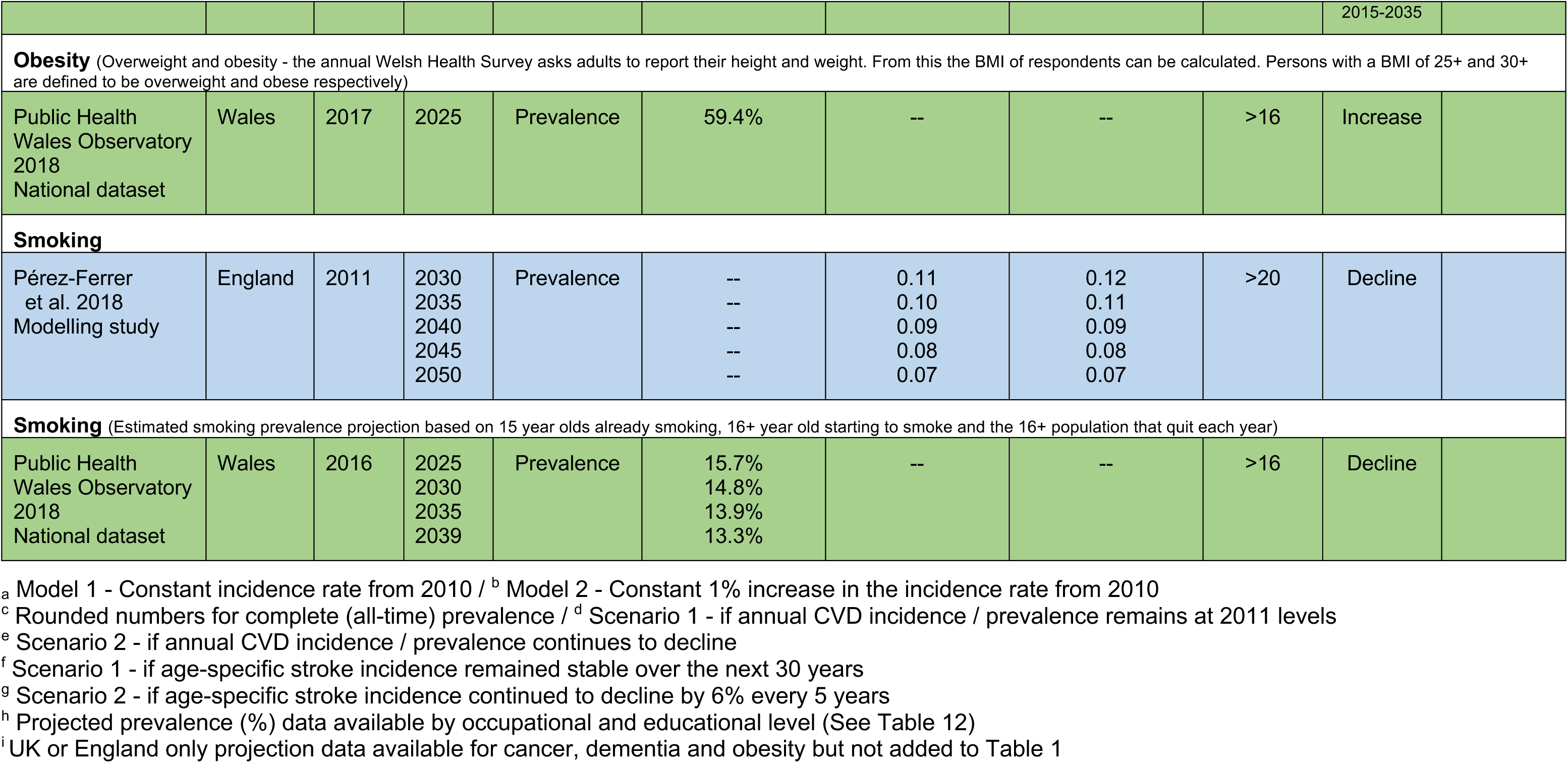
Summary table of studies by condition and year of projection data by country.

Studies were included if they reported on Welsh data, or, if no data is available from Wales, UK based data. Further details of the specific eligibility criteria used for selecting studies (Table 2), and methods used for conducting the evidence map are presented in Section 5. The results of the study selection process (Tables 3-4) and the detailed summary of the extracted data for each included studies are provided in Section 6.

**Table 2:**
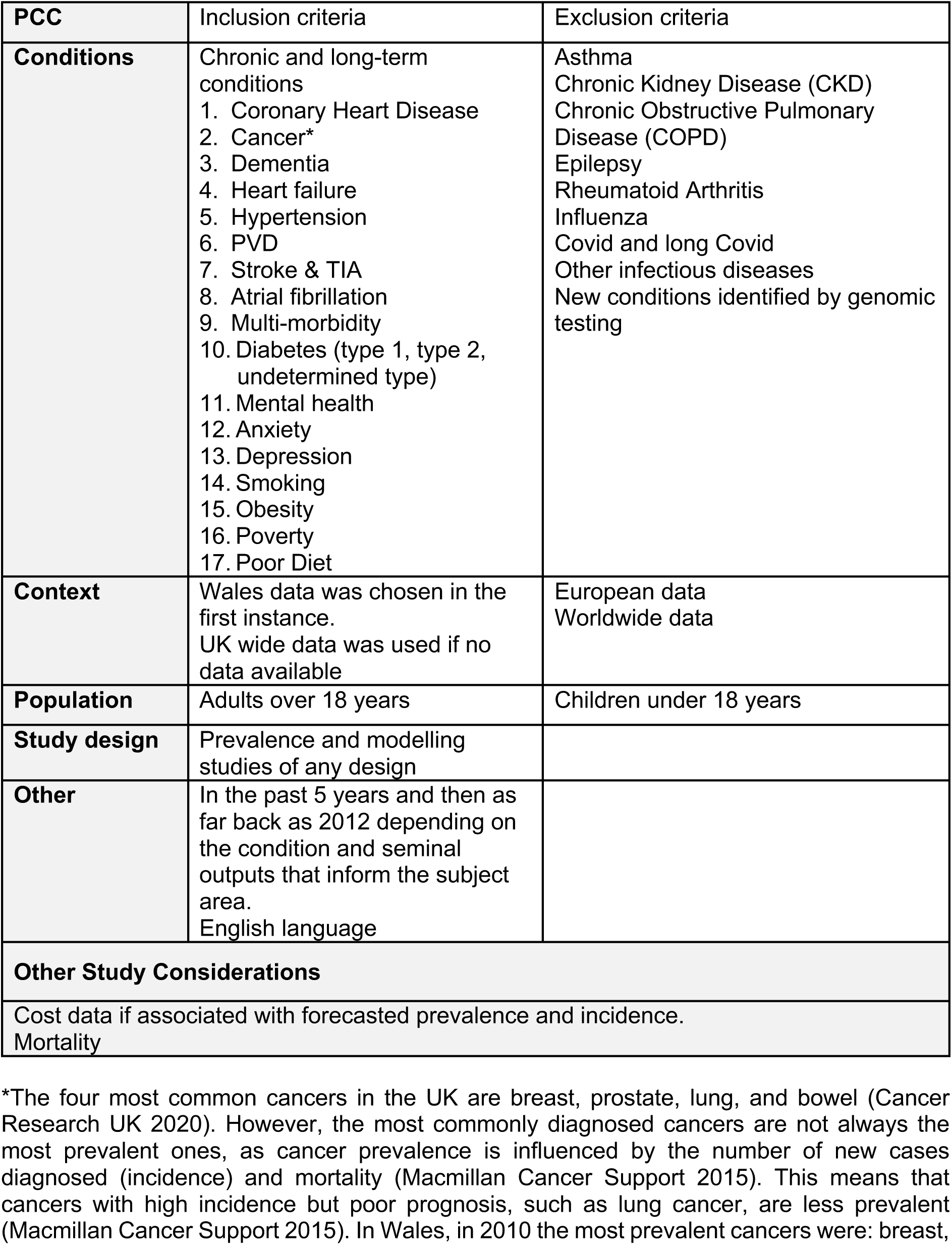
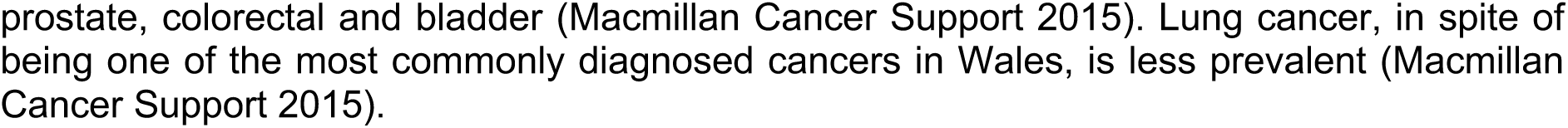
Eligibility Criteria.

| PCC | Inclusion criteria | Exclusion criteria |
| --- | --- | --- |
| **Conditions** | Chronic and long-term conditions<br>1. Coronary Heart Disease<br>2. Cancer*<br>3. Dementia<br>4. Heart failure<br>5. Hypertension<br>6. PVD<br>7. Stroke & TIA<br>8. Atrial fibrillation<br>9. Multi-morbidity<br>10. Diabetes (type 1, type 2, undetermined type)<br>11. Mental health<br>12. Anxiety<br>13. Depression<br>14. Smoking<br>15. Obesity<br>16. Poverty<br>17. Poor Diet | Asthma<br>Chronic Kidney Disease (CKD)<br>Chronic Obstructive Pulmonary Disease (COPD)<br>Epilepsy<br>Rheumatoid Arthritis<br>Influenza<br>Covid and long Covid<br>Other infectious diseases<br>New conditions identified by genomic testing |
| **Context** | Wales data was chosen in the first instance.<br>UK wide data was used if no data available | European data<br>Worldwide data |
| **Population** | Adults over 18 years | Children under 18 years |
| **Study design** | Prevalence and modelling studies of any design |  |
| **Other** | In the past 5 years and then as far back as 2012 depending on the condition and seminal outputs that inform the subject area.<br>English language |  |
| **Other Study Considerations** |  |  |
| Cost data if associated with forecasted prevalence and incidence.<br>Mortality |  |  |

\*The four most common cancers in the UK are breast, prostate, lung, and bowel (Cancer Research UK 2020). However, the most commonly diagnosed cancers are not always the most prevalent ones, as cancer prevalence is influenced by the number of new cases diagnosed (incidence) and mortality (Macmillan Cancer Support 2015). This means that cancers with high incidence but poor prognosis, such as lung cancer, are less prevalent (Macmillan Cancer Support 2015). In Wales, in 2010 the most prevalent cancers were: breast,
prostate, colorectal and bladder (Macmillan Cancer Support 2015). Lung cancer, in spite of being one of the most commonly diagnosed cancers in Wales, is less prevalent (Macmillan Cancer Support 2015).

**Table 3:** Flow of studies through the study selection process.

|  | Diabetes | Dementia | CVD<sup>a</sup> | Smoking<br>Obesity<br>Diet | Mental<br>illness | Multi-<br>morbidity | Cancer<sup>b</sup> |
| --- | --- | --- | --- | --- | --- | --- | --- |
| **Database** | **Hits** |  |  |  |  |  |  |
| Medline | 981 | 289 | 311 | 213 | 17 | 20 |  |
| EMBASE | 2667 | 444 | 728 | 490 | 53 | 47 |  |
| CINAHL | 365 | 146 | 80 | 325 | 88 | 15 |  |
| PsycINFO | NA | 69 | NA |  | 20 | NA |  |
| Scopus | 1,100 | 279 | 22 | 184 | 165 | 2 |  |
| **TOTAL** | **5123** | **1227** | **1141** | **1212** | **298** | **109** |  |
| **Screening processes** |  |  |  |  |  |  |  |
| Limited to 2018 to 2023 | 1600 | NA | NA | NA | NA | NA |  |
| Duplicates removed in endnote | 2993 | 565 | 355 | 359 | 45 | 22 |  |
| Imported into Rayyan | 1600 | 662 | 786 | 853 | 298 | 75 |  |
| Duplicates removed in Rayyan | 115 | 98 | 33 | 85 | 6 | 1 |  |
| Title and abstract screening | 1485 | 564 | 753 | 768 | 292 | 74 |  |
| Full text screening | 12 | 25 | 17 | 24 | 7 | 1 | 17 |
| Included modelling studies after full text screening | **2** | **6** | **6** | **8** | **2** | **0** | **6** |
| Included in Appendix 2 as | 1 | 1 | 2 |  |  |  | **2** |
| other outcome data that was not extracted |  |  |  |  |  |  |  |
| **Supplementary searches** |  |  |  |  |  |  |  |
| Modelling studies identified by back chaining | 1 | 0 | 1 |  | 0 | 0 | 0 |
| Modelling data identified within grey reports | 0 | 2 | 0 | 2 | 0 | 0 | 1 |
| Data from organisation websites | 1 | 0 | 0 | 0 | 0 | 0 | 0 |
| Data from national datasets included | 1 | 2 | 1 | 1 | 1 | 0 | 2 |
| **TOTAL** | **5** | **10** | **9** | **11** | **3** | **1** | **11** |

Key: CVD: cardiovascular diseases; NA: not applicable
<sup>a</sup> searches in this category included atrial fibrillation cardiovascular diseases (coronary heart disease, peripheral vascular disease; stroke and transient ischaemic attacks), heart failure and hypertension.
<sup>b</sup> Data provided in Table 4

**Table 4:**
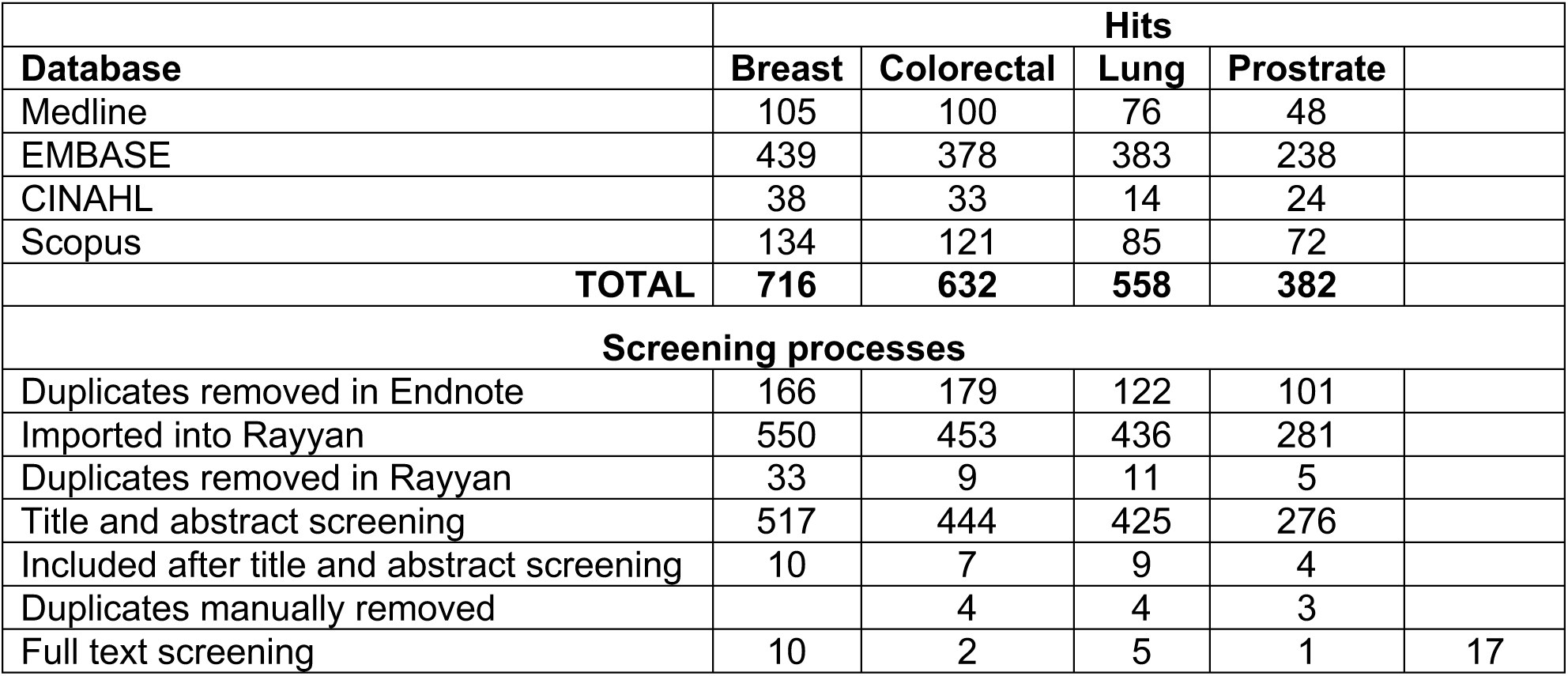
Flow of studies through the study selection process continued.

#### 2.1.1 Atrial fibrillation

Two modelling studies were identified that provided estimates for the projected prevalence of atrial fibrillation (Lane et al. 2017) and atrial fibrillation-related embolic vascular events (Yiin et al. 2014) and full details are provided in Table 5.

**Table 5:** Data extraction table of forecasted incidence / prevalence estimates for atrial fibrillation from modelling studies.

| Citation | Study Characteristics |  | Key Findings |
| --- | --- | --- | --- |
| Lane et al. 2017<br>Temporal trends in incidence, prevalence, and mortality of atrial fibrillation in primary care<br>https://pubmed.ncbi.nlm.nih.gov/28455344/ | <u>Condition/s</u><br>Atrial fibrillation<br><u>Population age (years)</u><br>≥18<br><u>Countries</u><br>UK | <u>Study design</u><br>Modelling study<br><u>Type of model</u><br>Model 1: Age dependent mortality constant incidence<br>Model 2: Constant excess mortality increased incidence<br><u>Data sources</u><br>Primary care databases<br>- CRPD<br>Population-based administration datasets<br>- ONS<br><u>Period (year)</u><br>2040, 2060 | <u>Projected prevalence of AF (number)</u><br>(Model 1 - constant incidence rate from 2010)<br>2010: 718,334<br>2040: 1,085,078 / 2060: 1,258,705<br>(Model 2- constant 1% increase in the incidence rate from 2010)<br>2010: 718,334<br>2040: 1,322,694 / 2060: 1,846,960<br><u>Projected prevalence of AF per 1000 (model 1)</u><br>2010: Overall: 14.5 / Female: 12.9 / Male: 16.3<br>2040: Overall: 20.5 / Female: 18.7 / Male: 22.3<br>2060: Overall: 25.4 / Female: 23.9 / Male: 26.9<br><u>Projected prevalence of AF per 1000 (model 2)</u><br>2010: Overall: 14.5 / Female: 12.9 / Male: 16.3<br>2040: Overall: 23.1 / Female: 20.4 / Male: 25.8<br>2060: Overall: 32.0 / Female: 29.0 / Male: 34.9<br><u>Trend</u><br>Projected increase<br>Prevalence rates are projected to rise faster in men than women |
| Yiin et al. 2014<br>Age-specific incidence, outcome, cost, and projected future burden of atrial fibrillation-related embolic vascular events: a population-based study | <u>Condition/s</u><br>AF related embolic vascular events<br><u>Population age (years)</u><br>≥80 years<br><u>Countries</u><br>UK | <u>Study design</u><br>Modelling study<br><u>Type of model</u><br>Projected likely future numbers of AF-related embolic events using 2002 to 2012 incidence rates from OXVASC and the ONS predictions of UK population structures in 2030 | <u>Projected number of AF-related incident ischemic strokes</u><br>2010: 18,176 (≥80 years)<br>2030: 34,694 (≥80 years)<br>2050: 58,621 (≥80 years)<br>2010: 10,569 (<80 years)<br>2030: 11,777 (<80 years)<br>2050: 12,557 (<80 years)<br><u>Projected number of AF-related incident systemic emboli</u> |
| https://pubmed.ncbi.nlm.nih.gov/25208551/ |  | and 2050 assuming no change in age-specific incidence.<br><u>Data sources</u><br>Prospective cohort study<br>- OXVASC 2002 to 2012<br>Population-based administration datasets<br>- ONS 2012<br><u>Period (year)</u><br>2030, 2050 | 2010: 3,976 (≥80 years)<br>2030: 7,968 (≥80 years)<br>2050: 14,353 (≥80 years)<br>2010: 1537 (≥80 years)<br>2030: 1,710 (<80 years)<br>2050: 1,821 (<80 years)<br><u>Trend</u><br>The projected number of incident AF-related events by 2050 is 87,353 (71,178 ischemic strokes and 16,175 systemic emboli)<br>Numbers of AF-related embolic events at age ≥80 years will treble by 2050 (72,974/year), with 83.5% of all events occurring in this age group.<br><u>Other outcome data (not extracted)</u><br>Costs of care |

Key: AF: Atrial fibrillation; CRPD: Clinical Practice Research Datalink; IMPACT-BAM: ONS: Office for National Statistics, OXVASC: Oxford Vascular Study

None of the studies provided projection estimates for diabetes within Wales, with data only available for the UK for 2030, 2040 and 2060 (Lane et al. 2017; Yiin et al. 2014).

The data sources included prospective cohort studies (for example Oxford Vascular Study (OXVASC) population-based administration datasets, for example ONS population estimates and primary care databases (Clinical Practice Research Datalink). Both studies did not provide specific information other than the data sources used (Lane et al. 2017, Yiin et al. 2014).

#### 2.1.2 Cancer

Five modelling studies were identified that provided estimates for the projected prevalence (Macmillan Cancer Support 2020, Maddams et al. 2012) or incidence of breast, colorectal, lung and/ prostate cancer (Smittenaar et al. 2016, Donnelly et al. 2020, Borras et al. 2016). Two further modelling studies provided estimates for the projected incidence for colorectal (Tsoi et al. 2017) and prostate cancer (Teoh et al. 2019).

Only one study provided projected prevalence estimates for Wales along with separate data for England Scotland and Northern Ireland for 2025, 2030 and 2040. The full details for Wales are provided in Table 6 (Macmillan Cancer Support 2020). Other studies provided projected incidence estimates of breast, colorectal, lung and prostate cancer for Northern Ireland for 2040 (Donnelly et al. 2020), UK for 2030, 2035 and 2040 (Smittenarr et al. 2016), and the UK for 2025 as part of a wider European study (Borras et al. 2016). Two further studies projected UK incidence for 2030 for colorectal (Tsoi et al 2017) and prostate cancer (Macmillan Cancer Support 2020) (Teoh et al 2019) as part of wider global studies. One further study provided projected prevalence estimates for the UK for 2030, 2035, and 2040 (Maddams et al 2012). Further details are provided in Appendix 2.

**Table 6:** Data extraction table of forecasted prevalence/incidence estimates for cancer from modelling studies.

| Citation | Study Characteristics |  | Key Findings |
| --- | --- | --- | --- |
| Macmillan Cancer Support 2020<br>Prevalence by cancer type, nation, sex and year<br>https://www.macmillan.org.uk/dfsmedia/1a6f23537f7f4519bb0cf14c45b2a629/5192-10061/macmillan-2020-cancer-prevalence-figures-and-methodology | <u>Condition/s</u><br>Cancer (all cancers excluding non-melanoma skin cancer)<br><u>Population age (years)</u><br>Any<br><u>Countries</u><br>Wales | <u>Study design</u><br>Modelling study<br><u>Data sources</u><br>Cancer registry data<br>- LCI Wales: 21 year prevalence (1995-2015), provided by WCISU<br><u>Type of model</u><br>Growth rates from Maddams et al. 2012 who used a discrete time model<br><u>Period (year)</u><br>2025, 2030, 2040 | <u>Prevalence - number of people</u><br>Rounded numbers for complete (all-time) prevalence<br>2015: Not reported<br>2025: 200,000 (all)<br>Female: 110,000 / Male: 90,000<br>2030: 230,000 (all)<br>Female: 130,000 / Male: 100,000<br>2040: 300,000 (all)<br>Female: 170,000 / Male: 130,000<br>Rounded numbers for 0-5 years prevalence (people diagnosed in the last 5 years)<br>2015: Not reported<br>2025: 70,000 (all)<br>Female: 30,000 / Male: 30,000<br>2030: 80,000 (all)<br>Female: 40,000 / Male: 40,000<br>2040: 90,000 (all)<br>Female: 50,000 / Male: 50,000<br><u>Trend</u><br>Projected increase |
| Macmillan Cancer Support 2020<br>Prevalence by cancer type, nation, sex and year<br>https://www.macmillan.org.uk/dfsmedia/1a6f23537f7f4519bb0cf14c45b2a629/5192-10061/macmillan-2020-cancer-prevalence-figures-and-methodology | <u>Condition/s</u><br>Breast (female only)<br><u>Population age (years)</u><br>Any<br><u>Countries</u> | <u>Study design</u><br>Modelling study<br><u>Data sources</u><br>Cancer registry data<br>- LCI Wales: 21 year prevalence (1995-2015), provided by WCISU | <u>Prevalence - number of people</u><br>Rounded numbers for complete (all-time) prevalence<br>2015: Not available<br>2025: 51,000<br>2030: 61,000 |
| [92-10061/macmillan-2020-cancer-prevalence-figures-and-methodology](https://www.macmillan.org.uk/dfsmedia/1a6f23537f7f4519bb0cf14c45b2a629/5192-10061/macmillan-2020-cancer-prevalence-figures-and-methodology) | Wales | <u>Type of model</u><br>Growth rates from Maddams et al. 2012 who used a discrete time model.<br><u>Period (year)</u><br>2025, 2030, 2040 | 2040: 85,000<br>Rounded numbers for 0-5 years prevalence (people diagnosed in the last 5 years)<br>2015: Not available<br>2025: 15,000<br>2030: 18,000<br>2040: 23,000<br><u>Trend</u><br>Projected increase |
| Macmillan Cancer Support 2020<br>Prevalence by cancer type, nation, sex and year<br>https://www.macmillan.org.uk/dfsmedia/1a6f23537f7f4519bb0cf14c45b2a629/5192-10061/macmillan-2020-cancer-prevalence-figures-and-methodology | <u>Condition/s</u><br>Colorectal<br><u>Population age (years)</u><br>Any<br><u>Countries</u><br>Wales | <u>Study design</u><br>Modelling study<br><u>Data sources</u><br>Cancer registry data<br>- LCI Wales: 21 year prevalence (1995-2015), provided by WCISU<br><u>Type of model</u><br>Growth rates from Maddams et al. 2012 who used a discrete time model.<br><u>Period (year)</u><br>2025, 2030, 2040 | <u>Prevalence - number of people</u><br>Rounded numbers for complete (all-time) prevalence<br>2015: Not available<br>2025: 22,000 (all)<br>Female: 9,000 / Male: 13,000<br>2030: 27,000 (all)<br>Female: 11,000 / Male: 16,000<br>2040: 36,000 (all)<br>Female: 14,000 / Male: 22,000<br>Rounded numbers for 0-5 years<sup>a</sup> prevalence (people diagnosed in the last 5 years)<br>2015: Not available<br>2025: 9,000 (all)<br>Female: 4,000 / Male: 6,000<br>2030: 11,000 (all)<br>Female: 4,000 / Male: 7,000<br>2040: 13,000 (all)<br>Female: 5,000 / Male: 9,000<br><u>Trend</u> |
|  |  |  | Projected increase |
| Macmillan Cancer Support 2020<br>Prevalence by cancer type, nation, sex and year<br>https://www.macmillan.org.uk/dfsmedia/1a6f23537f7f4519bb0cf14c45b2a629/5192-10061/macmillan-2020-cancer-prevalence-figures-and-methodology | <u>Condition/s</u><br>Lung<br><u>Population age (years)</u><br>Any<br><u>Countries</u><br>Wales | <u>Study design</u><br>Modelling study<br><u>Data sources</u><br>Cancer registry data<br>- LCI Wales: 21 year prevalence (1995-2015), provided by WCISU<br><u>Type of model</u><br>Growth rates from Maddams et al. 2012 who used a discrete time model.<br><u>Period (year)</u><br>2025, 2030, 2040 | <u>Prevalence - number of people</u><br>Rounded numbers for complete (all-time) prevalence<br>2015: Not available<br>2025: 6,000 (all)<br>Female: 3,510 / Male: 2,260<br>2030: 7,000 (all)<br>Female: 4,410 / Male: 2,300<br>2040: 9,000 (all)<br>Female: 6,610 / Male: 2,320<br>Rounded numbers for 0-5 years prevalence (people diagnosed in the last 5 years)<br>2015: Not available<br>2025: 4,000 (all)<br>Female: 2,270 / Male: 1,470<br>2030: 4,000 (all)<br>Female: 2,790 / Male: 1,480<br>2040: 5,000 (all)<br>Female: 3,950 / Male: 1,410<br><u>Trend</u><br>Projected increase |
| Macmillan Cancer Support 2020<br>Prevalence by cancer type, nation, sex and year<br>https://www.macmillan.org.uk/dfsmedia/1a6f23537f7f4519bb0cf14c45b2a629/5192-10061/macmillan-2020-cancer-prevalence-figures-and-methodology | <u>Condition/s</u><br>Prostate (male only)<br><u>Population age (years)</u><br>Any<br><u>Countries</u><br>Wales | <u>Study design</u><br>Modelling study<br><u>Data sources</u><br>Cancer registry data<br>- LCI Wales: 21 year prevalence (1995-2015), provided by WCISU<br><u>Type of model</u> | <u>Prevalence - number of people</u><br>Rounded numbers for complete (all-time) prevalence<br>2015: Not available<br>2025: 34,000<br>2030: 42,000<br>2040: 56,000<br>Rounded numbers for 0-5 years prevalence |
|  |  | Growth rates from Maddams et al. 2012 who used a discrete time model.<br><u>Period (year)</u><br>2025, 2030, 2040 | (people diagnosed in the last 5 years)<br>2015: Not available<br>2025: 13,000<br>2030: 15,000<br>2040: 17,000<br><u>Trend</u><br>Projected increase |

Key: LCI: Local Cancer Intelligence, WCISU: Welsh Cancer Intelligence and Surveillance Unit

The data source for the Welsh modelling study conducted by Macmillan Cancer Support (2020) was cancer registry data, with growth rates taken Maddams et al. (2012) who used a discrete time model in their prevalence projections. Macmillan Cancer Support (2020) did not provide any further detail about the modelling methods used.

Additionally, UK datasets were identified that provided projected incidence estimates for all cancers, including breast (female only), colorectal, lung, and prostate (Cancer Research UK 2023a, Cancer Research UK 2023b, Cancer Research UK 2023c, Cancer Research UK 2023e). The data sources were projected and observed cancer incidence from National Cancer Registration and Analysis Service, ISD Scotland, Welsh Cancer Intelligence and Surveillance Unit (WCISU), and Northern Ireland Cancer Registry. The modelling methods were described as an age-period-cohort modelling approach (Cancer Research UK 2023d). Moreover, One National (Wales) database for Social Care Data was found that contained projections of cancer prevalence (for all cancers) across different age groups up to 2043 for each local authority in Wales (Social Care Wales 2023). Separate projections were available for different survival periods, such as diagnosed with cancer over one year to five years ago (Social Care Wales 2023).

Three additional studies were identified that explored mortality rates (data extraction not conducted) and details are provided in Appendix 2.

#### 2.1.3 Cardiovascular diseases

Six modelling studies were identified that provided estimates for the projected prevalence or incidence of cardiovascular diseases (Collins et al. 2022, Guzman-Castillo et al. 2017, Kingston et al. 2018), stroke (King et al. 2020, Li et al. 2020, Kingston et al. 2018), myocardial infarction (MI) (Scarborough et al. 2022) and coronary heart disease (CHD)(Kingston et al. 2018) and full details are provided in Tables 7 and 11.

**Table 7:** Data extraction table of forecasted prevalence and incidence for cardiovascular diseases from modelling studies.

| Citation | Study Characteristics |  | Key Findings |
| --- | --- | --- | --- |
| Collins et al. 2022<br>What will the cardiovascular disease slowdown cost? Modelling the impact of CVD trends on dementia, disability, and economic costs in England and Wales from 2020-2029<br>https://pubmed.ncbi.nlm.nih.gov/35767575/ | <u>Condition/s</u><br>CVD<br><u>Population age (years)</u><br>35–100 years<br><u>Countries</u><br>England and Wales (Combined) | <u>Study design</u><br>Modelling study<br><u>Type of model</u><br>Stochastic Markov model<br>IMPACT-BAM<br><u>Data sources</u><br>Prospective cohort study<br>- ELSA<br>Population-based administration datasets<br>- ONS population estimates 2016<br>- NHS HES 2018/2019<br><u>Period (year)</u><br>2029 | <u>Denominator population data for 2011</u><br>Not reported<br><u>Scenario 1: CVD plateau - annual CVD incidence remains at 2011 levels</u><br>Remains at approximately 1,200 per 100,000 people<br><u>Scenario 1: CVD plateau - CVD prevalence remains at 2011 levels</u><br>Increase slightly over time to approximately 9%<br><u>Scenario 2: CVD fall - CVD incidences continue to decline</u><br>Decline to < 800 per 100,000<br><u>Scenario 2: CVD fall - CVD prevalence continues to decline</u><br>Fall to approximately 6%<br><u>Other outcome data (not extracted)</u><br>QALYs<br>Healthcare costs<br><u>Trend</u><br>After previous, dramatic falls, CVD incidence and mortality have recently plateaued in the UK |
| Guzman-Castillo et al. 2017<br>Forecasted trends in disability and life expectancy in England and Wales up to 2025: a modelling study.<br>https://pubmed.ncbi.nlm.nih.gov/28736759/ | <u>Condition/s</u><br>CVD-related disability<br><u>Population age (years)</u><br>35-100<br><u>Countries</u><br>England and Wales (Combined) | <u>Study design</u><br>Modelling study<br><u>Type of model</u><br>IMPACT-BAM<br>Markov Model<br><u>Data sources and dates</u> | <u>Projected prevalence – number of people (% change since 2015)</u><br><u>CVD-related disability</u><br>>65 years<br>2015: 588,000 (95% UI: 576,000 to 599,000)<br>2025: 527,000 (95% UI: 505,000 to 547,000)<br>–10·3% (95% UI: –13·6 to 7·3)<br>65 to 84 years |
|  |  | Population-based administration datasets<br>- ONS population estimates 2016<br>Prospective cohort study<br>- ELSA<br><u>Period (year)</u><br>2025 | 2015: 419,000 (95% UI: 410,000 to 428,000)<br>2025: 348,000 (95% UI: 335,000 to 363,000)<br>-16.9% (95% UI: -19.8 to 3.9)<br>>85 years<br>2015: 168,000 (95% UI: 163,000 to 174,000)<br>2025: 179,000 (95% UI: 166,000 to 190,000)<br>6.0% (95% UI: -0.8 to 12.6)<br><u>Trends</u><br>In people aged over 65 to 84 years, the numbers of CVD-related disability will decline from 2015 to 2025.<br>In people aged 85 years or older the numbers of CVD-related disability will increase from 2015 to 2025<br><u>Other data (not extracted)</u><br>Total life expectancy<br>Disability free life expectancy<br>Life expectancy with disability |
| King et al. 2020<br>The future incidence, prevalence and costs of stroke in the UK<br>https://pubmed.ncbi.nlm.nih.gov/31957781/ | <u>Condition/s</u><br>Stroke<br><u>Population age (years)</u><br>≥45 Years<br><u>Countries</u><br>UK | <u>Study design</u><br>Modelling study<br><u>Type of model</u><br>Cell-based simulation model<br><u>Data sources</u><br>Population-based administration datasets<br>- ONS 2014<br>Prospective cohort studies<br>Consultation exercise with 11 stroke experts<br><u>Period (year)</u><br>2025, 2035 | <u>Projected prevalence– number of stroke survivors (% change since 2015)</u><br><u>Total all strokes</u><br>2015: 950,200<br>2025: 1,424,100<br>2035: 2,119,400 (123%)<br><u>Females: Ischaemic stroke</u><br>2015: 397,300<br>2025: 594,800 / 2035: 912,500<br><u>Females: Haemorrhagic stroke</u><br>2015: 41,800<br>2025: 62,700 / 2035: 96,200<br><u>Males: Ischaemic stroke</u><br>2015: 462,500<br>2025: 693,300 / 2035: 1,004,300<br><u>Males: Haemorrhagic stroke</u><br>2015: 48,600 |
|  |  |  | 2025: 73,300 / 2035: 106,300<br><u>Total Ischaemic stroke</u><br>2015: 859,800<br>2025:1,288,100 / 2035:1,916,900<br><u>Total Haemorrhagic stroke</u><br>2015: 90,400<br>2025:136,100 / 2035:202,600<br><u>Incidence – number of new strokes</u><br>(% change since 2015)<br><u>Total all strokes</u><br>2015: 117,600<br>2025: 148,700<br>2035: 186,900 (59%)<br><u>Females: Ischaemic stroke</u><br>2015: 53,700<br>2025: 66,400 / 2035: 84,300<br><u>Females: Haemorrhagic stroke</u><br>2015: 6,400<br>2025: 7,900 / 2035:10,000<br><u>Males: Ischaemic stroke</u><br>2015: 51,700<br>2025: 66,600 / 2035:82,900<br><u>Males: Haemorrhagic stroke</u><br>2015: 5,900<br>2025:7,700 / 2035: 9,800<br><u>Total Ischaemic stroke</u><br>2015: 105,300<br>2025:133,000 / 2035:167,200<br><u>Total Haemorrhagic stroke</u><br>2015: 12,300<br>2025:15,700 / 2035: 19,700<br><u>Other outcome data (not extracted)</u><br>Health and social care costs |

|  |  |  | Trend<br>Projected increase in prevalence and incidence |
| --- | --- | --- | --- |
| Li et al. 2020<br>Trends in stroke incidence in high-income countries in the 21st Century: population-based study and systematic review<br>https://pubmed.ncbi.nlm.nih.gov/32208842/ | <u>Condition/s</u><br>Stroke<br><u>Population age (years)</u><br>All ages<br><u>Countries</u><br>UK | <u>Study design</u><br>Modelling study<br><u>Type of model</u><br>To estimate the number of incident stroke patients on a national basis in the UK from 2015 up to 2045, we used the age-specific incidence during 2014 to 2017 in OXVASC for standardization to the projected UK population.<br><u>Data sources</u><br>Prospective cohort studies<br>- OXVASC;2014–2017<br>Population-based administration datasets<br>- ONS 2016<br><u>Period (year)</u><br>2045 | <u>Denominator population data for 2015</u><br>Not reported<br><u>% change in incidence of new strokes since 2015</u><br><u>Scenario 1: if age-specific incidence remained stable over the next 30 years</u><br>2045: increase by 66%<br><u>Scenario 2: if age-specific stroke incidence continued to decline by 6% every 5 years</u><br>2045: increase by 13%<br><u>Trend</u><br>With the projected aging population If the age-specific incidence remained stable over the next 30 years, the number of incident stroke would increase by 66% from year 2015 to 2045.<br>With the projected aging population, if the age-specific stroke incidence continued to decline at a rate of 6% every 5 years, there would still be a 13% increase of the number of first-ever strokes in the UK by 2045. |
| Scarborough et al. 2022<br>Forecast of myocardial infarction incidence, events and prevalence in England to 2035 using a microsimulation model with endogenous disease outcomes<br>https://pubmed.ncbi.nlm.nih.gov/35771859/ | <u>Condition/s</u><br>Myocardial infarction<br><u>Population age (years)</u><br>55-85+years<br>(Four age groups: 55–64, 65–74, 75–84 and 85+)<br><u>Countries</u><br>England | <u>Study design</u><br>Modelling study<br><u>Type of model</u><br>microPRIME model (including microsimulation)<br><u>Data sources</u><br>Population-based administration datasets<br>- ONS mortality data<br>National Health Surveys<br>- HSE (1993 to 2017) | <u>Projected incidence for first MI (events per 100,000)</u><br>Females<br>2011: not reported<br>2035: 55-64 / 65-74 / 85+ years: no extractable data<br>2035: 75–84 years: 666 (95% UI: 427–949) per 100,000<br>Males<br>2011: not reported<br>2035: 55–64 years: 592 (95% UI: 395–813) per 100,000<br>2035: 65-74 / 75–84 / 95+ years: no extractable data<br><u>Modelled age-standardised incidence for MI (1<sup>st</sup> and subsequent MI)</u> |
|  |  | <u>Period (year)</u><br>2035 | Females: 482 (95% UI: 351–666) per 100,000<br>Males: 1042 (95% UI: 761–1371) per 100,000<br><u>Modelled prevalence rates from 2011</u><br>Females<br>55–64 and 65–74 years: decline by 2035<br>75–84 years: no description<br>85+ years: plateau by 2035 after initial increase<br>Males<br>55–64 and 65–74 years: decline by 2035<br>75–84 and 85+ years: increase by 2035<br><u>Trend</u><br>There may be an increase in incidence from MI in males but increases for women are unlikely.<br>Prevalence rates may fall in older men, and are likely to remain stable in women over the next decade and a half |

Key: CHD; Coronary heart disease; CRPD: clinical practice research datalink; CVD: Cardiovascular disease; ELSA: English Longitudinal Study of Ageing; HSE: Health Survey for England; IMPACT-BAM: IMPACT-Better Ageing Model; ONS: Office for National Statistics, OXVASC: Oxford Vascular Study; NHS HES: National Health Service Hospital Episode Statistics; UI: Uncertainty Interval. \*NHS HES was used for Wales only

**Table 8:** Data extraction table of forecasted prevalence for dementia.

| Citation | Study Characteristics |  | Key Findings |
| --- | --- | --- | --- |
| Ahmadi-Abhari et al. 2017<br>Temporal trend in dementia incidence since 2002 and projections for prevalence in England and Wales to 2040: modelling study<br>https://pubmed.ncbi.nlm.nih.gov/28679494/ | <u>Condition/s</u><br>Dementia<br><u>Population age (years)</u><br>>50 years<br><u>Countries</u><br>England and Wales (combined) | <u>Study design</u><br>Modelling study<br><u>Type of model</u><br>IMPACT-BAM<br>Markov Model<br><u>Data sources</u><br>Prospective cohort study - ELSA<br><u>Period (year)</u><br>2030, 2040 | <u>Prevalence - number of people</u><br>2016:766,600 (95% UI 735,200 – 796,900)<br>2030:1,091,600 (95% UI 1,034,200 to 1,146,200)<br>2040: 1,204,500 (95% UI 1,101,000 to 1,296 200)<br><u>Prevalence (%)</u><br>2016: 3.5% (95% UI: 3.4% to 3.7%)<br>2030: 4.3% (95% UI: 4.1% to 4.5%)<br>2040: 4.4% (95% UI: 4.0% to 4.7%)<br><u>Trend</u><br>Projected increase - the number of people with dementia is projected to increase.<br>Age specific dementia incidence is declining, and it is estimated to decline by about 21% from 2016 to 2040.<br>A sensitivity analysis without the incidence decline gave a much larger projected growth, of more than 1.9 million people with dementia in 2040.<br>The crude prevalence of dementia in population aged 50 or more and 65 or more is estimated to increase up to 2040 in men, whereas in women it declines after 2025 |
| Bandosz et al. 2019<br>Potential impact of diabetes prevention on mortality and future burden of dementia and disability: a modelling study<br>https://pubmed.ncbi.nlm.nih.gov/31732789/ | <u>Condition/s</u><br>Dementia<br><u>Population age (years)</u><br>≥65<br><u>Countries</u><br>England and Wales (combined) | <u>Study design</u><br>Modelling study<br><u>Type of model</u><br>IMPACT-BAM<br>Markov Model<br><u>Data sources and dates</u> | <u>Dementia incident cases (number of people)</u><br>2015: 142,000 (95% UI: 137,000 to 147,000)<br>2030: 187,000 (95% UI: 179,000 to 194,000)<br>2045: 209,000 (95% UI: 197,000 to 219,000)<br>2060: 204,000 (95% 189,100 to 218,000)<br><u>Data for men</u><br>2015: 61,000 (95% UI: 57,000 to 66,000) |
|  |  | Population-based administration datasets<br>- ONS population estimates 2016<br>Prospective cohort study<br>- ELSA<br><u>Period (year)</u><br>2030, 2045, 2060 | 2030: 86,000 (95% UI: 80,000 to 92,000)<br>2045: 99, 000 (95% UI: 91,000 to 107, 000)<br>2060: 99,000 (95% UI: 88,000 to 109,000)<br><u>Data for women</u><br>2015: 81,000 (95% UI 79,000 to 83,000)<br>2030: 101,000 (95% UI 97,000 to 104, 000)<br>2045: 110,000 (95% UI: 105,000 to 114,000)<br>2060: 105,000 (95% UI 99,000 to 110,000)<br><u>Trend</u><br>This increase in the number of annual incident cases occurred despite a continuous decline in the incidence rates of dementia. This suggests that the reason for the increasing number of projected new cases of dementia is the projected ageing of the population, due to increasing longevity |
| Guzman-Castillo et al. 2017<br>Forecasted trends in disability and life expectancy in England and Wales up to 2025: a modelling study<br>https://pubmed.ncbi.nlm.nih.gov/28736759/ | <u>Condition/s</u><br>Dementia related disability<br>Dementia and CVD related disability<br>CVD-related disability<br><u>Population age (years)</u><br>35-100<br><u>Countries</u><br>England and Wales (Combined) | <u>Study design</u><br>Modelling study<br><u>Type of model</u><br>IMPACT-BAM<br>Markov Model<br><u>Data sources and dates</u><br>Population-based administration datasets<br>- ONS population estimates 2016<br>Prospective cohort study<br>-ELSA<br><u>Period (year)</u><br>2025 | <u>Projected prevalence – number of people (% change since 2015)</u><br><u>Dementia related disability</u><br>≥65 years<br>2015: 468,000 (95% UI: 447,000 to 491,000)<br>2025: 699,000 (95% UI: 654,000 to 745,000)<br>49.1% (95% UI: 41.5 to 56.2)<br>65 to 84 years<br>2015: 289,000 (95% UI: 269,000 to 309,000)<br>2025: 405,000 (95% UI: 374,000 to 436,000)<br>40.3% (95% UI: 34.7 to 46.3)<br>≥85 years<br>2015: 179,000 (95% UI: 171,000 to 188,000)<br>2025: 293,000 (95% UI: 267,000 to 320,000)<br>63.1% (95% UI: 50.7 to 76.5) |
|  |  |  | <u>Dementia and CVD-related disability</u><br>>65 years<br>2015: 177,000 (95% UI: 171,000 to 183,000)<br>2025: 191,000 (95% UI: 178,000 to 203,000)<br>7.7% (95% UI: 1.3 to 14.0)<br>65 to 84 years<br>2015: 84,000 (95% UI: 79,000 to 88,000)<br>2025: 79,000 (95% UI: 73,000 to 85,000)<br>–5.3% (95% UI: –10.8 to 0.7)<br>>85 years<br>2015: 93,000 (95% UI: 89,000 to 97,000)<br>2025: 111,000 (95% UI: 102,000 to 120,000)<br>19.3% (95% UI: 9.8 to 29.0)<br><u>CVD-related disability</u><br>>65 years<br>2015: 588,000 (95% UI: 576,000 to 599,000)<br>2025: 527,000 (95% UI: 505,000 to 547,000)<br>–10.3% (95% UI: –13.6 to 7.3)<br>65 to 84 years<br>2015: 419,000 (95% UI: 410,000 to 428,000)<br>2025: 348,000 (95% UI: 335,000 to 363,000)<br>–16.9% (95% UI: –19.8 to 3.9)<br>>85 years<br>2015: 168,000 (95% UI: 163,000 to 174,000)<br>2025: 179,000 (95% UI: 166,000 to 190,000)<br>6.0% (95% UI: –0.8 to 12.6)<br><u>Trends</u><br>Projected increase in dementia-related disability cases across all age groups<br>In people aged over 65 to 84 years, the numbers of CVD-related disability and dementia and CVD-related disability will decline from 2015 to 2025. |
|  |  |  | The numbers of dementia-related disability will however increase for people aged over 65 to 84 years.<br>In people aged 85 years or older, the numbers of dementia and CVD related disability, dementia-related disability, and CVD-related disability will increase from 2015 to 2025<br><u>Other data (not extracted)</u><br>Total life expectancy<br>Disability free life expectancy<br>Life expectancy with disability |
| Li et al. 2021<br>Model-Based projection of dementia prevalence in China and Worldwide 2020-2050<br>https://pubmed.ncbi.nlm.nih.gov/34219732/ | <u>Condition/s</u><br>Dementia<br><u>Population age (years)</u><br>>60<br><u>Countries</u><br>UK<br>(data collected as part of a wider global study) | <u>Study design</u><br>Modelling study<br><u>Type of model</u><br>Based on Brookmeyer and Grays method<br><u>Data sources and dates</u><br>Population-based administration datasets<br>- ONS population estimates (no date given)<br>Prospective cohort studies<br>- CFAS I and II<br>Modelling study<br>- Ahmadi-Abhari et al. 2017<br><u>Period (year)</u><br>2050 | <u>Projected prevalence – number of people (% change since 2020)</u><br>2020: 1,100,000<br>2050:1,820,000 (65.93%)<br><u>Trend</u><br>Projected increase from 2020 |
| Prince et al. 2014<br>Dementia UK update | <u>Condition/s</u><br>Dementia<br><u>Population age (years)</u><br>>30 | <u>Study design</u><br>Modelling study<br><u>Type of model</u> | <u>Projected prevalence – number of people (% change since 2013)</u><br>2013: 815,827<br>2025: 1,142,677 (41%) |
| https://kclpure.kcl.ac.uk/portal/files/35828472/P326_AS_Dementia_Report_WEB2.pdf | <u>Countries</u><br>UK | Updated version of a model of the costs and outcomes of dementia that builds upon previous versions of the PSSRU aggregate long-term care model and the PSSRU dementia care model<br><u>Data sources and dates</u><br>Population-based administration datasets<br>- ONS population estimates 2013<br>Prospective cohort studies<br><u>Period (year)</u><br>2025, 2051 | 2051: 2,092,945 (175%)<br><u>Trend</u><br>Projected increase from 2013<br><u>Other data (not extracted)</u><br>Early onset dementia<br>Late onset dementia<br>Figure B (projected increases in number of people with dementia in the UK by age)<br>Figure C (projected increases in the number of people in the UK by gender)<br>Page xiv |
| Wittenberg et al. 2019<br>Projections of older people with dementia and costs of dementia care in the United Kingdom 2019-2040<br>https://www.lse.ac.uk/cpec/assets/documents/cpec-working-paper-5.pdf | <u>Condition/s</u><br>Dementia<br><u>Population age (years)</u><br>>65<br><u>Countries</u><br>Wales<br>England<br>Scotland<br>Northern Ireland | <u>Study design</u><br>Modelling study<br><u>Type of model</u><br>Updated version of model developed by CPEC the MODEM study<br><u>Data sources and dates</u><br>Population-based administration datasets<br>- ONS population estimates 2014<br>Modelling data<br>- PACSim model<br><u>Period (year)-</u><br>2025, 2030, 2040 | <u>Denominator population data for 2015</u><br>Not reported<br><u>Projected prevalence – number of people (% change since 2019)</u><br><u>England</u><br>2019: 748,000<br>2025: 898,500<br>2030: 1,046,100<br>2040: 1,352,400 (81%)<br><u>Wales</u><br>2019: 46,800<br>2025: 55,570<br>2030: 64,200<br>2040: 79,700 (70%)<br><u>UK</u><br>2019: 883,100<br>2025: 1,060,100<br>2030: 1,233,400<br>2040: 1,590,100 (80%) |
|  |  |  | <u>Trend</u><br>The number of people with dementia in Wales is projected to rise by 70% from 2019 to 2040<br>The prevalence rate of dementia in the UK is projected to reach 8.8% in 2040 (up from 7.1% in 2019)<br>The prevalence rate is projected to increase from 7.2% in 2019 to 8.3% in 2040 in England, from 6.3% to 7.7% in Scotland, from 7.0% to 9.0% in Wales, and from 6.9% to 8.5% in Northern Ireland<br><u>Other data (not extracted)</u><br>Projected number of older people with mild, moderate and severe dementia<br>Data from Figure 2 – Proportional increase in the number of older people by age groups between 2019 and 2040<br>Data from Table 2 - Costs of dementia for older people<br>Data from Figure 6 –Breakdown of total costs of dementia care in the UK, 2040<br>Data from Figure 7 - Breakdown of total costs of dementia care in the four countries, 2040 |
| Wittenberg et al. 2020<br>Projections of care for older people with dementia in England: 2015 to 2040<br>https://www.ncbi.nlm.nih.gov/pmc/articles/PMC7047814/ | <u>Condition/s</u><br>Dementia<br><u>Population age (years)</u><br>65+<br><u>Countries</u><br>England | <u>Study design</u><br>Modelling study<br><u>Type of model</u><br>Cell-based model in Excel with three parts which uses the PACSim dynamic microsimulation model<br><u>Data sources and dates</u><br>Population-based administration datasets<br>- ONS population estimates 2014 | <u>Projected prevalence – number of people (% change since 2015)</u><br><u>65+</u><br>2015: 9,710,000<br>2040: 15,293,000 (57%)<br><u>85+</u><br>2015: 1,297,000<br>2040: 3,083,000 (138%)<br><u>Number of older people with dementia</u><br>2015: 651,000 |
|  |  | Prospective cohort studies<br>- CFAS II<br>- MODEM<br><u>Period (year)</u><br>2040 | 2040: 1,351,000 (108%)<br><u>Other data (not extracted)</u><br>Table 2- Projected number of older people with mild, moderate and severe dementia<br><u>Trend</u><br>Projected increase from 2015 to 2040 |

Key: CEPC: Care Policy and Evaluation Centre; CFAS: Cognitive Function and Ageing Studies; CVD: Cardiovascular disease; ELSA: English Longitudinal Study of Ageing; IMPACT-BAM: IMPACT-Better Ageing Model, MODEM: Modelling Outcome and Cost Impacts of Interventions for Dementia study; ONS: Office for National Statistics; PACSim: Population Ageing and Care Simulation; UI: uncertainty interval; PSSRU: Personal Social Services Research Unit.

**Table 9:**
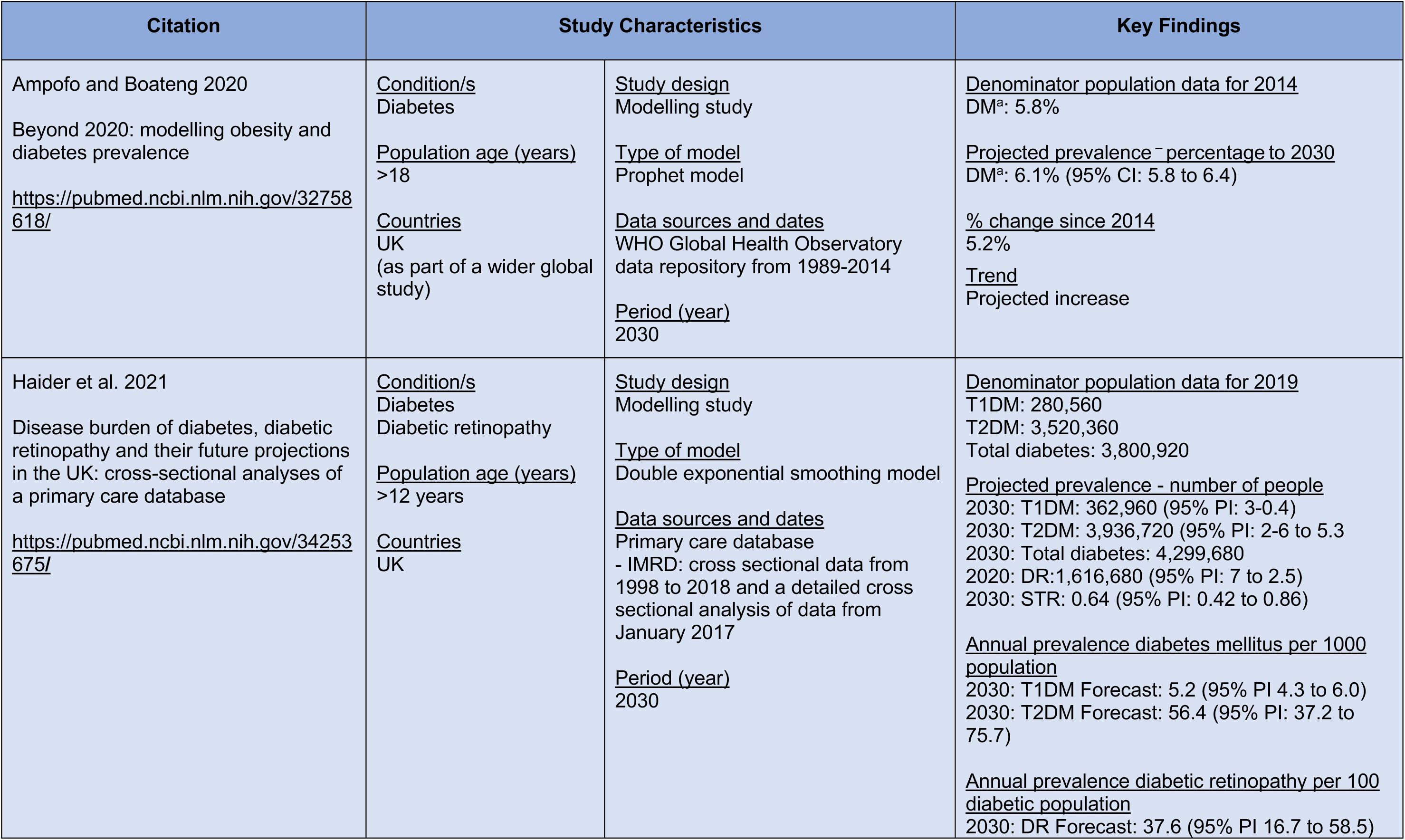
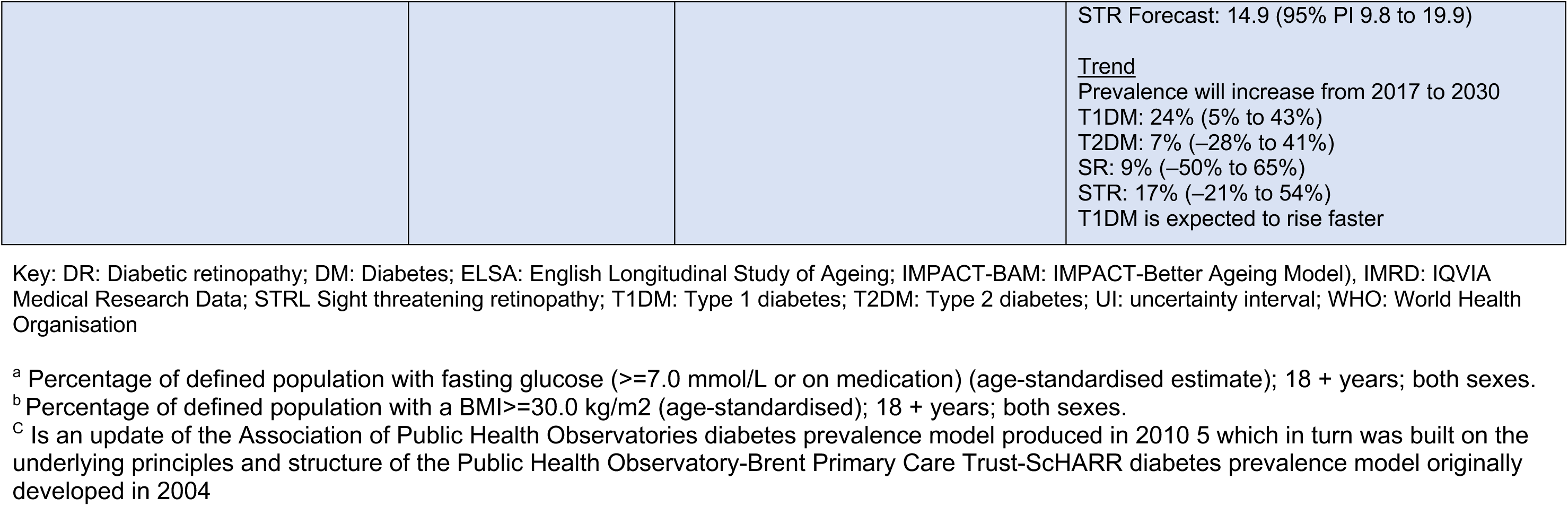
Data extraction table of forecasted prevalence for diabetes.

**Table 10:** Data extraction table of forecasted prevalence/incidence estimates for mental health.

| Citation | Study Characteristics |  | Key Findings |
| --- | --- | --- | --- |
| McDonald et al. 2021<br>Using epidemiological evidence to forecast population need for early treatment programmes in mental health: a generalisable Bayesian prediction methodology applied to and validated for first-episode psychosis in England<br>https:// | <u>Conditions</u><br>Psychosis care<br><u>Population (years)</u><br>16-64<br><u>Countries</u><br>England | <u>Study design</u><br>Modelling study<br><u>Type of model</u><br>Population-based prediction model (Bayesian prediction methodology)<br><u>Data Sources</u><br>Prospective cohort studies<br>- AESOP<br>- ELFEP<br>- SEPEA<br><u>Denominator population</u><br>2017<br><u>Period (year)</u><br>2019 to 2025 | <u>Denominator population data for 2017</u><br>16-64 years:<br>Treated: 9,346 / Probable: 8,038<br>16-35 years:<br>Probable: 5919 / Treated: Not reported<br>36-64 years<br>Probable: 2119 / / Treated: Not reported<br><u>Predicted probable and treated FEP incidence</u><br><u>16-64 years</u><br>2019: Treated<sup>a</sup>: 10,425 (95% CI: 9,766 to 11,060)<br>2019: Probable<sup>b</sup>: 8,987 (95% CI: 8,419 to 9,535)<br>2025: Treated: 11,067 (95% CI: 10,383 to 11,740)<br>2025: Probable: 9,541 (95% CI: 8,951 to 10,120)<br><u>16-35 years</u><br>2019: Treated: 6,683 (95% CI: 6,309 to 7,077)<br>2019: Probable: 5,761 (95% CI: 5,439 to 6,101)<br>2025: Treated: 7,041 (95% CI: 6,641 to 7,453)<br>2025: Probable: 6,070 (95% CI: 5,725 to 6,425)<br><u>36-64 years</u><br>2019: Treated: 3,742 (95% CI: 3,327 to 4,174)<br>2019: Probable: 3,226 (95% CI: 2,868 to 3,598)<br>2025: Treated: 4,026 (95% CI: 3,567 to 4,496)<br>2025: Probable: 3,471 (95% CI: 3,075 to 3,876)<br><u>% change from 2019 to 2025</u><br><u>16-64 years</u><br>Treated: 6.2% (95% CI: -3 to 15.6)<br>Probable: 6.4% (95% CI: -3.0 to -15.4)<br><u>16-35 years</u> |
|  |  |  | Treated: 5.6 (95% CI: -2.8 to 14.6)<br>Probable: 5.5 (95% CI: -2.7 to 13.9)<br><u>38-64 years</u><br>Treated: 7.8 (95% CI: -9.8 to 24.7)<br>Probable: 8.4 (95% CI: -7.4 to 26.8)<br><u>Data not extracted</u><br>Referred to EIP services and accepted by EIP assessment<br><u>Trend</u><br>Between 2019 and 2025, predicted treated caseloads increased by 6.2%, on average, but the authors could not exclude the possibility of no change or a small decline in case-loads over time (95% CI -3.2% to 15.6%) |

AESOP: The Aetiology and ethnicity in schizophrenia and other psychoses; EIP: early intervention psychosis; ELFEP: The East London first episode psychoses; FEP: first episode psychosis; SEPEA: Social epidemiology of psychoses in East Anglia
a - Treated: Treated by EIP services by annum; b - Probable: Probable FEP. (Based on PsyMaptic-A model 4)

**Table 11:** Data extraction table of forecasted prevalence/incidence estimates for multimorbidity’s.

| Citation | Study Characteristics |  | Key Findings |
| --- | --- | --- | --- |
| Kingston et al. 2018<br>Projections of multi-morbidity in the older population in England to 2035: estimates from the Population Ageing and Care Simulation (PACSim) model<br>https://doi.org/10.1093/ageing/afx201 | <u>Conditions</u><br>Multi-morbidity:<br>Chronic diseases and geriatric conditions<br>Chronic Diseases:<br>CHD, stroke, hypertension, diabetes, arthritis, cancer, respiratory disease, dementia, depression,<br>Geriatric Conditions:<br>hearing impairment, vision impairment and cognitive impairment (no dementia)<br><u>Population age (years)</u><br>Base population used: 35+<br>Projections for 65+<br><u>Countries</u><br>England | <u>Study design</u><br>Modelling study<br><u>Type of model</u><br>Dynamic microsimulation model, Population Ageing and Care Simulation (PACSim)<br><u>Data sources</u><br>Prospective cohort studies<br>- Understanding Society, wave 1<br>- ELSA<br>- CFAS<br><u>Period (year)</u><br>2025, 2035 | <u>Projected prevalence percentage (and numbers)</u><br>Cancer (Age 65+):<br>2015: 12.6% (1,224,900)<br>2025: 19.6% (2,297,700) / 2035: 23.7% (3,422,000)<br>% change recorded: 2015-2035: 179.4<br>CHD (Age 65+):<br>2015: 18.3% (1,778,700)<br>2025: 16.6% (1,937,800) / 2035: 15.0% (2,172,500)<br>% change recorded: 2015-2035: 22.1%<br>Dementia (Age 65+):<br>2015: 6.8% (659,700)<br>2025: 7.8% (918,800) / 2035: 8.5% (1,227,500)<br>% change recorded: 2015-2035: 86.1%<br>Depression (Age 65+):<br>2015: 2.3% (225,700)<br>2025: 1.3% (155,500) / 2035: 1.3% (191,600)<br>% change recorded: 2015-2035: -15.1%<br>Diabetes (Age 65+):<br>2015: 14.7% (1,428,400)<br>2025: 19.8% (2,317,900) / 2035: 21.6% (3,115,400)<br>% change recorded: 2015-2035: 118.1%<br>Hypertension (Age 65+):<br>2015: 49% (4,768,200)<br>2025: 54.9% (6,423,400) / 2035: 55.9% (8,080,400)<br>% change recorded: 2015-2035: 69.5%<br>Stroke (Age 65+):<br>2015: 7.5% (726,100)<br>2025: 8.7% (1,021,700) / 2035: 9.3% (1,337,500)<br>% change recorded: 2015-2035: 84.2%<br>2 more multi-morbidities (Age 65+)<br>2015: 54% (5,250,500) |
|  |  |  | 2025: 64.4% (7,535,600) / 2035: 67.8% (9,789,100)<br>% change recorded: 2015-2035: 86.4%<br>4 or more multi-morbidities (Age 65+)<br>2015: 9.8% (952,400)<br>2025: 14.9% (1,749,900) / 2035: 17.0% (2,453,200)<br>% change recorded: 2015-2035: 157.6%<br>2 or more multi-morbidities (Age 85+)<br>2015: 68.7% (905,200)<br>2025: 87.3% (1,539,400) / 2035: 90.5% (2,548,800)<br>% change recorded: 2015-2035: 181.6%<br>4 or more multi-morbidities (Age 85+)<br>2015: 14.9% (196,000)<br>2025: 33.3% (586,900) / 2035: 39.7% (1,117,500)<br>% change recorded: 2015-2035: 470.2% |

Key: CFAS: Cognitive Function and Ageing Study; CHD: Coronary heart disease; ELSA: English Longitudinal Study of Ageing; IMPACT-BAM: IMPACT-Better Ageing Model; UI: uncertainty interval

None of the modelling studies provided projection estimates for diabetes within Wales, with data only available for England for 2035 (Kingston et al. 2018, Scarborough et al. 2022), England and Wales (combined) for 2025 and 2029 (Collins et al. 2022; Guzman-Castillo et al. 2017), and the UK for 2025, 2029, 2035 and 2045 (King et al. 2020; Li et al. 2020). One further study provided projected prevalence estimates for the UK which was collected as part of a wider global study for 2030 (Ampofo and Boateng 2020).

The data sources included prospective cohort studies (for example English Longitudinal Study of Ageing (ELSA), Oxford Vascular Study (OXVASC); population-based administration datasets, for example ONS population estimates, hospital episode statistics (HES) and National Health Survey data (Health Survey for England (HSE); and consultation with experts. Different models were used across the studies including IMPACT-Better Ageing Model (Markov model) (Collins et al. 2022; Guzman-Castillo et al. 2017) a cell based simulation model (King et al. 2020), microsimulation models (microPRIME (Scarborough et al. 2022) and PACSim (Kingston et al. 2018)), an age dependent mortality constant incidence model (Lane et al. 2017) and a constant excess mortality increased incidence model (Lane et al. 2017).

Additionally, one National (Wales) dataset was identified that provided projected prevalence estimates for any heart condition excluding high blood pressure (adults in Wales who self-reported ever having been treated for a heart attack, or currently being treated for angina, heart failure or another heart conditions) and stroke (adults who self-reported ever having been treated for stroke) (Public Health Wales Observatory 2018b). The data sources were Welsh Heart Survey data, and the modelling methods was described as using Daffodil projections which applies Wales-level prevalence figures to projected population estimates for the areas in question to give a projected number of people who will have these conditions in the future (Public Health Wales Observatory 2017b).

Two additional modelling studies were identified for CHD that explored mortality rates (data extraction not conducted) and details are provided in Appendix 2.

#### 2.1.4 Dementia

Eight modelling studies were identified that provided estimates for the projected prevalence of dementia and full details are provided in Table 8 (Ahmadi-Abhari et al. 2017, Bandosz et al. 2020, Guzman-Castillo et al. 2017, Li et al. 2021, Prince et al. 2014, Wittenberg et al. 2019, Wittenberg et al. 2020, Kingston et al. 2018).

Only one study provided projected prevalence estimates for Wales along with separate data for England, Scotland, and Northern Ireland for 2025, 2030 and 2040 (Wittenberg et al. 2019) and full details are provided in Table 8. Other studies provided projected prevalence estimates for England for 2035 (Kingston et al. 2018) and 2040 (Wittenberg et al. 2020); England and Wales (combined) for 2025, 2030, 2040, 2045 and 2060 (Ahmadi-Abhari et al. 2017; Bandosz et al. 2019; Guzman-Castillo et al. 2017); and UK for 2025, and 2051 (Prince et al. 2014). One further study provided projected prevalence estimates for the UK which was collected as part of a wider global study for 2060 (Li et al. 2021).

The data sources included prospective cohort studies (for example ELSA, Cognitive Function and Ageing Studies (CFAS) I and II, Modelling Outcome and Cost Impacts of Interventions for Dementia (MODEM) study; population-based administration datasets, for example ONS population estimates, and other modelling data. Different models were used across the studies including IMPACT-Better Ageing Model (Markov model) (Ahmadi-Abhari et al. 2017, Bandosz et al. 2019, Guzman-Castillo et al. 2017). Brookmeyrer and Gray’s method (Li et al. 2021), an updated model of the costs and outcomes of dementia model which builds on the previous Personal Social Services Research Unit (PSSRU) aggregate long-term care model and the PSSRU dementia care model (Prince et al. 2014), an updated version of the model developed by the Care Policy and Evaluation Centre; the MODEM study (Wittenberg et al. 2019); Population Ageing and Care Simulation (PACSim) microsimulation (Kingston et al. 2018); and a cell-based model in Excel which included PACSim to estimate the number of older people with dementia (Wittenberg et al. 2020).

Additionally, one National (Wales) dataset was identified that provided projected prevalence estimates for dementia (estimated number of people with dementia based on the ‘Expert Delhi Consensus’) (Public Health Wales Observatory 2018b). The data sources were provided by Alzheimer’s and the modelling methods was described as using Daffodil projections which applies Wales-level prevalence figures to projected population estimates for the areas in question to give a projected number of people who will have these conditions in the future (Public Health Wales Observatory 2017b). One other National (Wales) database for Social Care Data was also found that contained projections of dementia prevalence for mild, moderate and severe cases up to 2043 for each local authority in Wales (Social Care Wales 2023).

One additional study was identified that explored disability adjusted life years (data extraction not conducted) and details are provided in Appendix 2.

#### 2.1.5 Diabetes

Three modelling studies were identified that provided estimates for the projected prevalence of diabetes and full details are provided in Table 9 (Ampofo & Boateng 2020, Haider et al. 2021, Kingston et al. 2018).

None of the studies provided projection estimates for diabetes within Wales, with data only available for England for 2025, 2030, 2035 (Kingston et al. 2018) and the UK for 2030 (Haider et al. 2021). One further study provided projected prevalence estimates for the UK which was collected as part of a wider global study for 2030 (Ampofo and Boateng 2020).

The data sources included National Health Surveys (HSE), population-based administration datasets, for example ONS population estimates and HES data), primary care databases (IQVIA Medical Research Data) and the WHO global health observatory data repository. Across the studies different models were utilised including, a double exponential smoothing model (Haider et al. 2021), Prophet model (Ampofo and Boateng 2020) and the PACSim microsimulation model (Kingston et al. 2018).

One National (Wales) dataset was identified that provided projected prevalence estimates for diabetes (adult respondents were asked whether they were currently being treated for diabetes, making no distinction between type 1 and type 2 diabetes) (Public Health Wales Observatory 2018b). The data sources were provided by the Welsh Health Survey and Welsh Government Daffodil projections which applies Wales-level prevalence figures to projected population estimates for the areas in question to give a projected number of people who will have these conditions in the future (Public Health Wales Observatory 2017b). Additionally, the web site of Diabetes UK provided a projection prevalence estimate but no further details were provided (Diabetes UK 2023a).

One additional scenario-based modelling study was identified (data extraction not conducted) and details are provided in Appendix 2.

#### 2.1.6 Heart failure

We did not find any modelling studies or grey literature reports for the projected prevalence or incidence of HF.

#### 2.1.7 Hypertension

One modelling study was identified for projecting prevalence of hypertension, although this only contained data for England (Kingston et al. 2018). This study used the PACSim microsimulation model to make projections for hypertension up to 2035 (Kingston et al. 2018).

#### 2.1.8 Mental illness

One modelling study was identified that provided estimates for the projected prevalence of psychotic disorders (McDonald et al. 2021) and a further modelling study was identified for depression (Kingston et al. 2018) full details are provided in Table 10. We did not find any modelling studies or grey literature reports for the projected prevalence or incidence of common mental disorders (anxiety) or bipolar disorder.

None of the studies provided projection estimates for mental illness within Wales, with data only available for England for 2025 and 2035 (Kingston et al. 2018; McDonald et al. 2021).

The data sources for the two studies included prospective cohort studies, such as AESOP, ELFEP, SEPEA, ELSA, and Understanding Society wave 1. Different models were used across the two studies, including PACSim microsimulation model (Kingston et al. 2018) and a population-based prediction model using Bayesian prediction methodology (McDonald et al. 2021).

One National (Wales) database for Social Care Data was found that contained projections of mental disorder prevalence across different age groups up to 2043 for each local authority in Wales (Social Care Wales 2023). Separate projections were available for bipolar disorder, personality disorder, psychotic disorder, and common mental health conditions (Social Care Wales 2023).

#### 2.1.9 Multi-morbidities

One modelling study was identified that provided estimates for the projected prevalence of multimorbidity (Kingston et al. 2018) and full details are provided in Table 11. None of the studies provided projection estimates for multi-morbidities within Wales, with data only available for England for 2025 and 2035. The data sources were prospective cohort studies and the Dynamic microsimulation model and the Population Ageing and Care Simulation (PACSim) model.

#### 2.1.10 Obesity

Nine modelling studies (reported across 10 publications) were identified that provided estimates for the projected prevalence of obesity (Bhimjiyani et al. 2016a, Bhimjiyani et al. 2016b, Ampofo & Boateng 2020, Cobiac & Scarborough 2021, Janssen et al. 2020, Keaver et al. 2019, Pérez-Ferrer et al. 2018, Pineda et al. 2018, World Obesity Federation 2023) and/ or morbid obesity (Keaver et al. 2020) and full details are provided in Table 12.

**Table 12:** Data extraction table of forecasted prevalence for obesity.

| Citation | Study Characteristics |  | Key Findings |
| --- | --- | --- | --- |
| Bhimjiyani et al. 2016a<br>Tipping the scales: Why preventing obesity makes economic sense<br>https://ukhealthforum.org.uk/wp-content/uploads/2019/02/Tipping-The-Scales-CRUK-Full-Report-FINAL.pdf<br>Bhimjiyani et al. 2016b<br>Extended obesity technical report: baseline scenario, hypothetical scenarios, SSB excise tax policy scenario<br>https://ukhealthforum.org.uk/wp-content/uploads/2018/11/CRUK-UKHF-Extended-Obesity-Technical-Report.pdf | <u>Condition/s</u><br>Obesity ($\geq 30$ kg.m<sup>2</sup>)<br><u>Population age (years)</u><br>18 to 100<br><u>Countries</u><br>England | <u>Study design</u><br>Modelling study<br><u>Type of model</u><br>UKHF model (dual-module modelling process)<br>-Module one: non-linear multivariate, categorical regression model fitted to cross-sectional risk factor data to create longitudinal projections to 2035<br>-Module two: microsimulation (Markov-type simulations) as a tool for predicting disease burden using longitudinal projections from module one<br><u>Data sources</u><br>National Health Survey<br>-HSE 2000 and 2012<br><u>Period (year)</u><br>2035 | <u>Projected prevalence of obesity (percentage)</u><br>2015: Female 30% / Male 29%<br>2025: Female 34% / Male 34%<br>2030: Female 37% / Male 36%<br>2040: Female 40% / Male 39%<br><u>Trend</u><br>Projected increase<br>Female: increase from 30% (2015) to 40% (2035) in women<br>Male: increase from 29% (2015) to 39% (2035) in men<br>Obesity is projected to increase across all age and income groups, apart from 30-39 year old men where trends are expected to be stable over time<br>Obesity is projected to increase most in the lower income groups<br><u>Data not extracted</u><br>Detailed BMI projections by age, sex and income quantile<br>Obesity prevalence projections by 1%, 10%, 20%, 50%, 100% BMI-related diseases<br>Results from the SSB tax scenario - the introduction of a 20% excise tax on sugary sweetened beverages |
| Cobiac and Scarborough 2021<br>Modelling future trajectories of obesity and body mass index in England<br>https://pubmed.ncbi.nlm.nih.gov/34077469/ | <u>Condition/s</u><br>Obesity and morbid obesity<br><u>Population age (years)</u><br>$\geq 18$<br><u>Countries</u> | <u>Study design</u><br>Modelling study<br><u>Type of model</u><br>Linear model (best-case future scenario)<sup>a</sup><br>Non-linear model (worst-case future scenario)<sup>b</sup> | <u>Denominator population data for 2017</u><br>Not reported<br><u>(% change in population at increased risk (BMI$\geq 25$kg/m<sup>2</sup>) since 2017)</u><br>Best-case scenario:<br>Projected decline from 66% to 53% (95% CI: 41% to 64%) |
|  | England | Monte Carlo cross validation approach<br><u>Data sources</u><br>National Health Survey<br>-HSE 1991 to 2017<br><u>Period (year)</u><br>2035 | Worst-case scenario:<br>Projected to remain stable at 64%<br>(95% CI: 37% to 90%)<br><u>Projected prevalence of obesity (percentage)</u><br>Best-case scenario: Males<br>15.6% (95% CI:12.6 to 18.6) Obese<br>7.82% (95% CI: 3.27 to 12.4) Very obese<br>Best-case scenario: Females<br>14.7% (95% CI:12.6 to 16.8) Obese<br>10.4% (95% CI: 6.22 to 14.6 ) Very obese<br>Worst-case scenario: Males<br>20.1% (95% CI:16.5 to 23.7) Obese<br>13.2% (95% CI: 2.73 to 23.7 Very Obese<br>Worst-case scenario: Females<br>17.9% (95% CI:14.8 to 21.1) Obese<br>15.7% (95% CI: 6.53 to 24.9) Very Obese<br><u>Trend</u><br>The linear models predict a substantial decline in the proportion of the population at increased risk of obesity-related disease (i.e., classified in our modelling as Overweight, Obese or Very obese). The non-linear models predict no or little decrease on average. From an average value of 66% in 2017, the modelling predicts that by 2035 this proportion will at best reduce to 53% (41% to 64%) or at worst remain relatively stable at 64% (37% to 90%) |
| Janssen et al. 2020<br>Obesity prevalence in the long-term future in 18 European countries and in the USA<br>https://pubmed.ncbi.nlm.nih.gov/33075798/ | <u>Condition/s</u><br>Obesity ($\geq 30$ kg.m<sup>2</sup>)<br><u>Population age (years)</u><br>20–84<br><u>Countries</u><br>UK<br>(as part of a wider global study) | <u>Study design</u><br>Modelling study<br><u>Type of model</u><br>Bayesian hierarchical model<br><u>Data sources</u><br>Population-based administration datasets<br>-NCD-RisC 2017 data | <u>Denominator population data for 2016</u><br>UK Males: 29.3<br>UK Females: 31.3<br><u>Projected age-standardized obesity prevalence (percentage)</u><br>UK: 2060 (males): 24.1 (95% PI: 21.6; 27.7)<br>UK: 2060 (females): 25.7 (95% PI: 23.4; 28.9)<br><u>Projected maximum levels of age-standardized obesity prevalence (percentage)</u> |
|  |  | <u>Period (year)</u><br>2060<br>2033 & 2034<br>(projected years that the maximum obesity prevalence will be reached for women and men respectively) | UK: (males): 36.9 (95% PI: 35.6; 38.8)<br>Projected year that the maximum will be reached and 95% projection intervals is 2034 (2032, 2038)<br>UK: 2033 (females): 36.9 (95% PI: 35.9; 38.5)<br>Projected year that the maximum will be reached and 95% projection intervals 2033 (2031, 2037)<br><u>Projected annual change in obesity prevalence from 2016 to peak year (in percentage points)</u><br>Males: 0.42 / Females 0.33<br><u>Projected annual change in obesity prevalence from peak year to 2060 (in percentage points)</u><br>Males: -0.49 / Females -0.42<br><u>Trend</u><br>Obesity in the UK is expected to reach maximum levels between 2032 and 2038 among men, and between 2031 and 2037 among women<br>In 2060, obesity prevalence is expected to decline |
| Keaver et al. 2019<br>Future trends in social inequalities in obesity in England, Wales and Scotland<br>https://pubmed.ncbi.nlm.nih.gov/30855666/ | <u>Condition/s</u><br>Overweight (25-25.99 kg.m<sup>2</sup>)<br>Obesity (≥30 kg.m<sup>2</sup>)<br><u>Population age (years)</u><br>≥16<br><u>Countries</u><br>England, Wales and Scotland | <u>Study design</u><br>Modelling study<br><u>Type of model</u><br>Non-linear multivariate regression model<br><u>Data sources</u><br>National Health Survey<br>-Welsh Health Survey (2004–14)<br><u>Period (year)</u><br>2035 | <u>Denominator population data for 2005</u><br>Prevalence (percentage) of obesity by occupational status for Wales<br>Routine and manual<br>Male: 21.2 / Female: 23.5<br>Intermediate:<br>Male: 19.5 / Female: 18.1<br>Managerial<br>Male: 17.5 / Female: 17.0<br>Prevalence (percentage) of obesity by educational level for Wales<br>Less than tertiary<br>Male: 20 / Female: 20.5<br>Tertiary |
|  |  |  | Male: 14 / Female: 14<br><u>Projected prevalence (percentage) of obesity by occupational status level for Wales in 2035</u><br>Routine and manual<br>Male: 40.1 / Female: 43.0<br>Intermediate:<br>Male: 38.9 / Female: 36.9<br>Managerial<br>Male: 32.1 / Female: 29.9<br><u>Percentage change in projected prevalence of obesity from 2005 by occupational status education level for Wales</u><br>Routine and manual<br>Male: 89.2 / Female: 83<br>Intermediate:<br>Male: 99.5 / Female: 103.9<br>Managerial<br>Male: 83.4 / Female: 75.9<br><u>Projected prevalence (percentage) of obesity by educational level for Wales in 2035</u><br>Less than tertiary<br>Male: 35.9 / Female: 40.3<br>Tertiary<br>Male: 26.2 / Female: 26.2<br><u>Percentage change in projected prevalence of obesity from 2005 by education level for Wales</u><br>Less than tertiary<br>Male: 79.6 / Female: 97.0<br>Tertiary<br>Male: 87.4 / Female: 87.4<br><u>Trend</u><br>Obesity prevalence in Wales is projected to across from 2005 across all levels of education and occupational categories by 2023 |
| Keaver et al. 2020<br>Morbid obesity in the UK: A modelling projection study to 2035<br>https://pubmed.ncbi.nlm.nih.gov/30160640/ | <u>Condition/s</u><br>Morbid obesity (BMI $\geq 40$ kg/m<sup>2</sup>)<br><u>Population age (years)</u><br>$\geq 15$<br><u>Countries</u><br>England, Wales and Scotland | <u>Study design</u><br>Modelling study<br><u>Type of model</u><br>Categorical multivariate non-linear regression model<br><u>Data sources</u><br>National Health Survey<br>-Welsh Health Survey (2004–14)<br><u>Period (year)</u><br>2025, 2030, 2035 | <u>Projected prevalence of morbid obesity in Wales (percentage)</u><br>2015: 3 (+ 95% CI 0.03)<br>2025: 6 (+ 95% CI 0.26)<br>2030: 8 (+ 95% CI 0.40)<br>2035: 11 (+ 95% CI 0.54)<br>(Equates to a population figure of 340,899 people being classed as morbidly obese in Wales in 2035.<br>By 2023 Welsh women aged 55–64 years are projected to have the had the highest prevalence of 20%.<br><u>Percentage change in projected prevalence of morbid obesity from 2015 in Wales</u><br>Overall:<br>2015: 3% / 2035: 11% (8% increase – equating to 340,899)<br>Males:<br>2015: 2% / 2035: 13% (11% increase)<br>Females:<br>2015: 3% / 2035: 9% (6% increase)<br><u>Trend</u><br>Projected increase<br>The prevalence of morbid obesity is predicted to increase by 2035 across the three UK countries, with Wales projected to have the highest rates |
| Pérez-Ferrer et al. 2018<br>Inequalities in smoking and obesity in Europe predicted to 2050: Findings from the EConDA project<br>https://pubmed.ncbi.nlm.nih.gov/29516788/ | <u>Condition/s</u><br>Obesity (BMI $\geq 30$kg/m<sup>2</sup>)<br><u>Population age (years)</u><br>$\geq 20$ (for England)<br><u>Countries</u><br>England<br>(as part of a wider European study) | <u>Study design</u><br>Modelling<br><u>Type of model</u><br>Nonlinear multivariate regression model<br><u>Data sources</u><br>National Health Survey<br>-HSE 2000 to 2013 | <u>Predicted prevalence ratio of obesity (percentage)</u><br>England<br>2015 (Females): 0.29 (95% PI: 0.26, 0.32)<br>2030 (Females): 0.36 (95% PI: 0.29, 0.43)<br>2035 (Females): 0.38 (95% PI: 0.29, 0.47)<br>2040 (Females): 0.40 (95% PI: 0.29, 0.51)<br>2045 (Females): 0.42 (95% PI: 0.29, 0.55)<br>2050 (Females): 0.45 (95% PI: 0.34-0.59)<br>England<br>2015 (Males): 0.29 (95% PI: 0.26, 0.32) |
|  |  | <u>Period (year)</u><br>2025, 2030, 2035, 2040, 2045, 2050 | 2030 (Males): 0.38 (95% PI :0.3, 0.46)<br>2035 (Males): 0.41 (95% PI : 0.32, 0.50)<br>2040 (Males): 0.44 (95% PI : 0.33, 0.55)<br>2045 (Males): 0.47 (95% PI : 0.34, 0.60)<br>2050 (Males): 0.49 (95% PI: 0.35, 0.63)<br><u>Trend</u><br>Projected decline |
| Pineda et al. 2018<br>Forecasting future trends in obesity across Europe: the value of improving surveillance<br>https://pubmed.ncbi.nlm.nih.gov/30308509/ | <u>Condition/s</u><br>Obesity (BMI ≥ 30kg/m<sup>2</sup>)<br><u>Population age (years)</u><br>≥ 20<br><u>Countries</u><br>Wales<br>(As part of a wider European study) | <u>Study design</u><br>Modelling study<br><u>Type of model</u><br>Non-linear categorical regression model<br><u>Data sources</u><br>Population-based administration datasets<br>-WHO BMI database<br>-National statistical databases<br>-Public health institutions and health surveillance reports<br>-PubMed and Science Direct databases<br>Between 2000 and August 2015<br><u>Period (year)</u><br>2025 | <u>Denominator population data</u><br>Not reported<br><u>Projected prevalence of obesity (percentage)</u><br>2025: Wales: 28% (95% CI 20; 36)<br><u>Trend</u><br>In Wales the prevalence was predicted to increase from 2015 to 20–30% in 2025 |
| World Obesity Federation 2023<br>World Obesity Atlas<br>https://s3-eu-west-1.amazonaws.com/wof- | <u>Condition/s</u><br>Obesity (BMI ≥ 30kg/m<sup>2</sup>)<br><u>Population age (years)</u><br>Adults: ≥20<br><u>Countries</u> | <u>Study design</u><br>Modelling study<br><u>Type of model</u><br>Not reported<br><u>Data sources</u> | <u>Denominator population data</u><br>Not reported<br><u>Projected prevalence of obesity (percentage)</u><br>2035: 46%<br><u>Data not extracted</u> |
| files/World_Obesity_Atlas_2023_Report.pdf | UK<br>(as part of a wider global study) | Population-based administration datasets<br>-NCD-RisC<br><u>Period (year)</u><br>2035 | Projected Healthcare impact of BMI $\geq 25\text{kg/m}^2$, US\$ million in 2020, 2025, 2030, and 2035<br>Projected total economic impact of BMI $\geq 25\text{kg/m}^2$, US\$ million in 2020, 2025, 2030, and 2035<br>Projected impact of BMI $\geq 25\text{kg/m}^2$ on GDP in 2020, 2025, 2030, and 2035<br><u>Trend</u><br>There will be 2.4% annual increase in obesity between 2020 and 2035 |

Key: BMI: body mass index, CI: confidence intervals, HSE: Health Survey for England; NCD-RisC: Non-communicable Diseases Risk Factor Collaboration; PI: projection intervals, WHO: World Health Organisation
<sup>a</sup> allowing for a quadratic relationship between the outcome variable and time, to reflect a potential reversal in upwards trends in BMI distribution
<sup>b</sup> applied an exponential function to reflect a potential flattening of trends in BMI distribution over time

Three studies provided projections for Wales (Keaver et al. 2019, Keaver et al. 2020, Pineda et al. 2018), one of these included Wales as part of a wider European study (Pineda et al. 2018). Three studies had UK data as part of a wider global study (Janssen et al. 2020, Ampofo & Boateng 2020, World Obesity Federation 2023), while three studies reported obesity data for England (Bhimjiyani et al. 2016a, Bhimjiyani et al. 2016b, Cobiac & Scarborough 2021, Pérez-Ferrer et al. 2018) one of which provided data for England as part of a wider European study (Pérez-Ferrer et al. 2018).

The data sources included National Heath Surveys (HSE, Welsh Health Survey) and population-based administration datasets (WHO Global Health Observatory data, NCD-RisC 2017 data, WHO BMI database). Different modelling methods were applied across the included studies, such as the Prophet model (Ampofo & Boateng 2020), the UKHF model which is a duel module modelling process including nonlinear multivariate regression and microsimulation (Bhimjiyani et al. 2016a, Bhimjiyani et al. 2016b), Bayesian hierarchical model (Janssen et al. 2020), non-linear multivariate regression model (Pérez-Ferrer et al. 2018, Keaver et al. 2019), and categorical multivariate nonlinear regression model (Keaver et al. 2020, Pineda et al. 2018). Cobiac & Scarborough (2021) used a linear model to predict best-case future scenarios, non-linear model for worst case future scenarios and a Monte Carlo cross validation approach. One study did not detail the model used for projecting prevalence (World Obesity Federation 2023).

Additionally, one National (Wales) dataset was identified that provided projected prevalence estimates for obesity (self-reported height and weight which was used to calculate BMI and overweight as a BMI of 25+ and obese as a BMI of 30+) (Public Health Wales Observatory 2018b). The data sources were provided by the Welsh Health Survey and Welsh Government and the modelling methods were provided as a hyperlink to a website that no longer exists (Public Health Wales Observatory 2017b).

#### 2.1.11 Poor diet/nutrition

We did not find any modelling studies or grey literature reports for the projected prevalence or incidence of poor diet/nutrition.

#### 2.1.12 Smoking

One modelling study was identified that provided estimates for the projected prevalence of smoking (Pérez-Ferrer et al. 2018) and full details are provided in Table 13. This study only included England as part of a wider European study. The data source for this study was a National Health Survey (a general lifestyle survey), and a nonlinear multivariate regression model was used to make projections.

**Table 13:** Data extraction table of forecasted prevalence for smoking.

| Citation | Study Characteristics |  | Key Findings |
| --- | --- | --- | --- |
| Pérez-Ferrer et al. 2018<br>Inequalities in smoking and obesity in Europe predicted to 2050: Findings from the EConDA project<br>https://pubmed.ncbi.nlm.nih.gov/29516788/ | <u>Condition/s</u><br>Smoking<br><u>Population age (years)</u><br>≥20 (for England)<br><u>Countries</u><br>England<br>(As part of a wider European study) | <u>Study design</u><br>Modelling<br><u>Type of model</u><br>Nonlinear multivariate regression model<br><u>Data sources</u><br>National Health Survey<br>-General lifestyle survey<br>2000 to 2011<br><u>Period (year)</u><br>2020, 2025, 2030, 2035, 2040, 2045, 2050 | <u>Predicted prevalence of smoking</u><br>England<br>2015 (Females): 0.17 (95% PI: 0.16, 0.18)<br>2030 (Females): 0.11 (95% P: 0.09, 0.13)<br>2035 (Females): 0.10 (95% P: 0.08, 0.12)<br>2040 (Females): 0.09 (95% P: 0.06, 0.12)<br>2045 (Females): 0.08 (95% PI: 0.05, 0.11)<br>2050 (Females): 0.07 (95% PI: 0.04, 0.10)<br>England<br>2015 (Males): 0.18 (95% PI: 0.17, 0.19)<br>2030 (Males): 0.12 (95% PI: 0.1, 0.14)<br>2035 (Males): 0.11 (95% PI: 0.08, 0.14)<br>2040 (Males): 0.09 (95% PI: 0.06, 0.12)<br>2045 (Males): 0.08 (95% PI: 0.05, 0.11)<br>2050 (Males): 0.07 (95% PI: 0.04, 0.10)<br><u>Data not extracted</u><br>2020, 2025<br><u>Trend</u><br>Projected decline |

Key: PI: projection intervals

Additionally, one National (Wales) dataset was identified that provided projected prevalence estimates for smoking (estimated smoking prevalence projection based on 15 year olds already smoking, 16+ year old starting to smoke and the 16+ population that quit each year. (Public Health Wales Observatory 2018b). The data sources were provided by the Welsh Health Survey, Welsh Government and the ONS. The modelling methods were based of smoking prevalence projections in Scotland (Public Health Wales Observatory 2017b).

### 2.2 Summary of current and projected prevalence and/or incidence data

This section is organised by the included long-term conditions – AF, CVD (CHD, PVD, stroke and TIAs), cancer (breast, colorectal and lung, prostate), dementia, diabetes, HF, hypertension, mental illness (anxiety, bipolar disorder, depression, psychosis, schizophrenia), and multi-morbidities. Three risk factors for long term conditions were also included, namely poor diet/nutrition, obesity and smoking. Global, UK (or English) and Welsh incidence and/or prevalence and wider risk factors for each condition and the three specific risks are presented from the wider literature. The most recent (current) global, UK and Welsh prevalence / incidence estimates are provided where available from the wider literature. Projected prevalence and/or incidence estimates are provided where available from the evidence retrieved for this rapid evidence map.

#### 2.2.1 Atrial fibrillation

##### Current global incidence

- Atrial fibrillation is the most frequent cardiac arrhythmia worldwide with 37.6 million cases (GBD 2017 Disease and Injury Incidence and Prevalence Collaborators 2018).
- There are almost 5 million new cases of AF and flutter diagnosed each year.
- The age-adjusted rate per 100,000, per year for all ages and sexes is 57.09 (uncertainty interval (44.07 to 71.90) (World Stroke Organisation 2022).

##### Current prevalence in the UK

- In the UK, there are an estimated 1,496,972 individuals living with AF (Public Health England 2019).
- Data from the Quality and Outcomes Framework (QOF) prevalence for AF (all ages) for 2021/2022 for England was 1,288,599 which demonstrates an increasing trend over time (Office for Health Improvement & Disparities 2022a).

##### Current prevalence / incidence in Wales

- Incidence rates for AF in 2017 per 100,000 of the adult population aged 15+ was 358.5 (men 434.0 and women 292.7) (British Heart Foundation 2020).
- Data from the Quality Assurance and Improvement Framework (QAIF) disease register for 2019/2020 estimates that estimates that around 79,000 people in Wales have been diagnosed with AF (British Heart Foundation 2023).
- Data provided by the British Heart Foundation (BHF) demonstrated that the prevalence of AF in Wales increased from 1.6% in 2006/2007 to 2.3% in 2018/2019 (British Heart Foundation 2020).

##### Projected prevalence / incidence in the UK

- Projections of AF prevalence demonstrated a constant yearly rise, increasing from 700,000 patients in 2010 to between 1.3 and 1.8 million patients with AF by 2060 (Lane et al. 2017).
- Prevalence rates are projected to rise faster in men than women (Lane et al. 2017).
- The projected number of incident AF-related events by 2050 is 87,353 (71,178 ischemic strokes and 16,175 systemic emboli). The numbers of AF-related embolic events at age ≥80 years will treble by 2050 (72,974/year), with 83.5% of all events occurring in this age group (Yiin et al. 2014).

##### Projected prevalence in Wales

- We did not find any modelling studies that estimated the projected prevalence of AF in Wales.

##### Risk factors

- There are a number of risk factors that increase the likelihood of a person developing AF, these include, age, ethnicity, diabetes, hypertension, obesity and congestive heart failure AF (Public Health England 2017).
- While several studies point to increased risk of AF with several risk factors, the main risks appear to be ageing, being male, and being Caucasian (Public Health England 2017).
- With the exception of ethnicity, the estimates presented here attempt to take into account the biggest predictors of AF (Public Health England 2017).

#### 2.2.2 Cancer

##### Breast cancer

###### Current global incidence and prevalence

- It is estimated that around 2.3 million new breast cancer (female only) cases were diagnosed in 2020, contributing 11.7% of all cancers globally (Sung et al. 2021).
- Prevalence (5-year) was estimated to be around 7.8 million worldwide in 2020 (Globocan 2020).

###### Current Incidence in the UK

- Based on Globocan (2020), the estimated number of new breast cancer (female only) cases in 2020 in the UK was 53,889.

###### Current incidence in Wales

- In 2019, 2,790 women were newly diagnosed with breast cancer in Wales (WCISU 2022).

###### Projected prevalence in the UK

- It was estimated that there will be approximately 1.2 million females living with a breast cancer diagnosis in the UK by 2030 and this number is projected to increase to approximately 1.68 million by 2040 (Maddams et al. 2012).

###### Projected incidence in the UK

- It was estimated that the projected yearly average number of new cases of female breast cancer will increase to 69,900 in 2038-2040 from 61,800 in 2023-2025 in the UK (Cancer Research UK 2023b).
- Projections for breast cancer incidence rates indicate a 1% increase between 2023-2025 and 2038-2040 to 175 cases per 100,000 females yearly average by 2038-2040 (Cancer Research UK 2023b).

###### Projected prevalence in Wales

- It was estimated that there will be 61,000 females living with a breast cancer diagnosis in Wales by 2030 and this number is projected to increase to approximately 85,000 by 2040 (Macmillan Cancer Support 2020).

###### Risk factors

- Older age is one of the main risk factors of being diagnosed with breast cancer (Cancer Research UK 2018b).
- Biological processes and exposure to risk factors can both contribute to the development of breast cancer (Cancer Research UK 2018).
- Biological processes include, endogenous factors (sex hormone levels, breast density), presence of in situ carcinoma, and reproductive factors (older age at first giving birth, younger age at menarche, older age at menopause) (Cancer Research UK 2018).
- Genetic risk factors can also lead to development of breast cancer, with over 90% of inherited cases caused by BRCA1 and BRCA2 mutations (NICE 2018).
- Regarding, exposure to risk factors, it is estimated that about 23% of breast cancer cases in UK might be preventable (Cancer Research UK 2018).
- Risk factors with ‘sufficient’ or ‘convincing’ evidence include alcohol consumption, hormone replacement therapy (HRT) (oestrogen-progestogen), oestrogen-progestogen contraceptives, diethylstilbesterol, exposure to ionising radiation, body fat levels (for post-menopausal breast cancer only), adult attained height or weight gain (Cancer Research UK 2018).
- For other risk factors there is ‘limited’ or ‘probable’ evidence, including Digoxin, HRT (oestrogen-only), ethylene oxide, polychlorinated biphenyls, shift work that could cause circadian disruption, smoking, body fat levels (for pre-menopausal breast cancer only), and higher birth weight (Cancer Research UK 2018).
- Certain reproductive factors, such as reproductive organ surgery or having children can decrease the chance of breast cancer (Cancer Research UK 2018).
- There is also ‘limited’ or ‘probable’ evidence that breastfeeding, physical activity and vigorous exercise can decrease the risk of breast cancer (Cancer Research UK 2018).

##### Colorectal cancer

###### Current global incidence and prevalence

- It is estimated that around 1.9 million new colorectal cancer cases were diagnosed in 2020, contributing 10% of all cancers globally (Sung et al. 2021, Globocan 2020).
- Prevalence (5-year) was estimated to be around 5.3 million worldwide in 2020 (Globocan 2020).

###### Current incidence in the UK

- Based on Globocan (2020), the estimated number of new colorectal cancer cases in 2020 in the UK was 52,128.

###### Current incidence in Wales

- In 2019, 2,513 people were newly diagnosed with colorectal cancer (1,404 male and 1,109 female) (WCISU 2022).

###### Projected prevalence in the UK

- It was estimated that there will be approximately 274,000 males and 200,000 females living with a diagnosis of colorectal cancer by 2030 and this number is projected to increase to approximately 377,000 and 255,000, respectively by 2040 (Maddams et al. 2012).

###### Projected incidence in the UK

- It was estimated that the projected yearly average number of new cases of colorectal cancer will increase to 47,700 in 2038-2040 from 42,800 in 2023-2025 in the UK (Cancer Research UK 2023a).
- Projections for colorectal cancer incidence rates indicate 8% decrease between 2023-2025 and 2038-2040 to 57 cases per 100,000 persons yearly average by 2038-2040 (Cancer Research UK 2023a).

###### Projected prevalence in Wales

- It was estimated that there will be 27,000 (16,000 males and 11,000 females) people living with a colorectal cancer diagnosis in Wales by 2030 and this number is projected to increase to approximately 36,000 (22,000 males and 14,000 females) by 2040 (Macmillan Cancer Support 2020).

###### Risk factors

- Older persons are more likely to have colorectal cancer. In the UK, adults 75 and older account for more than 40 out of every 100 new cases of colorectal cancer (more than 40%) (Cancer Research UK 2018b).
- Regarding, exposure to risk factors, it is estimated that 54% of colorectal cancer cases in the UK are preventable (Cancer Research UK 2018a).
- Risk factors with ‘sufficient’ or ‘convincing’ evidence of increasing the risk of colorectal cancers include alcoholic drinks, tobacco smoking, X-radiation, gamma-radiation, processed meat, body fatness, and adult attained height (Cancer Research UK 2018a).
- There is also convincing evidence that physical activity decreases the risk of colon but not rectal cancer (Cancer Research UK 2018a).
- Other risk factors with ‘limited’ or ‘probable’ evidence of increasing the risk of colorectal cancers include Asbestos, Schistosoma japonicum, and red meat (Cancer Research UK 2018a).
- Likewise, the evidence that Wholegrains, dietary fibre in foods, dairy products, calcium supplements, and Oestrogen-progestogen contraceptives decrease the risk of colorectal cancers is ‘limited’ or ‘probable’ (Cancer Research UK 2018a).

##### Lung cancer

###### Current global incidence and prevalence

- It is estimated that around 2.2 million new lung cancer cases were diagnosed in 2020, contributing 11.4% of all cancers globally (Sung et al. 2021).
- Prevalence (5-year) was estimated to be around 2.6 million worldwide in 2020 (Globocan 2020).

###### Current incidence in the UK

- Based on Globocan (2020), the estimated number of new lung cancer cases in 2020 in the UK was 51,983.

###### Current incidence in Wales

- In 2019, 2,515 people were newly diagnosed with lung cancer (1,312 male and 1,203 female) (WCISU 2022).

###### Projected prevalence in the UK

- It was estimated that there will be approximately 274,000 males and 200,000 females living with a lung cancer diagnosis by 2030 and this this number is projected to increase to approximately 377,000 males and 255,000 females respectively by 2040 (Maddams et al. 2012).

###### Projected incidence in the UK

- It was estimated that the projected yearly average number of new cases of lung cancer will increase to 66,200 in 2038-2040 from 55,400 in 2023-2025 in the UK (Cancer Research UK 2023c).
- Projections for lung cancer incidence rates indicate 2% decrease between 2023-2025 and 2038-2040 to 78 cases per 100,000 persons yearly average by 2038-2040 (Cancer Research UK 2023c).

###### Projected prevalence in Wales

- It was estimated that there would be 7,000 (2,300 males and 4,410 females) living with a lung cancer diagnosis in Wales by 2030 and this number is projected to increase to approximately 9,000 (2,320 males and 6,610 females) by 2040 (Macmillan Cancer Support 2020).

###### Risk factors

- It is estimated that 79 percent of lung cancer cases in the UK may be avoided by reducing exposure to risk factors (Cancer Research UK 2018c).
- Risk factors with ‘sufficient’ or ‘convincing’ evidence of increasing lung cancer include production of graphite (Acheson process), production of aluminium, iron and steel, arsenic and inorganic arsenic compounds[a] asbestos (all forms), beryllium (and compounds), bis(chloromethyl)ether; chloromethyl methyl ether (technical grade), cadmium (and compounds), chromium (VI) compounds, household coal combustion emissions, production of coal/coke, coal-tar pitch, diesel engine exhaust hematite mining, MOPP (vincristine-prednisone-nitrogen mustard-procarbazine mixture), nickel compounds, outdoor air pollution (and particulate matter within), painting, plutonium, radon-222 and its decay products, production of rubber, crystalline silica dust, soot, sulphur mustard, tobacco smoking, environmental tobacco smoke, x-radiation, gamma-radiation, welding fumes, and beta-carotene supplements (high dose, in smokers) (Cancer Research UK 2018c).
- Other risk factors have ‘limited’ or ‘probable’ evidence of increasing lung cancer. This includes strong inorganic acid mists, production of art glass, glass containers and pressed ware, household biomass fuel (primarily wood) combustion emissions, working as a roofer (due to oxidized bitumen exposure), working with mastic asphalt (due to hard bitumen exposure) manufacture of carbon electrodes, alpha-chlorinated toluenes with benzoyl chloride, cobalt metal with tungsten carbide, creosotes, diazinon, fibrous silicon carbide, emissions from high-temperature frying, hydrazine, working as a non-arsenical insecticides sprayer, printing processes, and 2,3,7,8-Tetrachlorodibenzo-para-dioxin (Cancer Research UK 2018c). Fruits and dietary carotenoids have ‘limited’ or ‘probable’ evidence of decreasing lung cancer (Cancer Research UK 2018c).

##### Prostate cancer

###### Current global incidence and prevalence

- It is estimated that around 1.4 million new prostate cancer cases were diagnosed in 2020, contributing 7.3% of all cancers globally (Sung et al. 2021, Globocan 2020). Prevalence (5-year) was estimated to be around 5 million worldwide in 2020 (Globocan 2020).

###### Current incidence in the UK

- Based on Globocan (2020), the estimated number of new prostate cancer cases in 2020 in the UK was 56,780.

###### Current incidence in Wales

- In 2019, 2,971 people were newly diagnosed with prostate cancer (WCISU 2022).

###### Projected prevalence in the UK

- It was estimated that there will be approximately 620,000 males living with a prostate cancer diagnosis by 2030 and this number is projected to increase to approximately 831,000 by 2040 (Maddams et al. 2012).

###### Projected incidence in the UK

- It was estimated that the projected yearly average number of new cases of prostate cancer will increase to 85,100 in 2038-2040 from 62,500 in 2023-2025 in the UK (Cancer Research UK 2023e).
- Projections for prostate cancer incidence rates indicate 15% increase between 2023-2025 and 2038-2040 to 223 cases per 100,000 persons yearly average by 2038-2040 (Cancer Research UK 2023e).

###### Projected prevalence in Wales

- It was estimated that there would be 42,000 males living with a prostate cancer diagnosis in Wales by 2030 and this number is projected to increase to approximately 56,000 by 2040 (Macmillan Cancer Support 2020).

###### Risk factors

- Prostate cancer is not clearly linked to any modifiable risk factor (Cancer Research UK 2018d).
- Increasing age, black ethnicity, family history of prostate cancer, and genetics are all non-modifiable risk factors associated with prostate cancer (NICE 2022g).
- One of the main risk factors for prostate cancer is becoming older with people who are 50 years or older have an increased risk (NICE 2022g).
- Those of black ethnicity have a greater incidence and death rate from prostate cancer than men of white race although the reason for this is unclear. Compared to white men, who have a 1-in-8 lifetime chance of developing prostate cancer, black men have a 1-in-4 lifetime risk (NICE 2022g).
- A person’s risk increases if they have a close family who has had prostate cancer, such as a brother or father (NICE 2022g).
- Risk factors with ‘sufficient’ or ‘convincing’ evidence of increasing prostate cancer are currently not available (Cancer Research UK 2018d).
- Risk factors with ‘limited’ or probable evidence of increasing prostate cancer include androgenic (anabolic) steroids, arsenic and inorganic arsenic compounds, cadmium (and compounds), malathion, rubber production, thorium-232 and its decay products, x-radiation, gamma-radiation, red meat, body fatness (limited and probable evidence for fatal prostate cancer, and advanced prostate cancer respectively) and adult attained height (Cancer Research UK 2018d).

#### 2.2.3 Cardiovascular diseases

Cardiovascular disease is a general term for conditions affecting the heart or blood vessels including CHD, strokes and TIAs, and PVD (Raleigh et al. 2022). Collectively CVDs have remained the leading causes of death worldwide and substantially contribute to loss of health and excess health system costs (Vaduganathan et al. 2022).

##### Current global incidence

- The global incidence of CVD in 2017 was 72,721,167 people (British Heart Foundation 2020).
- An estimated 17.9 million people died from CVDs in 2019, representing 32% of all global deaths. Of these deaths, 85% were due to heart attack and stroke (World Health Organization 2021a).

##### Current incidence / prevalence in the UK

- The incidence of CVD in the UK in 2017 was estimated to be 1,498,059 people (British Heart Foundation 2020).
- The prevalence age standardised rate per 100,000 inhabitants of the UK for 2017 was 6,200 (British Heart Foundation 2020).
- It was estimated (2018/2019 data) that 7,6 million people are living with heart and circulatory diseases in the UK (British Heart Foundation 2020).

##### Current incidence / prevalence in Wales

- The incidence of CVD in the Wales in 2017 was estimated to be 76,542 people (British Heart Foundation 2020).
- The prevalence age standardised rate per 100,000 inhabitants of Wales for 2017 was 6,069 (British Heart Foundation 2020).
- It was estimated (2018/2019 data) that 340,00 people are living with heart and circulatory diseases in Wales (British Heart Foundation 2020).

##### Projected incidence in the UK

- There has been a substantial decline in CVD incidence in England and Wales over recent decades, however evidence from modelling studies estimates that this decline will plateau by 2029 (Collins et al. 2022).
- In people aged over 65 to 84 years, the numbers of people with CVD-related disability England and Wales will decline from 2015 to 2025 by 16.9% whereas for those aged 85 years or older this will increase by 6% (Guzman-Castillo et al. 2017).
- Cardiovascular-related disability is projected to reduce in England and Wales by 16.9% between 2015-2025 (Guzman-Castillo et al 2017).
- There may be an increase in event rates from MI in England for men before 2035 but increases for women are unlikely. Prevalence rates may fall in older men and are likely to remain stable in women over the next decade and a half (Scarborough et al. 2022).

##### Projected prevalence in Wales

- We did not find any modelling studies that estimated the projected prevalence of CVD in Wales.
- From the National dataset the percentage of adults over 18 years living in Wales who have any heart condition (excluding hypertension) is estimated to increase to 31% by 2035. However, this increase is estimated to be for those aged 18-54 and over 75 years of age and a decline is predicted for those aged 65-74 years (Public Health Wales Observatory 2018a).

##### Risk factors

- Cardiovascular disease which includes CHD, cerebrovascular disease, hypertension, and heart failure, is caused by non-modifiable and modifiable risk factors (Public Health Wales Observatory 2023).
- Non-modifiable risk factors include age, sex, ethnicity, and family history of CVD. Cardiovascular disease becomes increasingly common with advancing age. Men are at greater risk of disease than pre-menopausal women, but at post menopause a women’s risk is the same as a man’s risk (Public Health Wales Observatory 2023).
- In the UK material deprivation is a well-established risk factor for all CVDs and accounts for one fifth of the life expectancy gap between the most and least deprived communities in England (Raleigh et al. 2022).
- People from South Asian and Black groups are at highest risk of mortality from ischaemic heart disease, and in South Asian men and women it is 1.5 times more likely than that of the general population (Hollis et al. 2016; Raleigh et al. 2022).
- Chronic kidney disease has also been shown to be an independent risk factor for CHD development (Hollis et al 2016). Risk factors that can be controlled or modified include hypertension, raised cholesterol, diabetes, tobacco use, physical inactivity, unhealthy diet and obesity, and alcohol misuse (Public Health Wales Observatory 2023).
- Co-morbidities increasing the risk of CVD include hypertension, high cholesterol levels or dyslipidaemia, atrial fibrillation, diabetes, and chronic kidney disease (Raleigh et al 2022). Other risk factors include environmental risks such as ambient particulate matter air pollution, household air pollution from solid fuels, lead exposure, and low or high temperature (Vaduganthan et al 2022).

##### Coronary heart disease

###### Current global prevalence

- The global incidence of CHD in 2017 was 10, 636,538 people (British Heart Foundation 2020).

###### Current prevalence / incidence in the UK

- Data from the QOF prevalence for CHD (all ages) for 2021/2022 for England was 1,856,476 – which demonstrates a decreasing trend over time (Office for Health Improvement & Disparities 2022b).
- The incidence of CHD in the UK in 2017 was estimated to be 288,396 people (British Heart Foundation 2020).
- The prevalence age standardised rate per 100,000 inhabitants for 2017 was 1814 (British Heart Foundation 2020). It was estimated (2018/2019 data) that 2,272,226 (3.2%) people are living with CHD in the UK (British Heart Foundation 2020).

###### Current prevalence /incidence in Wales

- Incidence rates for CHD in 2017 for 100,000 of adult population aged 15+ was 254.2 (Men 335.5 and women 182.5) (British Heart Foundation 2020).
- The incidence of CHD in Wales in 2017 was estimated to be 10,584 people (British Heart Foundation 2020).
- The prevalence age standardised rate per 100,000 inhabitants for 2017 was 1,519 (British Heart Foundation 2020).
- It was estimated (2018/2019 data) that 117,733 (3.6%) people are living with CHD in Wales (British Heart Foundation 2020).

###### Projected prevalence in the UK

- It was estimated that the prevalence of CHD for those over the age of 65 in England by 2035 will be 2,172,500 people, which is a projected increase of 22.1% since 2015 (Kingston et al. 2018).

###### Projected prevalence in Wales

- We did not find any modelling studies that estimated the projected prevalence of CHD in the UK.

###### Risk factors

- See CVD narrative above.

##### Peripheral vascular disease

###### Current global prevalence

- It is estimated that >200 million people have PVD worldwide, with symptoms ranging from none to severe (Shu & Santulli 2018).
- Peripheral vascular disease is relatively uncommon among younger people; however, the prevalence of PVD rises with age and affects a substantial proportion of the elderly population (>20% in >80-year-old individuals (Shu & Santulli 2018).

###### Current prevalence / incidence in the UK

- Data from the QOF prevalence for PVD (all ages) for 2021/2022 for England was 355,295 – which demonstrates a decreasing trend over time (Office for Health Improvement & Disparities 2022h).
- This is supported by a UK cohort study which found a decreasing incidence and prevalence of symptomatic PVD diagnosed in primary care, suggested to have been influenced by secondary prevention strategies (NICE 2022e).
- It was estimated (2018/2019 data) that 361,833 (0.6%) people are living with PVD in England (British Heart Foundation 2020).

###### Current incidence in Wales

- Incidence rates for PVD in 2017 for 100,000 of adult population aged 15+ was 58.4 (Men 79.8 and women 38.5) (British Heart Foundation 2020).

###### Projected prevalence in the UK

- We did not find any modelling studies that estimated the projected prevalence of PVD in the UK.

###### Projected prevalence in Wales

- We did not find any modelling studies that estimated the projected prevalence of PVD in Wales.

###### Risk factors

- Smoking is one of the strongest risk factors for PVD and doubles the odds to PVD compared to non-smokers (NICE 2022f).
- Diabetes is strongly associated with an increased risk of PVD which increases the risk of developing PVD by 2-4 times (Shu & Santulli 2018).
- 90% of cases of PVD are accounted for by atherosclerosis and smokers have 2.5 times the risk of developing PVD (Shu & Santulli 2018).
- The likelihood of developing PVD is 50% greater among African Americans (Shu & Santulli 2018).
- Other risk factors for PVD include advanced age, hypertension, hypercholesterolaemia, known atherosclerotic disease elsewhere, chronic kidney disease and high serum homocysteine (NICE 2022f).
- The odds of developing PVD increase with higher numbers of risk factors with one risk factor leading to a 1.5 fold increase and three or more to a 10-fold increase (NICE 2022f).

##### Stroke or transient ischaemic attacks

###### Current global prevalence / incidence

- There are over 12.2 million new strokes each year worldwide and one in four people over age 25 will have a stroke in their lifetime (World Stroke Organisation 2022).
- Globally, there are over 110 million people currently living who have experienced stroke (World Stroke Organisation 2022).

###### Current prevalence / incidence in the UK

- Data from the QOF prevalence for stroke (all ages) for 2021/2022 for England was 1,117,509 which demonstrates an increasing trend over time (Office for Health Improvement & Disparities 2022j).
- The incidence of stroke in the UK in 2017 was estimated to be 107,372 people (British Heart Foundation 2020).
- The prevalence age standardised rate per 100,000 inhabitants of the UK for 2017 was 873 (British Heart Foundation 2020).
- It was estimated (2018/2019 data) that 1,300,423 (1.8%) people are living with stroke or TIA in the UK (British Heart Foundation 2020).

###### Current prevalence / incidence in Wales

- Incidence rates for stroke in 2017 for 100,000 of adult population aged 15+ was 300.0 (Men 328.3 and women 272.9) (British Heart Foundation 2020).
- The incidence of CHD in Wales in 2017 was estimated to be 6,266 people (British Heart Foundation 2020).
- The prevalence age standardised rate per 100,000 inhabitants of Wales for 2017 was 963 (British Heart Foundation 2020).
- It was estimated (2018/2019 data) that 68,870 (2.1%) people are living with stroke or TIA in the UK (British Heart Foundation 2020).

###### Projected prevalence in the UK

- With the projected aging population If the age-specific incidence remained stable over the next 30 years, the number of incident stroke would increase by 66% from year 2015 to 2045 (Li et al. 2020).
- With the projected aging population, if the age-specific stroke incidence continued to decline at a rate of 6% every 5 years, there would still be a 13% increase of the number of first-ever strokes in the UK by 2045 (Li et al. 2020).
- The prevalence of stroke survivors in the UK is expected to reach 2,119,400 by 2035, which is a projected increase of 123% since 2015 (King et al. 2020).
- The incidence (number of new strokes) in the UK is expected to reach 186,900 by 2035, which is a projected increase of 59% since 2015 (King et al. 2020).
- It was estimated that the prevalence of stroke for those over the age of 65 in England by 2035 will be 1,337,500 people, which is a projected increase of 84.2% since 2015 (Kingston et al. 2018).

###### Projected prevalence in Wales

- We did not find any modelling studies that estimated the projected prevalence of stroke in Wales.
- From the National dataset the percentage of adults over 18 years living in Wales who have had a stroke is estimated to increase to 33% by 2035 (Public Health Wales Observatory 2018a).

###### Risk factors

- Globally high systolic blood pressure is the largest single risk for stroke (World Stroke Organisation 2022).
- Risk factors for stroke and TIA include modifiable lifestyle factors such as smoking, alcohol misuse and drug abuse, physical inactivity, and poor diet (NICE 2022h).
- Established CVD including hypertension, atrial fibrillation (causes 20% of ischaemic strokes), infective endocarditis, valvular disease, carotid artery diseases, congestive heart failure, previous MI, congenital or structural heart disease are all risk factors (NICE 2022h).
- People with heart failure are 2-3 times more likely to have a stroke (British Heart Foundation 2023).
- People with diabetes are twice as likely to have a stroke as people without diabetes stroke (British Heart Foundation 2023).
- Several other medical conditions are also considered risk factors including migraine, diabetes, CKD, obstructive sleep apnoea (NICE 2022h).
- Other factors include older age, male sex, anticoagulation, previous or family history of stroke, lower level of education and genetic factors (NICE 2022h).

#### 2.2.4 Dementia

##### Current global prevalence

- It is estimated that around 55.2 million people worldwide have dementia and there are 10 million new cases each year (Alzheimer’s Research UK 2023).

##### Current prevalence in the UK

- In 2019 it was estimated that there are approximately 885,000 older people (aged 65 years and over) living with dementia in the UK (Wittenberg et al. 2019).

##### Current prevalence in Wales

- In 2019 it was estimated that that there are approximately 46,800 older people (aged 65 years and over) living with dementia in the Wales (Wittenberg et al. 2019).

##### Projected prevalence in the UK

- It was estimated that there will be approximately 1,233,400 people living with dementia in the UK by 2030 (Wittenberg. et al. 2019) and this number is projected to increase to approximately 1,590,100 people by 2040 (Wittenberg. et al. 2019).
- It was estimated that there will be 1,227,500 people aged over 65 years living with dementia In England by 2035, which is a projected increase of 86.1% increase since 2015 (Kingston et al. 2018).

##### Projected prevalence in Wales

- It was estimated that there would be 64,200 people living with dementia in the Wales by 2030 and this number is projected to increase to approximately 79,700 by 2040 (Wittenberg et al. 2019).
- This is a projected to rise by 70% from 2019 to 2040, due to improvements in life expectancy with the largest increases expected to occur in the oldest age brackets of 80 and above (Wittenberg et al. 2019).
- This represents a projected to increase from 7.0% to 9.0% in prevalence rate of dementia in the Wales (Wittenberg et al. 2019).
- It was estimated that there would be 6,821 people living with dementia in Wales aged between 65 and 74 years by 2025 and this is expected to further increase to 7,344 by 2030 and 7,853 by 2035 based on the Public Health Wales Observatory (2018a) dataset.
- For other age groups, projection trends vary (Public Health Wales Observatory 2018a). The projected number of people living with dementia between 75 and 79 years of age would be 9,346 by 2025 with a decrease expected by 2030 to 8,630. While numbers would increase to 9,203 by 2035 (Public Health Wales Observatory 2018a).
- Regarding age group 80 to 84, the projected number of people living with dementia would be 12,944 by 2025 with further increase expected by 2030 to 15,889. However, the number of people living with dementia in the 80-84 age group was projected to decrease to 14,832 by 2035 (Public Health Wales Observatory 2018a).
- For people over the age of 85, it was estimated that dementia prevalence would be 26,144 in 2025, with further increase expected in 2030 to 32,225 and in 2035 to 40,881 (Public Health Wales Observatory 2018a).
- Projected prevalence rates by local authorities in Wales for dementia are available from the <u>Social Care Wales Dataset.</u>

##### Risk factors

- Older age is the strongest risk factor for dementia (NICE 2022c) followed by genes (Alzheimer’s Society 2021).
- Other non-modifiable risk factors include mild cognitive impairment, learning disability, Apolipoprotein E, cardiovascular disease, cerebrovascular disease, and Parkinson’s disease (Alzheimer’s Society 2021, NICE 2022c).
- A third of instances of Alzheimer’s disease globally may be caused by modifiable risk actors (NICE 2022c).
- There are twelve risk factors that have been identified as having the potential to prevent or delay the onset of up to 40% of dementias and this include; lower educational attainment, hypertension, hearing impairment, smoking, obesity, depression, physical activity, diabetes, low social engagement and support, alcohol consumption, traumatic brain injury, and air pollution (Alzheimer’s Society 2021).
- Some long-term health conditions may create problems with a person’s thinking and memory that can evolve into dementia in severe instances. These conditions include: multiple sclerosis, HIV, rheumatoid arthritis, and kidney disease (Alzheimer’s Society 2021).

#### 2.2.5 Diabetes

##### Current global prevalence

- The global prevalence of diabetes in 20–79 year olds in 2021 was estimated to be 10.5% (536.6 million people), rising to 12.2% (783.2 million) in 2045 (Sun et al. 2022).
- Diabetes prevalence was similar in men and women and was highest in those aged 75–79 years and prevalence (in 2021) was estimated to be higher in urban (12.1%) than rural (8.3%) areas, and in high-income (11.1%) compared to low-income countries (5.5%) (Sun et al. 2022).

##### Current prevalence in the UK

- In 2019 it was estimated that that there are approximately 3,919,505 people diagnosed with diabetes in the UK (Diabetes UK 2023b).
- Diabetes QOF prevalence (17 Years Plus) for 2021/2022 was 3,625,401 which shows an increasing trend (Office for Health Improvement & Disparities 2022d).

##### Current prevalence in Wales

- In 2019 it was estimated that that there are approximately 198,883 people diagnosed with diabetes in Wales (Diabetes UK, 2023).
- Diabetes UK reports that Wales has the highest prevalence of diabetes in the UK and that in 2020 an additional 10,695 people were diagnosed with diabetes and that more than 209,015 people in Wales are living with diabetes.

##### Projected prevalence in the UK

- It was estimated that there will be approximately 362,960 people diagnosed with Type 1 diabetes, 3,936,720 with Type 2 diabetes (4,299,680 overall) by 2030 and this is a projected to rise by 24% and 7% respectively from 2013 to 2030 (Haider et al. 2021).
- The prevalence percentage of diabetes mellitus was projected to be 6.1% by 2030 which represents a 5.2% increase since 2014 (Ampofo & Boateng 2020).
- It was estimated that the prevalence of diabetes for those over the age of 65 in England by 2035 will be 3,115,400 people, which is a projected increase of 118.1% since 2015 (Kingston et al. 2018).

##### Projected prevalence in Wales

- We did not find any modelling studies that estimated the projected prevalence of diabetes in Wales.
- The percentage of adults over 18 years living in Wales with diabetes is estimated to increase to 21% by 2035 (Public Health Wales Observatory 2018a).
- On their website Diabetes UK reports that If current trends continue, 311,000 people in Wales could have diabetes by 2030 (Diabetes UK 2023a).

##### Risk factors

- Type 1 diabetes is the consequence of absolute insulin deficiency, usually due to an autoimmune process involving pancreatic beta cells. The main risk factors for Type 1 diabetes are genetic and environmental factors. As Type 1 diabetes is a heritable condition, having a sibling or parent with Type 1 diabetes increases the risk by up to 9% (NICE 2023a).
- For people with genetic predisposition to type 1 diabetes, autoimmunity to the pancreatic beta cells can be triggered by certain environmental factors. These environmental factors include diet, vitamin D exposure, obesity, being exposed to viruses associated with islet inflammation (such as enteroviruses) early in life, and decreased gut-microbiome diversity (NICE 2023a).
- Type 2 diabetes is estimated to make up 90% of total diabetes cases (Diabetes UK 2023c). Modifiable risk factors play a large role in the likelihood of someone suffering with Type 2 diabetes. Obesity makes up 80-85% of the total risk for Type 2 diabetes development, inactivity also contributes to this increased risk (NICE 2023b).
- A positive family history increases the risk of developing Type 2 diabetes by 2-6 fold cases (Diabetes UK 2023c).
- Age and ethnicity are risk factors for type 2 diabetes with those over 40 or other 25 and of African, Asian, and Afro-Caribbean descent. Those of African, Asian, and Afro-Caribbean descent being 2-4 times more likely to develop type 2 diabetes (cases (Diabetes UK 2023c).
- Other risk factors include: high blood pressure, high waist measurement, smoking, history of gestational diabetes, heart attack or stroke, polycystic ovary syndrome, mental health conditions (schizophrenia, bipolar and depression), alcohol, gestational diet history, low fibre, high glycaemic index diet (linked to obesity), drug treatments (statins, corticosteroids, antipsychotic medication), metabolic syndrome and having a low birth weight for gestational age (NICE 2023b).

#### 2.2.6 Heart failure

##### Current global prevalence

- Heart failure affects more than 64 million people worldwide (Savarese et al. 2023).

##### Current prevalence in the UK

- Data from the QOF prevalence for HF (all ages) for 2021/2022 for England was 586,237 which demonstrates an increasing trend over time (Office for Health Improvement & Disparities 2022e).
- The prevalence of heart failure slowly increases with age (until about 65 years) and then increases more rapidly (1 in 35 people aged 65-74 years, 1 in 15 people aged 75 to 85 years and 1 in 7 people aged 85 years and older) (NICE 2023e).
- It was estimated (2018/2019 data) that 658,944 (0.9%) people are living with HF in the UK (British Heart Foundation 2020).

##### Current prevalence / incidence in Wales

- Incidence rates for HF in 2017 for 100,000 of adult population aged 15+ was 158.1 (Men 187.8 and women 135.0) (British Heart Foundation 2020).
- It was estimated (2018/2019 data) that 34,453 (1.1%) people are living with HF in Wales and that this has increased slightly from 1.0% in 2006/2007 (British Heart Foundation 2020).

##### Projected prevalence in the UK

- We did not find any modelling studies that estimated the projected prevalence of HF in the UK.

##### Projected prevalence in Wales

- We did not find any modelling studies that estimated the projected prevalence of HF in Wales.

##### Risk factors

- Heart failure has been classified into three subtypes HF with reduced ejection fraction (HFrEF), HF with preserved ejection fraction (HFpEF) and HF mid-range ejection fraction (HFmrEF) with demographic and clinical characteristics differing between sub types (Savarese and Lund 2017).
- Savarese and Lund (2017) suggest that patients with HFpEF are more likely to be women and older, obese with other cardiovascular and non-cardiovascular comorbidities whilst the main determinant of HFrEF is coronary artery disease (coronary artery disease includes previous history of MI, hypertension, AF and diabetes).
- Coronary heart disease is the leading cause of HF in developed countries (Cowie et al. 2016).
- Other risk factors also include MI, congenital heart disease, age, hypertension, diabetes, body mass index (BMI), smoking, elevated low-density lipoprotein cholesterol, alcohol, sleep apnoea, AF, ethnicity, gender (Cowie et al. 2016).

#### 2.2.7 Hypertension

##### Current global prevalence

- The World Health Organisation (WHO) estimates that globally 1.28 billion adults have hypertension with the global age-standardized prevalence of hypertension in adults aged 30–79 years in 2019 was 32% in women and 34% in men (NICE 2023f).

##### Current prevalence in the UK

- It was estimated that around 1.8 million individuals were living with hypertension in 2017 which equates to 26.2% of the population (1 in 4 adults) (Public Health England 2020).
- It was estimated (2018/2019 data) that 9,930,115 (1.9%) people are on their GPs hypertension register in the UK (British Heart Foundation 2020).
- Data from the QOF prevalence for hypertension (all ages) for 2021/2022 for England was 8,604,825 which demonstrates an increasing trend over time.

##### Current prevalence in Wales

- It was estimated (2018/2019 data) that 512,542 (15.8%) people are on their GPs hypertension register in Wales (British Heart Foundation 2020).
- It is estimated that there are now around 700,00 adults in Wales who have high blood pressure (British Heart Foundation 2023).

##### Projected prevalence in the UK

- It was estimated that the prevalence of hypertension for those over the age of 65 in England by 2035 will be 6,423,400 people, which is a projected increase of 69.5% since 2015 (Kingston et al. 2018).

##### Projected prevalence in Wales

- We did not find any modelling studies that estimated the projected prevalence of hypertension in Wales.

##### Risk factors

- Similar to other cardiovascular disorders, risk factors for hypertension consist of modifiable and non-modifiable factors (NICE 2023g).
- Non modifiable risk factors include age (rises with advancing age), gender (up to 65 years women tend to have lower blood pressure than men, between 65-74 years women tend to have higher blood pressure), genetic factors (positive family history), and ethnicity, such as people of black African and black Caribbean origin are more at risk (NICE 2023g).

`o` There is mixed evidence regarding people of South Asian descent, although recent evidence suggests that men and women from this ethnic group have lower blood pressure than people from European descent (Pugliese et al. 2016).
`o` Men and women from Sub Saharan descent have been found to have higher blood pressure than people from European descent (Pugliese et al. 2016).
- Modifiable factors include lifestyle (smoking, excessive alcohol, unhealthy diet, high BMI (BMI>25), obesity, lack of physical activity), anxiety, stress, and depression (NICE 2023g, Pugliese et al. 2016).

`o` Regarding dietary issues, low sodium and increased potassium intake could reduce blood pressure (Pugliese et al. 2016).
`o` Other factors include social deprivation (in association with socio-economic quantile, education, and economic trajectory) and co-existing diabetes or kidney disease (NICE 2023g).
`o` Job strain has also been found to associate with higher blood pressure (Pugliese et al. 2016).
`o` In addition, air pollution is associated with high blood pressure, particularly in relation to long term exposure (Pugliese et al. 2016).

#### 2.2.8 Mental illness

##### Common mental disorders – depression and anxiety

Common mental disorders (CMDs) comprise different types of depression and anxiety which are usually less disabling than major psychiatric disorders and do not usually affect insight or cognition but can cause marked emotional distress and interfere with daily function. Symptoms of depression and anxiety frequently co-exist, with the result that many people meet criteria for more than one CMD. Their higher prevalence means the cumulative cost of CMDs to society is great (Stansfeld et al. 2016).

###### Current global prevalence

- Anxiety disorders are the most common psychiatric disorders (NICE 2023c).
- In 2019, 301 million people were living with an anxiety disorder including 58 million children and adolescents (World Health Organization 2022a).
- Generalized anxiety disorder (GAD) is more common in high income countries and has a lifetime prevalence of 1–7% in Europe (NICE 2023c).
- Depression is a leading cause of disability worldwide (NICE 2022d).
- The reported prevalence of depression varies widely depending on the country (both within and between countries), definitions of depression, methodology of the study and the population (NICE 2022d).
- The average lifetime prevalence estimate in high-income countries was 14.6% for adults (NICE 2022d).
- In 2019, 280 million people were living with depression, including 23 million children and adolescents (World Health Organization 2022a).

###### Current prevalence in the UK

- Data from the QOF prevalence for depression (18+ years) for 2021/2022 for England was 6,235,456 – which demonstrates an increasing trend over time (Office for Health Improvement & Disparities 2022c).
- Data estimates from Public Health England for 2021 estimated that 3,625,426 (9.2%) of the population have mixed anxiety and depressive disorder (Office for Health Improvement & Disparities 2022f).
- Data from the Adult Psychiatric Morbidity Survey (APMS) conducted in 2014 reported that 17.5% of working-age adults had symptoms of CMD. The prevalence of GAD increased from 4.7% in 2007 (and 4.4% in 1993) to 6.6% in 2014 and depression rose from 2.6% in 2007 (and 2.2% in 1993) to 3.8% in 2014 (Stansfeld et al. 2016).

###### Current prevalence in Wales

The latest data from the National Survey for Wales (2019/2020) reports that the prevalence of CMD in Wales for adults aged over 16 years was 16% and this translates to approximately 493,000 people.

###### Projected prevalence in the UK

- It was estimated that the prevalence of depression for those over the age of 65 in England by 2035 will be 191,600 people, which is a projected decline of 15.1% since 2015 (Kingston et al. 2018).

###### Projected prevalence in Wales

- We did not find any modelling studies that estimated the projected prevalence of CMDs in Wales
- However, projected prevalence rates by local authorities in Wales for CMDs are available from the <u>Social Care Wales Dataset</u>.

###### Risk factors

- There are several risk factors for depression which include: female sex, older age, previous or family history of depression or other mental health conditions or substance misuse, personal, social or environmental factors such as stress, poverty, unemployment, social isolation, child maltreatment, chronic physical health conditions such as diabetes, chronic obstructive pulmonary disease, CVD, chronic pain, epilepsy and stroke (NICE 2022d).
- Environmental factors include childhood adversity (maltreatment, neglect, maternal depression, family disruption, domestic violence, parental alcoholism or drug use, parental mental health problems, overprotective or overly harsh parenting style, bullying or victimization among peers), history of physical, sexual or emotional trauma (physical or sexual abuse, motor vehicle accident, sudden bereavement), and sociodemographic factors (separated, widowed or divorced, unemployment, low socioeconomic status or education levels) (NICE 2023d).
- Anxiety disorders include GAD, panic disorder, phobias, and obsessive-compulsive disorder. Since 2007 CMDs have increased in midlife men and women (aged 55 to 64) and approached significance in young women (aged 16 to 24). They are more common in certain groups of the population including Black women, adults under the age of 60 who lived alone, women who lived in large households, adults not in employment, those in receipt of benefits and those who smoked cigarettes (Stansfield at al. 2014).
- Multiple factors contribute the development of GAD, with both genetic and environmental causes identified (NICE 2023d).
- Family history of anxiety disorders, depression or other psychiatric disorders can lead to increased risk of developing GAD. People with comorbid anxiety disorder, such as panic disorder, or social phobia, are more likely to experience GAD. Female sex is also associated with GAD, as women are twice as likely to experience GAD than men. In addition, people with chronic physical conditions, such as CVD, cancer, respiratory disease, diabetes, or arthritis, are more likely to experience GAD (NICE 2023d).

##### Bipolar disorder

###### Current global prevalence

- Globally, bipolar disorder is ranked as the 18^th^ most common health condition in years lived with disability (NICE 2022a).
- In 2019, 40 million people experienced bipolar disorder (World Health Organization 2022a).
- Bipolar I disorder might be slightly more prevalent than bipolar II disorder (NICE 2022a).
- The World Mental Health Survey Initiative reported that the lifetime aggregate prevalence from 11 countries (not UK) was 2.4% for bipolar disorders (Merikangas et al. 2011).

###### Current prevalence in the UK

- Data from the APMS reported that Approximately 2% of the UK adult population screened positive for bipolar disorder in 2014 with similar rates between men and women (Marwaha et al. 2016).
- Younger age groups were more affected with 3.4% of young people aged 16-24 years diagnosed with bipolar disorder (NICE 2022a). In comparison, bipolar disorder affected 0.4% of 65-74-year-olds (NICE 2022a).

###### Current prevalence in Wales

- We were unable to identify any recent National survey data for Wales specifically for bipolar disorder.

###### Projected prevalence in the UK

- We did not find any modelling studies that estimated the projected prevalence of bipolar disorders in the UK.

###### Projected prevalence in Wales

- We did not find any modelling studies that estimated the projected prevalence of bipolar disorders in Wales.
- However, projected prevalence rates by local authorities in Wales for bipolar disorders are available from the <u>Social Care Wales Dataset</u>.

###### Risk factors

- The interplay of genetical and environmental factors can contribute to the development of bipolar disorder (NICE 2022b).
- Among psychiatric disorders, bipolar disorder is considered the most heritable condition (NICE 2022b).
- People with a first-degree relative diagnosed with bipolar disorder have a lifetime risk of the condition which is approximately five times greater than that of the general population (NICE 2022b).
- Environmental factors that could lead to development or relapse of bipolar disorder include early life stress, maternal death before the age of five, childhood trauma or abuse, emotional neglect, Toxoplasma gondii exposure, cannabis/cocaine use or exposure (NICE 2022b).
- The likelihood of screening positive for bipolar disorder varied with employment status with unemployed or economically inactive 16–64-year-olds more likely to screen positive than those in employment (Marwaha et al. 2016).
- Bipolar disorder is comorbid with other disorders including substance misuse, anxiety disorders, personality disorders and attention-deficit/ hyperactivity disorder ((Marwaha et al. 2016).

##### Psychotic disorders -psychosis and schizophrenia

Psychotic disorders are a group of mental health conditions that are characterized by a range of symptoms, including hallucinations, delusions, disorganized thinking, and abnormal behaviour that are severe enough to distort perception of reality. The main types of psychotic disorders are schizophrenia and affective psychosis (Bebbington et al. 2016).

###### Current global prevalence

- In 2019 it was estimated that schizophrenia affects approximately 24 million people or 1 in 300 people worldwide (World Health Organization 2022a).

###### Current prevalence in the UK

- Data from the APMS in 2014 reported that the overall, the prevalence of ‘psychotic disorder in the past year’ has remained broadly stable at less than one adult in a hundred (0.4% in 2007, 0.7% in 2014). In both men and women, the highest prevalence was observed among those aged 35 to 44 years (1.0% and 0.9% respectively) (Bebbington et al. 2016).

###### Current prevalence in Wales

- The latest data from the <u>National Survey for Wales (2019/2020)</u> reports that the prevalence of psychotic disorders in Wales for adults aged over 16 years was 16%.

###### Projected incidence in the UK

- For 2025 it was predicted that there would be a treated caseload of 11,067 and a probable first episode psychosis case-load of 9,541 or those aged 16–64 years (McDonald et al. 2021).
- Between 2019 and 2025 it was predicted that treated caseloads will increase by 6.2%; but it was also acknowledged that there is also a possibility of a small decline (McDonald et al. 2021).

###### Projected prevalence in Wales

- We did not find any modelling studies that estimated the projected prevalence of psychotic disorders in Wales.
- However, projected prevalence rates by local authorities in Wales for psychotic disorders are available from the <u>Social Care Wales Dataset.</u>

###### Risk factors

- Rates of psychotic disorder have been found to be higher in men from ethnic groups and higher in black men (3.2%) than men from other ethnic groups. In the 2014 APMS survey It was more common in people who live alone, socioeconomic factors were strongly linked with psychotic disorder (Bebbington et al 2016).
- Interactions between several genetic, social and environmental risk factors appear to be involved, including heritability, family heritage, stressful life events, migration, urban living, cannabis use, other substance use, medication use, early life factors, parental age (>40 years and <under 20 years) and exposure to the protozoan parasite **-** *Toxoplasma gondii* (NICE 2021).

#### 2.2.9 Multi-morbidities

The term “multimorbidity” refers to the coexistence of two or more chronic conditions in the same individual (World Health Organization 2016). As a consequence of an ageing population and an increase in long term conditions, the number of people with multiple conditions is set to rise, impacting safety in primary care (World Health Organization 2016).

The true extent of multimorbidity is acknowledged as being difficult to gauge (Academy of Medical Science, 2018; Stafford et al, 2018) There is no agreed definition or classification system for reporting, resulting in an evidence base that is often fragmented and difficult to interpret, limiting any conclusions regarding both scale and impact (The Academy of Medical Sciences 2018). There is also no agreement on which, or how many conditions should be considered leading to widely differing estimates of prevalence (Stafford et al. 2018).

##### Current global prevalence

- A report by the Academy of Medical Science (2018) notes that multimorbidity is considered a norm and not an exception within primary healthcare services of most high-income countries and is prevalence appears to be increasing.
- A European study, from 2013, estimated that 50 million people in the European Union were living with multiple chronic diseases (Rijken et al. 2023).

##### Current prevalence in the UK

- The percentage prevalence of multimorbidity across the UK is between 23.2% (Scotland - (Barnett et al. 2012) and 27.5% (England - (Cassell et al. 2018)) and is higher for females and increases significantly with age (Cassell et al. 2018).
- One in three patients admitted to hospital as an emergency has five or more health conditions, up from one in ten a decade ago (Department of Health and Social Care 2021).
- Although having multiple conditions is often thought of as being related to old age, 30% of people with 4+ conditions are under 65 years of age, and this percentage is higher in disadvantaged areas (Stafford et al. 2018).

##### Current prevalence in Wales

- We did not find any data on the prevalence of multimorbidity in Wales.

##### Projected prevalence in the UK

- It was estimated that the prevalence of two or more multi-morbidities in England by 2035 will be 9,789,100 people aged over 65 years and 2,548,800 people aged over 85, which is a projected increase of 86.4%and 181.6% respectively since 2015.
- It was estimated that the prevalence of four or more multi-morbidities in England by 2035 will be 2,453,200 people aged over 65 years and 1,117,500 people aged over 85, which is a projected increase of 157.6% and 470.2% respectively since 2015.

##### Projected prevalence in Wales

- We did not find any modelling studies that estimated the projected prevalence of psychotic disorders in Wales.

##### Risk factors

- Most epidemiological research has focused on the contribution of discrete factors (e.g., smoking, blood pressure, diabetes) to the risk of developing specific conditions (e.g., coronary heart disease, stroke) however it is recognised that individuals are not exposed to single risk factors in isolation. Instead, patients typically experience multiple risk factors in different and dynamic combinations often resulting in multiple conditions at different time points (The Academy of Medical Sciences 2018).
- It is not fully understood how the many biological, psychological, behavioural, socioeconomic, and environmental risk factors act in the genesis of multimorbidity due to the complexity (The Academy of Medical Sciences 2018).
- Future efforts to understand how risk factors interact to produce specific clusters of conditions are required, both in the general population and in specified population subgroups (The Academy of Medical Sciences 2018).

#### 2.2.10 Obesity

##### Current global prevalence

- The worldwide prevalence of obesity has almost tripled between 1997 and 2016 (World Health Organization 2021b).
- In 2016 it was estimated that more than 1.9 billion adults aged 18 years and older were overweight and of these over 650 million adults were obese (World Health Organization 2021b).
- In 2016 approximately 13% of the world’s adult population (11% of men and 15% of women) were obese (World Health Organization 2021b).

##### Current prevalence in the UK

- Data from the QOF prevalence for obesity (18+ years) for 2021/2022 for England was 4,791,038 – which demonstrates a decreasing trend over time (Office for Health Improvement & Disparities 2022g).
- Between 1993 and 2011, obesity rates rose from 13% to 24% in men and from 16% to 26% in women (NICE 2023h).

##### Current prevalence in Wales

- In 2020 it was reported that 61% of adults in Wales were classified as overweight or obese; including 36% who were overweight, 22% who were obese and 3% who were morbidly obese (Hannah et al. 2021).

##### Projected prevalence in the UK

- It is estimated that the projected prevalence of obesity will continue to rise in the England (Bhimjiyani et al. 2016a; Cobiac and Scarborough 2021).
- Obesity in the UK is expected to reach maximum levels between 2032 and 2038 among men, and between 2031and 2037 among women (Janssen et al. 2020). In 2060, obesity prevalence is expected to decline (Janssen et al. 2020).

##### Projected prevalence in Wales

- It is estimated that the percentage of adults who are overweight or obese in Wales will increase to approximately 62.2% by 2025 (Public Health Wales Observatory 2018a).
- Obesity estimates vary across studies however an increase in obesity in Wales is projected by 2025 and 2035.

##### Risk factors

- Obesity can be influenced by lifestyle factors such as food and drink consumption, physical inactivity, social and psychological factors such as low self-esteem or depression, obesity-related genes including the gene which controls the production of leptin, medical conditions and certain medications (NICE 2023i).
- Other risk factors include age, menopause, prior pregnancy, sleep deprivation, less formal education and low socioeconomic status (NICE 2023i).

#### 2.2.11 Smoking

##### Current global prevalence

- In 2020, an estimated 22.3% of the global population aged 15 years and older were current users of some form of tobacco, down from approximately one third (32.7%) in 2000 (World Health Organization 2022c).
- In 2000, about one half of men (49.3%) and one in six women (16.2%) aged 15 years and older were current users of some form of tobacco. By 2020, the proportion of men using tobacco had declined to slightly over one in three (36.7%), while that of women had declined to one in thirteen (7.8%) (World Health Organization 2022c).
- As a result of COVID-19, very little prevalence data is available from surveys done in 2020 and 2021 therefore the impact of the pandemic on National and global prevalence rates of tobacco use is not yet known (World Health Organization 2022c).

##### Current prevalence in the UK

- Data from the QOF prevalence for smoking (15+ years) for 2021/2022 for England was 7,803,228 – which demonstrates a decreasing trend over time (Office for Health Improvement & Disparities 2022i).
- In the UK, in 2021, 13.3% of people aged 18 years and over smoked cigarettes, which equates to around 6.6 million people in the population; this is the lowest proportion of current smokers since records started in 2011 based on our estimates from the Annual Population Survey (Office for National Statistics 2022).
- In 2021, 15.1% of men smoked compared with 11.5% of women in the UK; this trend has been consistent since 2011 (Office for National Statistics 2022).

##### Current prevalence in Wales

- The percentage of adults reporting to be a current smoker, aged 18+ in 2016 was 18.1% (males) and 15.8% (females) (Public Health Wales Observatory 2017a).
- In Wales in 2021 14.1% of people aged 18 years and over smoked cigarettes.

##### Projected prevalence in the UK

- Smoking prevalence in England is projected to continue to decline. The decline is projected to be faster in relative terms among more advantaged groups; therefore, relative educational inequalities in smoking prevalence are projected to increase (Pérez-Ferrer et al. 2018).

##### Projected prevalence in Wales

- It is estimated that smoking prevalence of all persons aged over 16 years in Wales will decline to 13.3% by 2039 from 2016 assuming no change in the numbers of people starting or quitting smoking (Public Health Wales Observatory 2018a).

##### Smoking related diseases

- Smoking is one of the leading risk factors driving the UK’s high burden of preventable ill health and premature mortality. All are socioeconomically patterned and contribute significantly to widening health inequalities (Everest et al. 2022).
- According to the World Health Organisation, tobacco kills more than 8 million people a year worldwide, including around 1.2 million deaths from exposure to second hand smoke, and is one of the biggest public health threats (World Health Organization 2022b).
- It is reported that there are over 7000 chemical components in tobacco smoke, of these, over 250 are toxic or carcinogenic, such as aldehydes, nitrides, and others that irritate the respiratory tract. Smoking can damage nearly all organs of the human body and is one of the main risk factors for respiratory infection and infectious diseases (Jiang et al. 2020).
- To reduce exposure to risk factors and tackle inequalities, governments will need to deploy multiple approaches that address the complex system of influences shaping people’s behaviour (Everest et al. 2022).
- Active smoking cessation promotion and education are practical and cost-effective measures to prevent the risk of infection caused by smoking and to reduce the incidence rate and mortality of infection (Jiang et al. 2020).

#### 2.2.12 Bottom line summary

The extent of the available evidence for each condition is summarised in the form of a graphical map (See Figure 1), populated by information describing the number and type of projected prevalence or incidence data (organisational data, National datasets, or modelling studies) with the conditions paired alongside the country (Wales, England and UK). If a condition was described as having projected prevalence or incidence data this was indicated on the evidence map using the following key: projected prevalence from organisational data (OP), projected prevalence from a National data set (NP), projected incidence from a National data set (PI), projected prevalence from a modelling study (MP), and projected incidence from a modelling study (MI). We did not identify any Welsh modelled prevalence or incidence data for AF, HF, hypertension, or multi-morbidities.

## 3. DISCUSSION

### 3.1 Summary of the findings

The projected prevalence and/or incidence rates for all long-term conditions listed in Table 1, across the UK and considering all-ages rates, indicated that AF, cancer, CVD, dementia, diabetes, hypertension, multi-morbidities, psychotic disorders, stroke, will increase and for some long-term conditions this is anticipated to be in addition to the impacts of demographics. Conversely, the prevalence of depression among older people is expected to decline.

We did not find any evidence (modelling studies or grey literature reports) for the projected prevalence or incidence of HF. However, Savarese and Lund (2017) predicted that globally the prevalence of HF will steadily increase. For PVD, QOF prevalence data has demonstrated a decreasing trend over time as of 2021 (Office for Health Improvement & Disparities 2022h) but we did not find any evidence for projected prevalence or incidence.

Although the majority of the evidence reports a decline in the incidence of CVD in England and Wales, Collins et al. 2022 reported that this decline will plateau by 2029. The BHF reports that the annual number of heart and circulatory deaths in Wales has fallen by half since it was established and that it is anticipated that despite an ageing and growing population improved survival rates from heart and circulatory events could see the numbers of people living with CVD continue to decline (British Heart Foundation 2023). However, it is anticipated that CVD prevalence rates will rise among individuals aged over 85 years, while decreasing among younger age groups (Guzman-Castillo et al. 2017). Only one modelling study was identified for projecting prevalence of hypertension (high blood pressure) however, this only included English data which estimated a 69.5% increase from 2015 to 2035. We did not find any modelling studies or grey literature reports that estimated the projected prevalence of hypertension in Wales. Over time, there has been a notable rise in the prevalence of stroke. Projections indicate that by 2035, the percentage of Welsh adults who have experienced a stroke will reach 33%, representing a significant increase. Modifiable lifestyle factors, such as smoking, alcohol misuse, drug abuse, physical inactivity, and poor diet, contribute to the risk of stroke and transient ischemic attacks (NICE 2022h). We found two studies that provided estimates for the projected prevalence or incidence of AF and AF-related embolic vascular events in the UK. Extrapolating this data to the population of Wales based on ONS data for those aged 80+ estimated that this would double by 2050 (Welsh Government 2023).

If current trends continue the number of people in Wales living with cancer will rise, with projected prevalence expected to reach just around 300,000 by 2040 (Macmillan Cancer Support 2020). Screening and early diagnosis or early detection of risks is also important, and genetic testing could help identify people at risks of developing certain conditions, such as breast cancer (Pashayan et al. 2020). With timely risk stratification, preventative measures can often be taken, and for example in women with genetic predisposition to breast cancer, measures such as chemo- or surgical prevention could help reduce the chances of developing the disease (Pashayan et al. 2020).

By 2040, it is projected that the prevalence of dementia among the population of Wales will increase. This is primarily attributed to the expanding elderly demographic and extended life expectancy, with the most significant growth anticipated among individuals aged 80 years and older. The most significant risk factor for dementia is advancing age (NICE 2022c), closely followed by genetic factors (Alzheimer’s Society 2021). Additionally, hypertension, smoking, obesity, diabetes, and alcohol consumption are among the other risk factors associated with dementia (Alzheimer’s Society 2021). By making efforts to reduce these risk factors, it is possible to prevent or delay the onset of dementia by up to 40% (Alzheimer’s Society 2021).

We reported that the prevalence of depression is projected to decline by 15.1% by 2035 in England for over 65 year-olds (Kingston et al. 2018). However, we did not find projected prevalence data for depression, anxiety, psychotic or bipolar disorders for Wales from modelling studies or the grey literature. Frequently, symptoms of depression and anxiety co-occur, leading to a significant number of individuals meeting the criteria for multiple common mental disorders (Stansfeld et al. 2016). Depression and anxiety are influenced by various risk factors, encompassing social demographic factors, age, sex, ethnicity, health conditions, socio-economic status, and environmental factors (such as childhood adversity) (NICE 2023d). While the findings of this REM indicate that the prevalence of depression will decline by 2035, this only includes people over the age of 65 (Kingston et al. 2018), which would not consider the working population in the UK. More research will be needed into how common mental disorders affect the working age population. Further findings from this REM predicted that there will be a 6.2% increase (2019 to 2025) in the caseloads of those treated for psychotic disorders, but the authors of the modelling study that reported this data also acknowledged that there is also a possibility of a small decline (McDonald et al. 2021). The risk factors for bipolar disorder, psychosis and schizophrenia are similar but also include genes, substance misuse, trauma, and *Toxoplasma gondii* exposure (NICE 2022b).

According to the UK Governments’ Health and Care White Paper (2021), the proportion of patients admitted to hospitals as emergency cases with five or more health conditions has increased from one in ten a decade ago to one in three. We did not find any projected prevalence data for Wales but data for England shows that the proportion of the population with four of more diseases is projected to double by 2035 (Kingston et al. 2018).

The prevalence of diabetes is projected to increase, and this is partially influenced by the aging population. Obesity and poor diet are the primary risk factors for type 2 diabetes (NICE 2023b). If these factors continue to escalate, the incidence of diabetes will rise at a faster rate than what can be attributed to demographic growth alone (NICE 2023b). Obesity is also a risk factor for other long-term conditions including dementia and hypertension. Nine modelling studies were identified that provided estimates for the projected prevalence of obesity and/ or morbid obesity with three of these providing projections for Wales. From these nine studies we found that although estimates for the future prevalence of obesity vary, an increase in obesity in Wales is projected by 2025 and 2035. In the UK, it is anticipated that obesity levels will reach their peak between 2032 and 2038 for men, and between 2031 and 2037 for women. However, by the year 2060, obesity prevalence is expected to decline (Janssen et al. 2020). Obesity can be influenced by various lifestyle factors, including the consumption of food and drinks, physical inactivity, as well as social and psychological factors such as low self-esteem or depression (NICE 2023i). Additionally, genetic factors, including the gene responsible for regulating leptin production, medical conditions, and certain medications, can also contribute to obesity (NICE 2023i).

Over time, adult smoking rates in Wales have shown a consistent decline. However, it remains a significant cause of ill health despite this decreasing trend. Smoking rates are projected to continue to decline across all age groups in Wales and are predicted to reach 13.3% by 2039 assuming no change in the numbers of people starting or quitting smoking (Public Health Wales Observatory 2018b). A more recent analysis of data from the National Survey for Wales has found that due to a declining trend, the current smoking prevalence in Wales is lower than previous projections and updated projections suggest that smoking prevalence in Wales could be less than 5% by late 2030. Smoking data, however, should be treated with caution as self-reported smoking rates are often slightly lower than the actual number due to a lack of truthfulness in people’s responses when surveyed. Smoking is a well-established risk factor for numerous chronic conditions, including various types of cancers, dementia, and cardiovascular diseases (CVDs). Other modelling studies have provided valuable insights into the potential impact of policy changes on smoking rates in relation to the burden of smoking-related conditions (Allen et al. 2016, Hunt et al. 2018, Knuchel-Takano et al. 2018, Kypridemos et al. 2017). By implementing effective policies aimed at tobacco control, such as increasing taxation on tobacco products, implementing smoking bans in public places, and launching public health campaigns, it is possible to encourage a decline in smoking rates and subsequently lower the burden of smoking-related diseases.

By focusing on long-term conditions where the risk factors are modifiable through individual lifestyle changes and public health interventions, the projected trajectory of prevalence could be mitigated; this includes CHD, AF, stroke, some cancers, and type 2 diabetes. By encouraging positive lifestyle changes, such as adopting healthy eating habits, engaging in regular physical activity, and avoiding harmful behaviours like smoking and excessive alcohol consumption, individuals can significantly reduce their risk of developing certain disease.

### 3.2 Limitations of the available evidence

We did not perform a quality appraisal for the studies included in this REM, and therefore, it is important to exercise caution when formulating conclusions based on the available evidence.

### 3.3 Summary of the Evidence gaps

On conducting the REM, we were unable to identify any modelled prevalence or incidence data specific to Wales for conditions such as AF, HF, hypertension, or multi-morbidities. Which highlights a gap in the available data pertaining to the prevalence and incidence of these specific health issues in the context of Wales.

### 3.4 Strengths and limitations of this Rapid Evidence Map

A strength of this REM is that the literature searches were performed by both an information specialist and a skilled systematic review methodologist. The searches were conducted across multiple databases and employed various combinations of search terms to ensure a comprehensive coverage of chronic and long term conditions and risk factors. The initial search for evidence on poor diet and nutrition yielded no results, prompting the research team to conduct an additional search using the term “suboptimal nutrition.” However, this subsequent search also failed to retrieve any relevant findings. Future work in this field should consider identifying the dietary risks that are projected to have the most significant impact on disease burden in Wales, such as whole grains, fruits, legumes, and so on. Subsequent searches could then be conducted specifically targeting these identified dietary factors.

In addition, we conducted searches for grey literature sources, including national datasets, to gather projected prevalence and incidence data, enabling the identification of knowledge gaps. However, due to the rapid nature of REMs and certain streamlined processes, such as study selection being conducted by one researcher, it is possible that available modelling studies were missed. Furthermore, due to the broad scope of chronic and long term conditions and risk factors, this REM had to be limited to certain topics and we did not search for evidence of asthma, chronic kidney disease, chronic obstructive pulmonary disease, epilepsy, rheumatoid arthritis, influenza, COVID-19, long Covid, other infectious diseases and new conditions identified by genomic testing. Thus, the availability of projected prevalence or incidence data and subsequent research gaps in these areas were not identified.

### 3.5 Implications and next steps

- Further research and data collection initiatives are necessary to address knowledge gaps, particularly for conditions, such as AF, HF, hypertension, and multi-morbidities and to provide a more comprehensive understanding of the burden of these conditions in the Welsh population.
- Increases are expected in the incidence and prevalence of most conditions included in this rapid evidence map, thus interventions, such as early diagnosis and prevention, should be brought to the forefront of healthcare to mitigate this rising prevalence of several preventable long term conditions.
- Lifestyle change and certain public health interventions could help prevent the development of some long-term conditions, such as CHD, AF, stroke, type 2 diabetes and some cancers. A shift in NHS focus to prevention through behavioural science approaches and increased investment into such interventions should be considered by policy makers and healthcare leaders.
- Smoking, excessive drinking and obesity are particular candidates for targeted preventive work, especially in areas of deprivation, to lessen health inequalities.

#### 3.6 Economic considerations*

Mental Health problems cost the Welsh economy £4.8 billion per annum. 72% of these incurred costs are attributed to productivity losses of people living with mental health conditions and costs experienced by unpaid informal carers (McDaid et al., 2022).

**CVD costs the Welsh economy £800 million per annum. 33% of these incurred costs are attributed to healthcare costs [£260 million], 20% to informal care costs [£150 million] and 6% to social care costs [£50 million] (Collins et al., 2022).

**Dementia costs the Welsh economy £700 million per annum. 35% of these incurred costs are attributed to social care costs [£260 million], 24% to informal care costs [£180 million] and 12% to healthcare costs [£90 million] (Collins et al., 2022).

Increased levels of deprivation are associated with increased risks of multimorbidity (Pathirana & Jackson, 2018). Individuals living with multimorbidity have higher total healthcare costs, including higher hospital and care transition costs (Soley-Bori et al., 2021).

People living in the most deprived areas in Wales face considerably reduced healthy life expectancy when compared to the least deprived areas. The difference in healthy life expectancy between those living in the least and most deprived areas of Wales is 18 years for women. The difference is 17 years in men (The Health Foundation, 2022). Living in the most deprived areas in Wales is also strongly correlated with reduced labour market participation (Murray et al., 2022)

\**This section has been completed by the Centre for Health Economics & Medicines Evaluation (CHEME), Bangor University*

*\*\*These figures have been calculated pro rata using <u>ONS 2021 population figures for</u> <u>England and Wales.</u> With Wales representing 5.2% of the population of England and Wales as a combined entity.*

## Abbreviations

Acronym: Full Description
APMS: Adult Psychiatric Morbidity Survey
AF: Atrial fibrillation
BHF: British Heart Foundation
BMI: Body mass index
CFAS: Cognitive Function and Ageing Studies
CHD: Coronary heart disease
CMDs: Common mental disorders
COPD: Chronic obstructive pulmonary disease
CVD: Cardiovascular disease
ELSA: English Longitudinal Study of Ageing
GAD: Generalized anxiety disorder
GBD: Global burden of disease
HES: Hospital episode statistics
HF: Heart failure
HFmrEF HF: mid-range ejection fraction
HFrEF HF: with reduced ejection fraction
HRT: Hormone replacement therapy
HSE: Health Survey for England
MI: Myocardial infarction
MODEM: Modelling Outcome and Cost Impacts of Interventions for Dementia study
ONS: Office for National Statistics
PVD: Peripheral vascular disease
QAIF: Quality Assurance and Improvement Framework
QOF: Quality Outcomes Framework
TIA: Transient ischaemic attacks
UI: Uncertainty interval
WCISU: Welsh Cancer Intelligence and Surveillance Unit
WHO: World Health Organisation

## 5. RAPID EVIDENCE MAP METHODS

### 5.1 Eligibility criteria

The CoCoPop framework has been used to inform the eligibility criteria: **C**onditions and **C**ontext, **P**opulation (Munn et al. 2020)

### 5.2 Literature search

#### 5.2.1 Evidence sources

Comprehensive searches have been conducted across four or five databases for English language publications from 2012 to March 2023 according to the condition.

- On the Ovid Platform: Medline, Embase, Emcare, PsycINFO
- On the Ebsco Platform: Cumulative Index of Nursing
- SCOPUS

The websites of key third sector and government organisations relevant to each condition will also be searched.

#### 5.2.1 Search Strategy

An initial search of MEDLINE has been undertaken using the following terms: trend* or future or projected or projecti* or predicti* or forecast* or model adj prevalence or incidence

AND
UK or “United Kingdom” or Wales or England or Scotland or Ireland
AND
Dementia or Alzheimer*

This informed the development of a search strategy which was tailored for each information source for each condition. A full search strategy for Medline is detailed in Appendix B. The reference list of all included publications screened for additional studies. One deviation from the protocol is that due to time constraints we were not able to conduct forward citation tracking.

#### 5.2.2 Reference management

All citations retrieved from the database searches were imported or entered manually into EndNote^TM^ (Thomson Reuters, CA, USA) and duplicates removed. At the end of this process the citations that remain were imported to Rayyan^TM^ and any further duplicates removed.

### 5.3 Study selection process

One reviewer screened all the citations using the information provided in the title and abstract using the Rayyan^TM^, and a second reviewer screened 10% of the citations and any disagreements were resolved by discussion. For citations that appeared to meet the inclusion criteria, or in cases in which a definite decision could not be made based on the title and/or abstract alone, the full texts of all citations were retrieved. The full texts have been screened for inclusion by one reviewer using a purposefully developed screening tool and all decisions were checked by a second reviewer, and any disagreements were resolved by discussion. The flow of citations through each stage of the review process for each condition has been tabulated.

### 5.4 Data extraction and coding/charting

Data extraction and coding was performed on included full-text studies by one reviewer and checked by another using a data extraction tool. The data extraction tool includes items related to study characteristics (citation, year data collected, country, condition, study design, source of data) and in addition data on projected prevalence/incidence (by age and gender where available). A coding strategy was followed in order to ensure the consistency in data extraction. The data extraction tool was first piloted between the reviewers in a random sample of 4 included studies, in order to ensure consistency in extraction and any discrepancies were resolved either by consensus.

### 5.5 Assessment of methodological quality

Due to the substantial amount of evidence obtained and the timeframe for this rapid evidence map (REM), we were unable to perform a quality appraisal, thus deviating from the protocol.

### 5.6 Data summary

The data has been tabulated and reported narratively as a series of thematic summaries across each condition (Thomas et al 2017).

## 6. EVIDENCE

### 6.1 Study selection flow chart

### 6.2 Data extraction tables

### 6.3 Information available on request

The protocol is available on request, and the search strategies and a list of excluded studies for each condition are available in the additional information.

## Data Availability

All data produced in the present study are available upon reasonable request to the authors

## 7.2 Acknowledgements

The authors would like to thank Brendan Collins and Sarah Meredith from Science Evidence Advice Division, Welsh Government, and Christoforos Pavlakis (public involvement representative) for their time, valuable insight and contribution to this work.

## APPENDIX

## Appendix 1: Ovid MEDLINE(R) search results for each condition

### Diabetes MEDLINE

|  | Search terms | Hits |
| --- | --- | --- |
| 1 | exp Diabetes Mellitus, Type 1/ or exp Diabetes Mellitus, Type 2/ or exp Diabetes Mellitus/ | 499,719 |
| 2 | diabet*.tw. | 752,072 |
| 3 | 1 or 2 | 817,185 |
| 4 | ((trend* or future or projected or projecti* or predicti* or forecast* or model*) adj10 (prevalence or incidence or epidemiolog* or burden* or cost* or morbidity or mortality or "life expectancy" or "risk factor*")).tw. | 274,541 |
| 5 | exp Prevalence/ | 340,067 |
| 6 | exp Incidence/ | 298,901 |
| 7 | 5 or 6 | 611,623 |
| 8 | (trend* or future or projected or projecti* or predicti* or forecast* or model*).tw. | 5,542,168 |
| 9 | 7 and 8 | 147,031 |
| 10 | 4 or 9 | 373,233 |
| 11 | exp Wales/ | 15,131 |
| 12 | exp United Kingdom/ | 388,650 |
| 13 | (UK or "united kingdom" or wales or england or scotland or ireland).tw. | 265,536 |
| 14 | 11 or 12 or 13 | 534,362 |
| 15 | 3 and 10 and 14 | 1544 |
| 16 | limit 15 to yr="2013 - 2023" | **981** |

### Breast cancer MEDLINE

|  | Search terms | Hits |
| --- | --- | --- |
| 1 | exp Breast Neoplasms/ | 337,884 |
| 2 | (breast adj3 (cancer* or carcinom* neoplasm* or tumor* or oncolog* or malignan*)).ti. | 225,326 |
| 3 | 1 or 2 | 374,441 |
| 4 | ((trend* or future or projected or projecti* or predicti* or forecast* or model*) adj10 (prevalence or incidence or epidemiolog* or burden* or cost* or morbidity or mortality or "life expectancy" or "risk factor*")).ti. | 38,322 |
| 5 | exp Prevalence/ | 340,151 |
| 6 | exp Incidence/ | 298,945 |
| 7 | 5 or 6 | 611,745 |
| 8 | (trend* or future or projected or projecti* or predicti* or forecast* or model*).tw. | 5,546,173 |
| 9 | 7 and 8 | 147,065 |
| 10 | 4 or 9 | 175,207 |
| 11 | exp Wales/ | 15,132 |
| 12 | exp United Kingdom/ | 388,680 |
| 13 | (UK or "united kingdom" or wales or england or scotland or ireland).tw. | 265,782 |
| 14 | 11 or 12 or 13 | 534,611 |
| 15 | 3 and 10 and 14 | 203 |
| 16 | limit 15 to yr="2012 - 2023" | **105** |

### Colorectal cancer MEDLINE

|  | Search terms | Hits |
| --- | --- | --- |
| 1 | exp Colorectal Neoplasms/ | 234,010 |
| 2 | ((colorectal* or colon* or rectal*) adj3 (cancer* or carcinom* or neoplasm* or tumo?r or oncolog* or malignan*)).ti. | 147,673 |
| 3 | 1 or 2 | 257,458 |
| 4 | ((trend* or future or projected or projecti* or predicti* or forecast* or model*) adj10 (prevalence or incidence or epidemiolog* or burden* or cost* or morbidity or mortality or "life expectancy" or "risk factor*")).ti. | 38,322 |
| 5 | exp Prevalence/ | 340,151 |
| 6 | exp Incidence/ | 298,945 |
| 7 | 5 or 6 | 611,745 |
| 8 | (trend* or future or projected or projecti* or predicti* or forecast* or model*).tw. | 5,546,173 |
| 9 | 7 and 8 | 147,065 |
| 10 | 4 or 9 | 175,207 |
| 11 | exp Wales/ | 15,132 |
| 12 | exp United Kingdom/ | 388,680 |
| 13 | (UK or "united kingdom" or wales or england or scotland or ireland).tw. | 265,782 |
| 14 | 11 or 12 or 13 | 534,611 |
| 15 | 3 and 10 and 14 | 154 |
| 16 | limit 15 to yr="2012 - 2023" | **100** |

### Lung cancer MEDLINE

|  | Search terms | Hits |
| --- | --- | --- |
| 1 | exp Lung Neoplasms/ | 270,604 |
| 2 | (lung adj3 (cancer* or carcinom* neoplasm* or tumo?r or oncolog* or malignan*)).ti. | 127,713 |
| 3 | 1 or 2 | 293,711 |
| 4 | ((trend* or future or projected or projecti* or predicti* or forecast* or model*) adj10 (prevalence or incidence or epidemiolog* or burden* or cost* or morbidity or mortality or "life expectancy" or "risk factor*")).ti. | 38,322 |
| 5 | exp Prevalence/ | 340,151 |
| 6 | exp Incidence/ | 298,945 |
| 7 | 5 or 6 | 611,745 |
| 8 | (trend* or future or projected or projecti* or predicti* or forecast* or model*).tw. | 5,546,173 |
| 9 | 7 and 8 | 147,065 |
| 10 | 4 or 9 | 175,207 |
| 11 | exp Wales/ | 15,132 |
| 12 | exp United Kingdom/ | 388,680 |
| 13 | (UK or "united kingdom" or wales or england or scotland or ireland).tw. | 265,782 |
| 14 | 11 or 12 or 13 | 534,611 |
| 15 | 3 and 10 and 14 | 147 |
| 16 | limit 15 to yr="2012 - 2023" | **76** |

### Prostatecancer MEDLINE

Prostate cancer MEDLINE
|  | Search terms | Hits |
| --- | --- | --- |
| 1 | exp prostatic neoplasms/ | 146,876 |
| 2 | (prostat* adj3 (cancer* or neoplasm* or tumor* or oncolog* or malignan* or carcinom* or adeno*)).ti. | 113,452 |
| 3 | 1 or 2 | 163,065 |
| 4 | ((trend* or future or projected or projecti* or predicti* or forecast* or model*) adj10 (prevalence or incidence or epidemiolog* or burden* or cost* or morbidity or mortality or "life expectancy" or "risk factor*")).ti. | 38,322 |
| 5 | exp Prevalence/ | 340,151 |
| 6 | exp Incidence/ | 298,945 |
| 7 | 5 or 6 | 611,745 |
| 8 | (trend* or future or projected or projecti* or predicti* or forecast* or model*).tw. | 5,546,173 |
| 9 | 7 and 8 | 147,065 |
| 10 | 4 or 9 | 175,207 |
| 11 | exp Wales/ | 15,132 |
| 12 | exp United Kingdom/ | 388,680 |
| 13 | (UK or "united kingdom" or wales or england or scotland or ireland).tw. | 265,782 |
| 14 | 11 or 12 or 13 | 534,611 |
| 15 | 3 and 10 and 14 | 94 |
| 16 | limit 15 to yr="2012 - 2023" | **48** |

### Cardiovascular disease MEDLINE

|  | Search terms | Hits |
| --- | --- | --- |
| 1 | (cardiovascular disease* or heart failure or peripheral vascular disease* or stroke or cerebrovascular accident or transient isch?emic attack* or atrial fibrillation or hypertension).tw. | 1,086,410 |
| 2 | exp hypertension/ | 314,564 |
| 3 | exp Atrial Fibrillation/ | 69,584 |
| 4 | exp Ischemic Attack, Transient/ | 21,771 |
| 5 | exp Stroke/ | 169,164 |
| 6 | exp Peripheral Vascular Diseases/ | 64,345 |
| 7 | exp Cardiovascular Diseases/ | 2,691,604 |
| 8 | 1 or 2 or 3 or 4 or 5 or 6 or 7 | 3,052,829 |
| 9 | ((trend* or future or projected or projecti* or predicti* or forecast* or model*) adj10 (prevalence or incidence or epidemiolog* or burden* or cost* or morbidity or mortality or "life expectancy" or "risk factor*")).tw. | 38,352 |
| 10 | exp Prevalence/ | 340,265 |
| 11 | exp Incidence/ | 299,020 |
| 12 | 10 or 11 | 611,932 |
| 13 | (trend* or future or projected or projecti* or predicti* or forecast* or model*).tw. | 1,150,565 |
| 14 | 12 and 13 | 25,514 |
| 15 | 9 or 14 | 53,679 |
| 16 | exp Wales/ | 15,134 |
| 17 | exp United Kingdom/ | 388,736 |
| 18 | (UK or "united kingdom" or wales or england or scotland or ireland).tw. | 265,943 |
| 19 | 16 or 17 or 18 | 534,775 |
| 15 | 8 and 15 and 19 | 540 |
| 16 | limit 20 to yr="2013 - 2023" | **311** |

### Dementia MEDLINE

|  | Search terms | Hits |
| --- | --- | --- |
| 1 | exp Dementia/ | 200,904 |
| 2 | (dementia or alzheimer*).tw. | 262,266 |
| 3 | 1 or 2 | 305,651 |
| 4 | ((trend* or future or projected or projecti* or predicti* or forecast* or model*) adj10 (prevalence or incidence or epidemiolog* or burden* or cost* or morbidity or mortality or "life expectancy" or "risk factor*")).tw. | 274,560 |
| 5 | exp Prevalence/ | 339,986 |
| 6 | exp Incidence/ | 298,888 |
| 7 | 5 or 6 | 611,533 |
| 8 | (trend* or future or projected or projecti* or predicti* or forecast* or model*).tw. | 5,542,257 |
| 9 | 7 and 8 | 147,007 |
| 10 | 4 or 9 | 373,243 |
| 11 | exp Wales/ | 15,128 |
| 12 | exp United Kingdom/ | 388,676 |
| 13 | (UK or "united kingdom" or wales or england or scotland or ireland).tw. | 265,727 |
| 14 | 11 or 12 or 13 | 534,553 |
| 15 | 3 and 10 and 14 | 439 |
| 16 | limit 15 to yr="2013 - 2023" | **289** |

### Mental health MEDLINE

|  | Search terms | Hits |
| --- | --- | --- |
| 1 | exp Schizophrenia/ | 114,372 |
| 2 | exp Bipolar Disorder/ | 44,818 |
| 3 | exp psychotic disorders/ | 57,969 |
| 4 | exp schizoaffective disorder/ | 57,969 |
| 5 | exp psychosis/ | 57,969 |
| 6 | Paranoid Disorders/ | 4251 |
| 7 | exp Personality Disorders/ | 44,831 |
| 8 | ("severe mental illness" or "persistent mental illness").ti. | 2575 |
| 9 | (schizo* or "mood disorder*" or "personality disorder*" or psychotic* or psychosis or psychoses).ti. | 141,910 |
| 10 | (bipolar or mania or Schizophrenia).tw. | 186,230 |
| 11 | ((chronic* or sever* or serious or persistent* or enduring or debilitating) adj2 (mental* or psychological*) adj2 (ill* or disorder* or health)).ti. | 6588 |
| 10 | depression.ti. | 122,127 |
| 11 | anxiety.ti. | 65,494 |
| 14 | 1 or 2 or 3 or 4 or 5 or 6 or 7 or 8 or 9 or 10 or 11 or 12 or 13 | 486,917 |
| 15 | ((trend* or future or projected or projecti* or predicti* or forecast* or model*) adj10 (prevalence or incidence or epidemiolog* or burden* or cost* or morbidity or mortality or "life expectancy" or "risk factor*")).ti. | 38,362 |
| 16 | exp Prevalence/ | 340,358 |
| 17 | exp Incidence/ | 299,088 |
| 18 | 16 or 17 | 612,090 |
| 19 | (trend* or future or projected or projecti* or predicti* or forecast* or model*).ti. | 1,150,870 |
| 20 | 18 and 19 | 25,522 |
| 21 | 15 or 20 | 53,691 |
| 22 | exp Wales/ | 15,137 |
| 23 | exp United Kingdom/ | 388,817 |
| 24 | (UK or "united kingdom" or wales or england or scotland or ireland).tw. | 90,093 |
| 25 | 22 or 23 or 24 | 417,231 |
| 26 | 14 and 21 and 25 | 37 |
| 27 | limit 26 to yr="2013 - 2023" | 17 |

### Multi-morbidity MEDLINE

|  | Search terms | Hits |
| --- | --- | --- |
| 1 | exp multiple chronic conditions/ | 686 |
| 2 | (multiple adj2 (condition* or illness* of disease*)).ti. | 776 |
| 3 | (multi-morbidit* or co-morbidit* or comorbidit* or multimorbid* or co morbidity or multi morbidity).ti. | 5608 |
| 4 | 1 or 2 or 3 | 6738 |
| 5 | ((trend* or future or projected or projecti* or predicti* or forecast* or model*) adj10 (prevalence or incidence or epidemiolog* or burden* or cost* or morbidity or mortality or "life expectancy" or "risk factor*")).ti. | 38,352 |
| 6 | exp Prevalence/ | 340,265 |
| 7 | exp Incidence/ | 299,020 |
| 8 | 6 or 7 | 611,932 |
| 9 | (trend* or future or projected or projecti* or predicti* or forecast* or model*).ti. | 1,150,565 |
| 10 | 8 and 9 | 25,514 |
| 11 | 5 or 10 | 53,679 |
| 12 | exp Wales/ | 15,134 |
| 13 | exp United Kingdom/ | 388,736 |
| 14 | (UK or "united kingdom" or wales or england or scotland or ireland).tw. | 265,943 |
| 15 | 11 or 12 or 13 | 534,775 |
| 16 | 3 and 10 and 14 | 12 |
| 17 | limit 15 to yr="2013 - 2023" | 10 |

### Grey literature sources searched

| Organisation | URL |
| --- | --- |
| Alzheimer's Disease International | https://www.alz.co.uk/ |
| Alzheimer's Europe | http://www.alzheimer-europe.org/ |
| Alzheimer's Society | https://www.alzheimers.org.uk/ |
| Cancer Research UK | https://www.cancerresearchuk.org/health-professional/cancer-statistics-for-the-uk |
| Dementia Profile | https://fingertips.phe.org.uk/profile-group/mental-health/profile/dementia/data#page/1 |
| Dementia statistics hub | https://dementiastatistics.org/statistics/prevalence-projections-in-the-uk/ |
| Global Burden of Disease | https://www.healthdata.org/gbd |
| Health Survey for England, 2017 | http://healthsurvey.hscic.gov.uk/support-guidance/public-health/health-survey-for-england-2017.aspx |
| Health Survey for England, 2021 | https://beta.ukdataservice.ac.uk/myaccount/credentials<br>https://beta.ukdataservice.ac.uk/datacatalogue/series/series?id=2000021 |
| International Diabetes Federation | https://idf.org/ |
| Macmillan Cancer Support | https://www.macmillan.org.uk/about-us/what-we-do/research/cancer-prevalence |
| National Social Care Data Portal for Wales | https://www.socialcaredata.wales/dataset?c=175&p=171 |
| NICE clinical knowledge summaries | https://cks.nice.org.uk/ |
| Office for Health Improvement & Disparities | https://www.gov.uk/government/organisations/office-for-health-improvement-and-disparities |
| Public Health Wales | https://phw.nhs.wales/ |
| Public Health Wales Observatory | https://phw.nhs.wales/services-and-teams/observatory/ |
| Stats Wales – Health and Social Care | https://statswales.gov.wales/Catalogue/Health-and-Social-Care |
| UK Health Forum | https://ukhealthforum.org.uk/our-work/ |
| Welsh Cancer Intelligence and Surveillance Unit (WCISU) | https://phw.nhs.wales/services-and-teams/welsh-cancer-intelligence-and-surveillance-unit-wcisu/cancer-incidence-in-wales-2002-2019/ |
| WHO – Global Cancer Observatory (Globocan) | https://gco.iarc.fr/today/home |
| World Health Organisation | https://www.who.int/ |
| World Obesity Federation | https://www.worldobesity.org/ |

## Appendix 2: Studies with other outcome data not extracted

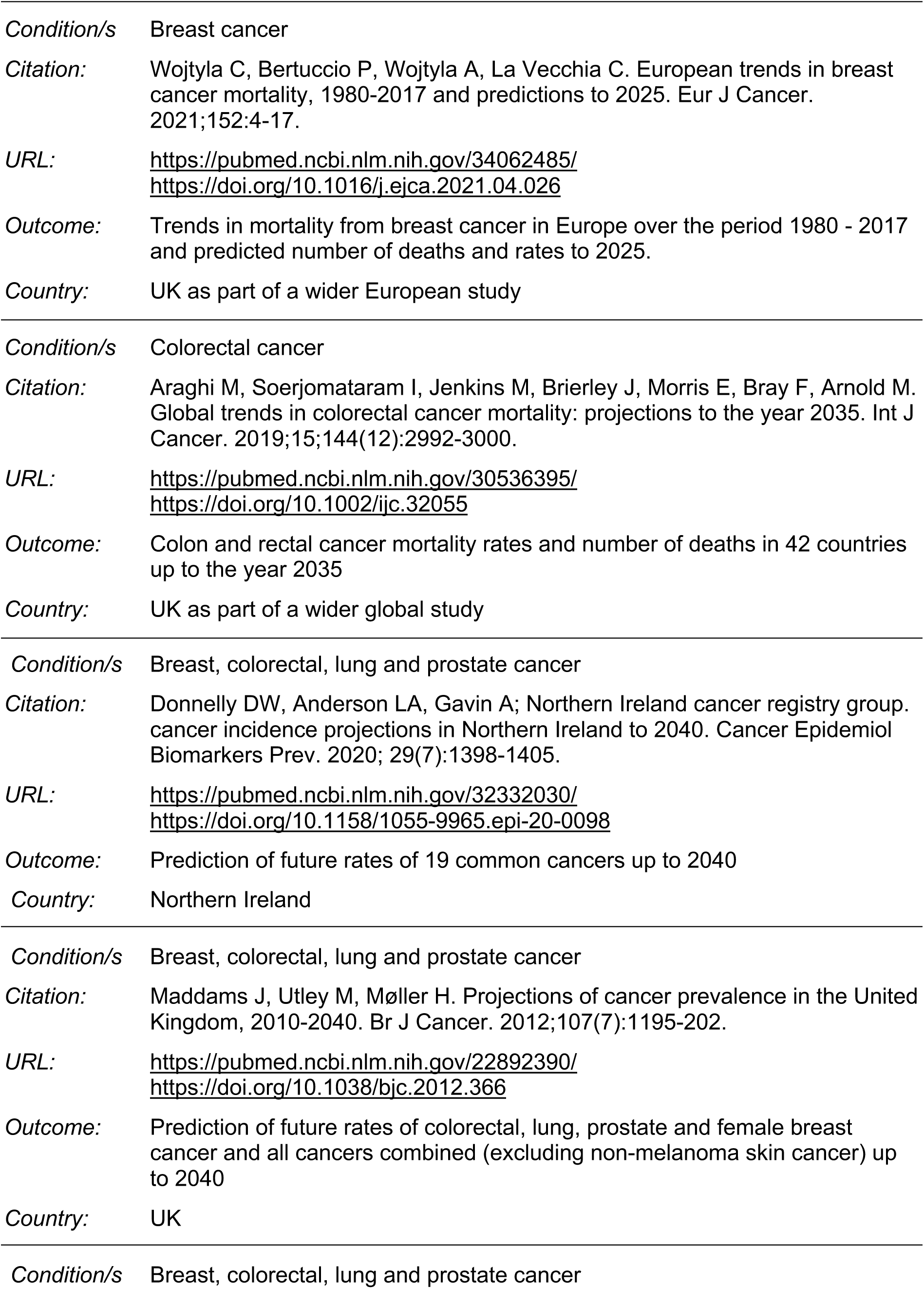
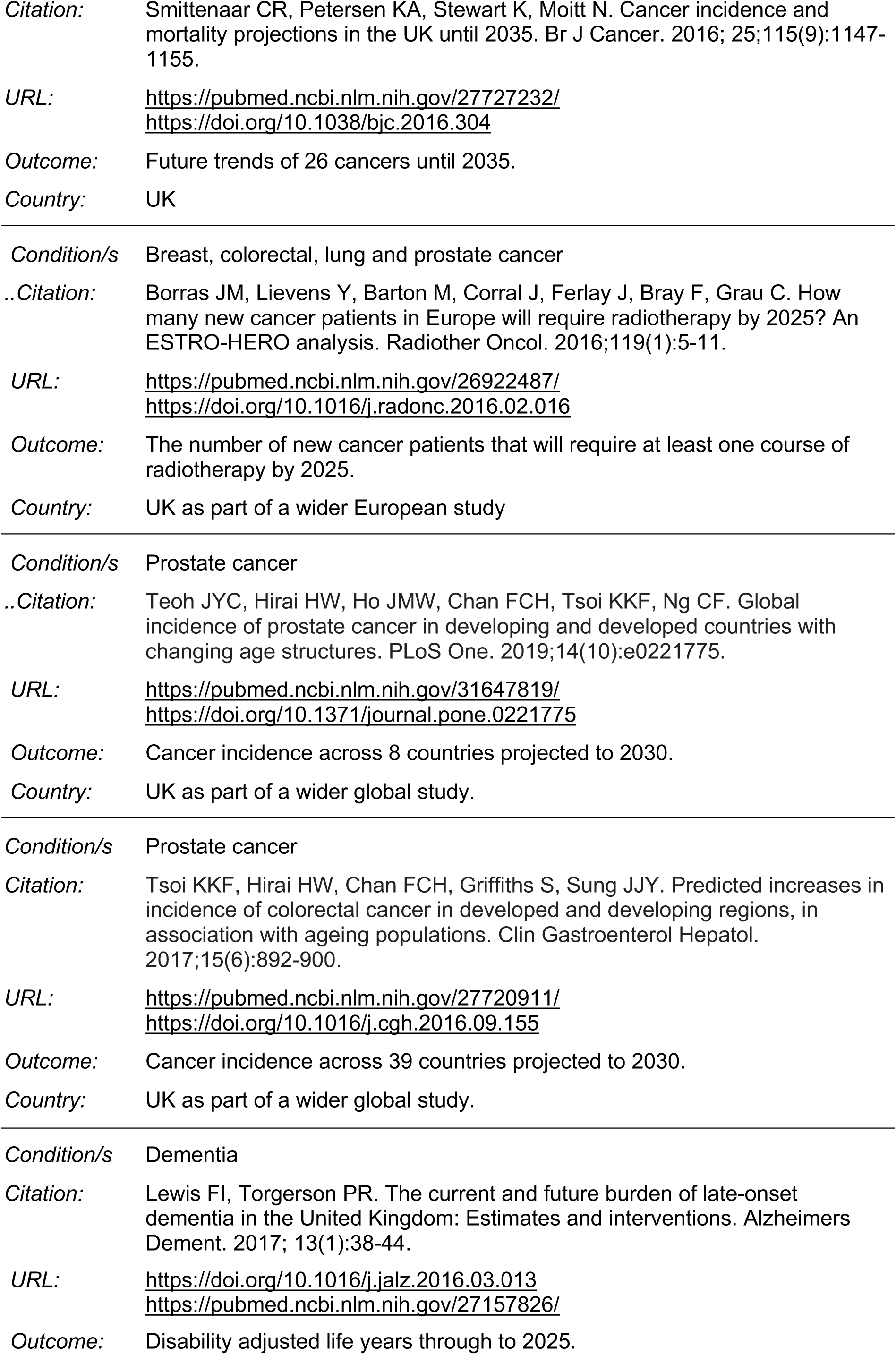
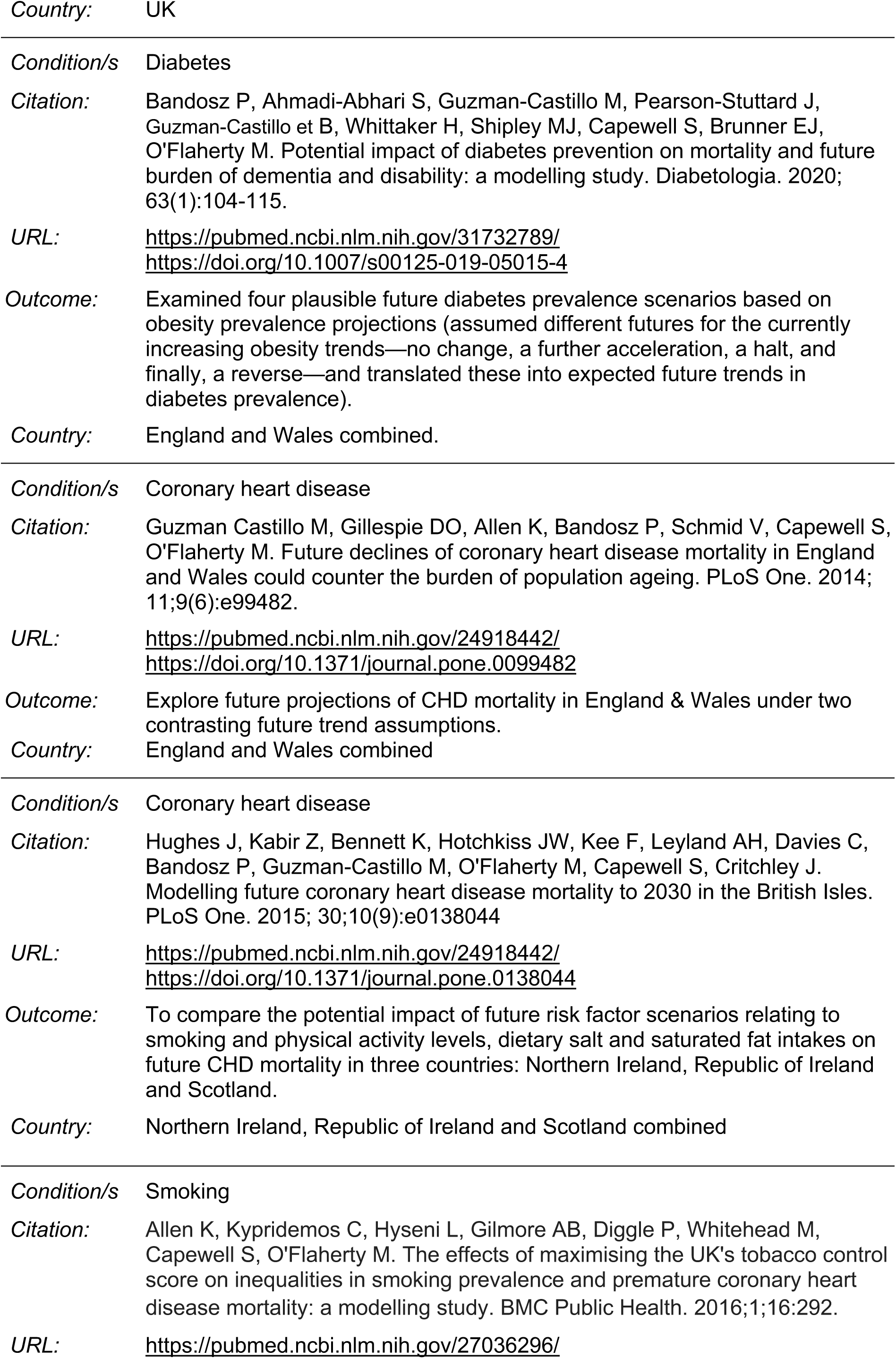
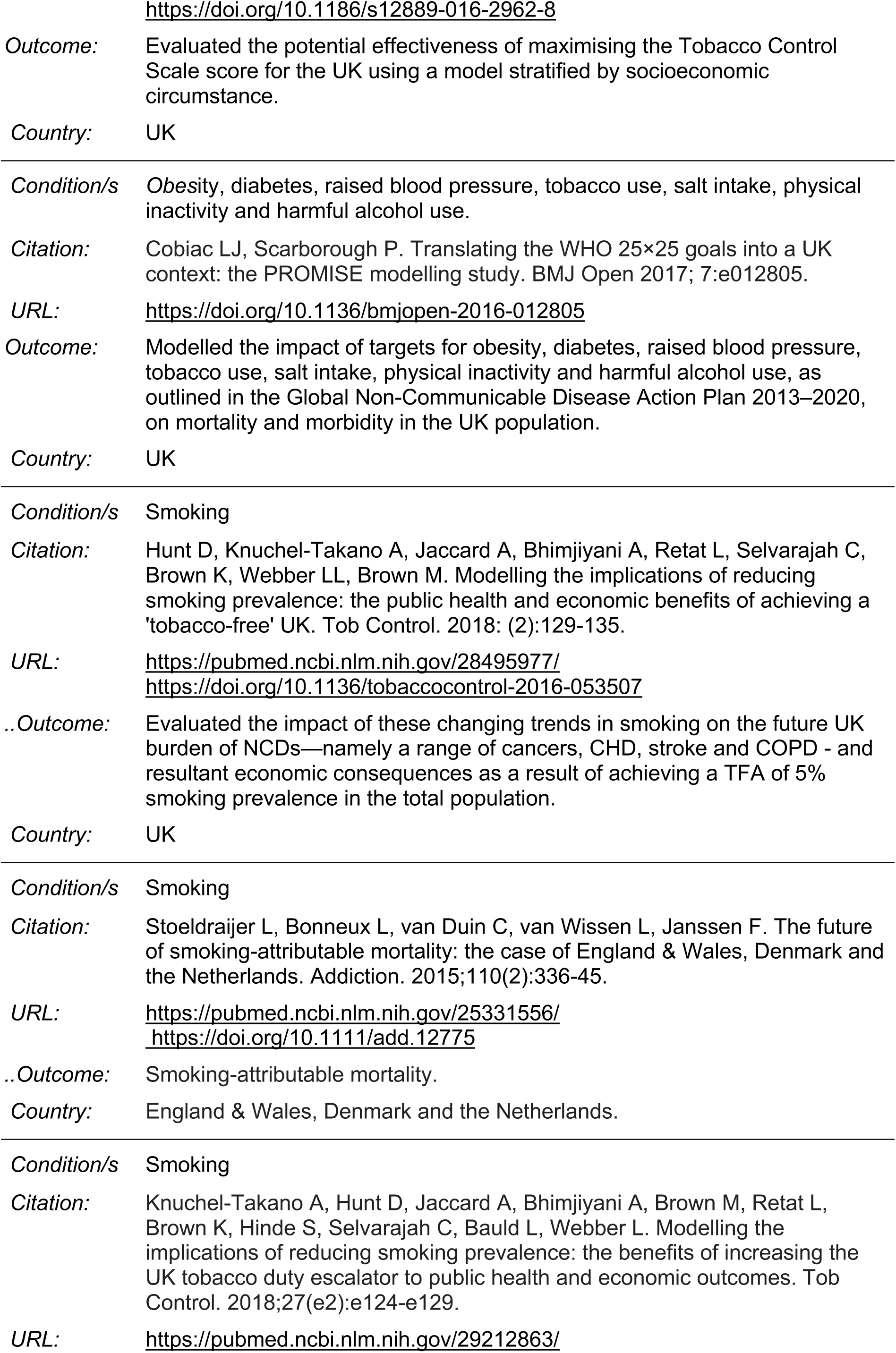
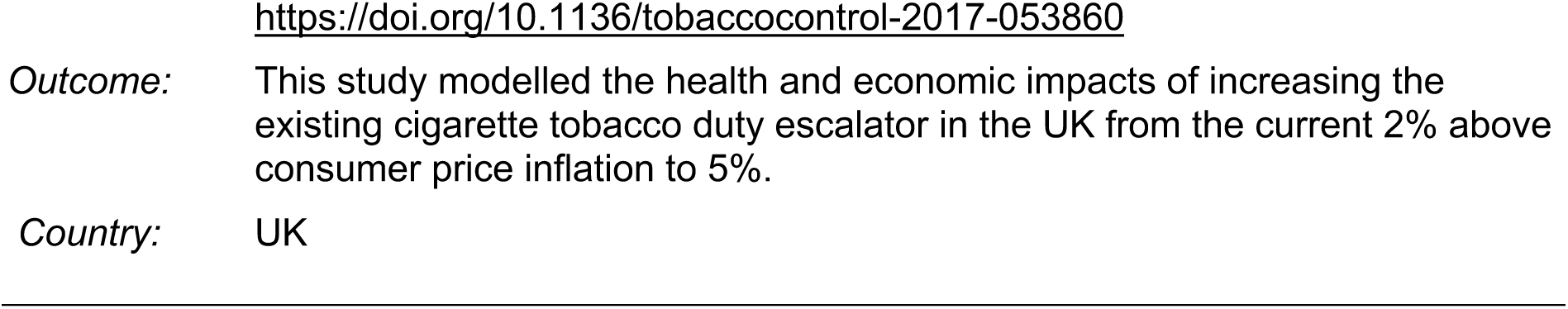

## REFERENCES

Ahmadi-Abhari S, Guzman-Castillo M, Bandosz P, et al. (2017). Temporal trend in dementia incidence since 2002 and projections for prevalence in England and Wales to 2040: modelling study. Bmj. 358: j2856. https://doi.org/10.1136/bmj.j2856

Allen K, Kypridemos C, Hyseni L, et al. (2016). The effects of maximising the UK’s tobacco control score on inequalities in smoking prevalence and premature coronary heart disease mortality: a modelling study. BMC Public Health. 16: 292. https://doi.org/10.1186/s12889-016-2962-8

Alzheimer’s Research UK. (2023). Statistics about dementia. Dementia Statistics Hub. Available at: https://dementiastatistics.org/statistics-about-dementia/ [Accessed 07/04/2023].

Alzheimer’s Society. (2021). Risk factors for dementia – Factsheet 450LP. Alzheimer’s Society. Available at: https://www.alzheimers.org.uk/sites/default/files/pdf/factsheet_risk_factors_for_dementia.pdf [Accessed 07/04/2023].

Ampofo AG, Boateng EB. (2020). Beyond 2020: modelling obesity and diabetes prevalence. Diabetes research and clinical practice. 167: 108362. https://doi.org/10.1016/j.diabres.2020.108362

Bandosz P, Ahmadi-Abhari S, Guzman-Castillo M, et al. (2020). Potential impact of diabetes prevention on mortality and future burden of dementia and disability: a modelling study. Diabetologia. 63(1): 104–15. https://doi.org/10.1007/s00125-019-05015-4

Barnett K, Mercer SW, Norbury M, et al. (2012). Epidemiology of multimorbidity and implications for health care, research, and medical education: a cross-sectional study. Lancet. 380(9836): 37–43. https://doi.org/10.1016/s0140-6736(12)60240-2

Bebbington P, Rai D, Strydom A, et al. (2016). Chapter 5: Psychotic disorder. In: McManu sS, Bebbington P, Jenkins R & Brugha T (eds.) Adult Psychiatric Morbidity Survey 2014. Leeds: NHS Digital.

Bhimjiyani A, Knuchel-Takano A, Hunt D, et al. (2016a). Tipping the scales: Why preventing obesity makes economic sense. Cancer Research UK, UK Health Forum. Available at: https://ukhealthforum.org.uk/wp-content/uploads/2019/02/Tipping-The-Scales-CRUK-Full-Report-FINAL.pdf

Bhimjiyani A, Knuchel-Takano A, Jaccard A, et al. (2016b). Extended obesity technical report: Baseline scenario, Hypothetical scenarios, SSB excise tax policy scenario. Cancer Research UK, UK Health Forum. Available at: https://ukhealthforum.org.uk/wp-content/uploads/2018/11/CRUK-UKHF-Extended-Obesity-Technical-Report.pdf

Borras JM, Lievens Y, Barton M, et al. (2016). How many new cancer patients in Europe will require radiotherapy by 2025? An ESTRO-HERO analysis. Radiotherapy and Oncology. 119: S153. https://doi.org/10.1016/j.radonc.2016.02.016

British Heart Foundation. (2020). The British Heart Foundation. BHF UK Factsheet - Heart and Circulatory Disease Statistics. Available at: https://www.bhf.org.uk/what-we-do/our-research/heart-statistics/heart-statistics-publications/cardiovascular-disease-statistics-2020 [Accessed 16/04/2023].

British Heart Foundation. (2023). Wales Factsheet. Available at: https://www.bhf.org.uk/-/media/files/for-professionals/research/heart-statistics/bhf-cvd-statistics-wales-factsheet.pdf?rev=aac0954754d14efb8da6348599b7514a&hash=F609B99EC0E150A7F70845542CAE77C7 [Accessed 16/04/2023].

Cancer Research UK. (2018a). Bowel cancer risk. Available at: https://www.cancerresearchuk.org/health-professional/cancer-statistics/statistics-by-cancer-type/bowel-cancer/risk-factors#heading-One [Accessed 30/03/2023].

Cancer Research UK. (2018b). Breast cancer risk. Available at: https://www.cancerresearchuk.org/health-professional/cancer-statistics/statistics-by-cancer-type/breast-cancer/risk-factors [Accessed 30/03/2023].

Cancer Research UK. (2018c). Lung cancer risk. Available at: https://www.cancerresearchuk.org/health-professional/cancer-statistics/statistics-by-cancer-type/lung-cancer/risk-factors [Accessed 30/03/2023].

Cancer Research UK. (2018d). Prostate cancer risk. Available at: https://www.cancerresearchuk.org/health-professional/cancer-statistics/statistics-by-cancer-type/prostate-cancer/risk-factors [Accessed 30/03/2023].

Cancer Research UK. (2020). Cancer incidence for common cancers. Available at: https://www.cancerresearchuk.org/health-professional/cancer-statistics/incidence/common-cancers-compared#ref- [Accessed 08/04/2023].

Cancer Research UK. (2023a). Bowel cancer incidence statistics. Available at: https://www.cancerresearchuk.org/health-professional/cancer-statistics/statistics-by-cancer-type/bowel-cancer/incidence#heading-Four [Accessed 08/04/2023].

Cancer Research UK. (2023b). Breast cancer incidence (invasive) statistics. Available at: https://www.cancerresearchuk.org/health-professional/cancer-statistics/statistics-by-cancer-type/breast-cancer/incidence-invasive#heading-Four [Accessed 08/04/2023].

Cancer Research UK. (2023c). Lung cancer incidence statistics. Available at: https://www.cancerresearchuk.org/health-professional/cancer-statistics/statistics-by-cancer-type/lung-cancer/incidence#heading-Four [Accessed 08/04/2023].

Cancer Research UK. (2023d). Our calculations explained - Age-period cohort models for projecting cancer incidence and mortality. Available at: https://www.cancerresearchuk.org/health-professional/cancer-statistics/cancer-stats-explained/our-calculations-explained#heading-Zero [Accessed 08/04/2023].

Cancer Research UK. (2023e). Prostate cancer incidence statistics. Available at: https://www.cancerresearchuk.org/health-professional/cancer-statistics/statistics-by-cancer-type/prostate-cancer/incidence#heading-Three [Accessed 08/04/2023].

Cassell A, Edwards D, Harshfield A, et al. (2018). The epidemiology of multimorbidity in primary care: a retrospective cohort study. British JOurnal of General Practice. 68(669): e245–e51. https://doi.org/10.3399/bjgp18X695465

Cobiac LJ, Scarborough P. (2021). Modelling future trajectories of obesity and body mass index in England. PLoS ONE 16(6): e0252072. https://doi.org/10.1371/journal.pone.0252072

Collins B, Bandosz P, Guzman-Castillo M, et al. (2022). What will the cardiovascular disease slowdown cost? Modelling the impact of CVD trends on dementia, disability, and economic costs in England and Wales from 2020-2029. PLoS ONE [Electronic Resource]. 17(6): e0268766. https://dx.doi.org/10.1371/journal.pone.0268766

Cowie M, Mian S, Morris J, et al. (2016). Heart failure prevalence models for small populations: technical document produced for Public Health England. Imperical College London Available at: https://fingertips.phe.org.uk/documents/Heart-failure-prevalence-model-Technical-Document-v2.1.docx [Accessed 07/04/2023].

Department of Health and Social Care. (2021). Integration and innovation: working together to improve health and social care for all. Department of Health and Social Care,. Available at: https://assets.publishing.service.gov.uk/government/uploads/system/uploads/attachment_data/file/960549/integration-and-innovation-working-together-to-improve-health-and-social-care-for-all-print-version.pdf [Accessed 27/04/2023].

Diabetes UK. (2023a). Diabetes in Wales Available at: https://www.diabetes.org.uk/in_your_area/wales/diabetes-in-wales [Accessed 16/04/2023].

Diabetes UK. (2023b). Diabetes prevalence 2019. Available at: https://www.diabetes.org.uk/professionals/position-statements-reports/statistics/diabetes-prevalence-2019 [Accessed 16/04/2023].

Diabetes UK. (2023c). Risk factors for type 2 diabetes. Available at: https://www.diabetes.org.uk/preventing-type-2-diabetes/diabetes-risk-factors [Accessed 08/04/2023].

Donnelly DW, Anderson LA, Gavin A. (2020). Cancer incidence projections in Northern Ireland to 2040. Cancer Epidemiology Biomarkers and Prevention. 29: 1398–405. https://doi.org/10.1158/1055-9965.EPI-20-0098

Everest G, Marshall L, Fraser C, et al. (2022). Addressing the leading risk factors for ill health: A review of government policies tackling smoking, poor diet, physical inactivity and harmful alcohol use in England. The Health Foundation;. Available at: https://doi.org/10.37829/HF-2022-P10 [Accessed 27/04/2023].

GBD 2017 Disease and Injury Incidence and Prevalence Collaborators. (2018). Global, regional, and national incidence, prevalence, and years lived with disability for 354 diseases and injuries for 195 countries and territories, 1990-2017: a systematic analysis for the Global Burden of Disease Study 2017. Lancet. 392(10159): 1789–858. https://doi.org/10.1016/s0140-6736(18)32279-7

Globocan. (2020). Cancer today. Available at: https://gco.iarc.fr/today/home [Accessed 30/03/2023].

Guzman-Castillo M, Ahmadi-Abhari S, Bandosz P, et al. (2017). Forecasted trends in disability and life expectancy in England and Wales up to 2025: a modelling study. Lancet Public Health. 2(7): e307–e13. https://doi.org/10.1016/s2468-2667(17)30091-9

Haider S, Thayakaran R, Subramanian A, et al. (2021). Disease burden of diabetes, diabetic retinopathy and their future projections in the UK: cross-sectional analyses of a primary care database. BMJ Open. 11(7): e050058. https://doi.org/10.1136/bmjopen-2021-050058

Hannah V, Woolridge A, Rich L, et al. (2021). The primary care needs of people living with overweight and obesity in Wales: summary Public Health Wales. Available at: https://phw.nhs.wales/publications/publications1/the-primary-care-needs-of-people-living-with-overweight-and-obesity-in-wales-summary/ [Accessed 07/04/2023].

Hunt D, Knuchel-Takano A, Jaccard A, et al. (2018). Modelling the implications of reducing smoking prevalence: the public health and economic benefits of achieving a ‘tobacco-free’ UK. Tobacco Control. 27(2): 129–35. https://doi.org/10.1136/tobaccocontrol-2016-053507

Janssen F, Bardoutsos A, Vidra N. (2020). Obesity prevalence in the long-term future in 18 European countries and in the USA. Obesity Facts. 13(5): 514–27. https://doi.org/10.1159/000511023

Jiang C, Chen Q, Xie M. (2020). Smoking increases the risk of infectious diseases: A narrative review. Tobacco Induced Diseases. 18: 60. https://doi.org/10.18332/tid/123845

Keaver L, Perez-Ferrer C, Jaccard A, et al. (2019). Future trends in social inequalities in obesity in England, Wales and Scotland. Journal of Public Health. 42(1): e52–e8. https://doi.org/10.1093/pubmed/fdz022

Keaver L, Xu B, Jaccard A, et al. (2020). Morbid obesity in the UK: A modelling projection study to 2035. Scandinavian Journal of Public Health. 48(4): 422–7. https://doi.org/10.1177/1403494818794814

King D, Wittenberg R, Patel A, et al. (2020). The future incidence, prevalence and costs of stroke in the UK. Age & Ageing. 49(2): 277–82. https://doi.org//10.1093/ageing/afz163

Kingston A, Robinson L, Booth H, et al. (2018). Projections of multi-morbidity in the older population in England to 2035: estimates from the Population Ageing and Care Simulation (PACSim) model. Age & Ageing. 47(3): 374–80. https://doi.org/10.1093/ageing/afx201

Knuchel-Takano A, Hunt D, Jaccard A, et al. (2018). Modelling the implications of reducing smoking prevalence: the benefits of increasing the UK tobacco duty escalator to public health and economic outcomes. Tobacco Control. 27(e2): e124–e9. https://doi.org/10.1136/tobaccocontrol-2017-053860

Kypridemos C, Reeve J, Buchan I, et al. (2017). PL02 Tobacco control in england: using microsimulation modelling to quantify the potential impact of a tobacco-free generation or a total ban. Journal of Epidemiology and Community Health. 71(Suppl 1): A51–A. https://doi.org/10.1136/jech-2017-SSMAbstracts.101

Lane DA, Skjøth F, Lip GYH, et al. (2017). Temporal trends in incidence, prevalence, and mortality of atrial fibrillation in primary care. Journal of the American Heart Association 6(5): e005155. https://doi.org/10.1161/jaha.116.005155

Li F, Qin W, Zhu M, et al. (2021). Model-based projection of dementia prevalence in China and worldwide: 2020-2050. Journal of Alzheimer’s Disease. 82(4): 1823–31. https://doi.org/10.3233/JAD-210493

Li L, Scott CA, Rothwell PM. (2020). Trends in stroke incidence in high-income countries in the 21st Century: population-based study and systematic review. Stroke. 51(5): 1372–80. https://doi.org/10.1161/STROKEAHA.119.028484

Macmillan Cancer Support. (2015). 20 year cancer prevalence in Wales: An increasingly granular understanding of the cancer population. Available at: https://www.macmillan.org.uk/documents/aboutus/research/researchandevaluationreports/ourresearchpartners/20yrcancerprevalencewales.pdf [Accessed 08/04/2023].

Macmillan Cancer Support. (2020). Prevalence by cancer type, nation, sex and year. Available at: https://www.macmillan.org.uk/dfsmedia/1a6f23537f7f4519bb0cf14c45b2a629/5192-10061/macmillan-2020-cancer-prevalence-figures-and-methodology.

Maddams J, Utley M, Moller H. (2012). Projections of cancer prevalence in the United Kingdom, 2010-2040. British Journal of Cancer. 107: 1195–202. https://doi.org/10.1038/bjc.2012.366

Marwaha S, Sal N, Bebbington P. (2016). Chapter 9: Bipolar disorder. In: McManus S, Bebbington P, Jenkins R & Brugha T (eds.) Mental health and wellbeing in England: Adult Psychiatric Morbidity Survey 2014. Leeds: NHS Digital

McDaid, D., Park, A.-L., Davidson, G., John, A., Knifton, L., Morton, A., & Thorpe, L. (2022). The economic case for investing in the prevention of mental health conditions in the UK. Mental Health Foundation, February 2022 [Accessed 08/06/2023].

McDonald K, Ding T, Ker H, et al. (2021). Using epidemiological evidence to forecast population need for early treatment programmes in mental health: a generalisable Bayesian prediction methodology applied to and validated for first-episode psychosis in England. The British Journal of Psychiatry. 219(1): 383–91. https://doi.org/10.1192/bjp.2021.18

Merikangas KR, Jin R, He JP, et al. (2011). Prevalence and correlates of bipolar spectrum disorder in the world mental health survey initiative. Archives of General Psychiatry. 68(3): 241–51. https://doi.org/10.1001/archgenpsychiatry.2011.12

Murray E, Shelton N, Head J et al. (2022) Health and Place: How levelling up health can keep older workers working. International Longevity Centre – UK and The Health Foundation, 2022. [Accessed 13/06/2023].

NICE. (2018). Breast cancer - managing FH: What is known about the genetic risk factors? Clinical Knowledge Summaries Available at: https://cks.nice.org.uk/topics/breast-cancer-managing-fh/background-information/genetic-risk-factors/ [Accessed 08/04/2023].

NICE. (2021). Psychosis and schizophrenia: What causes psychotic disorders? Clinical Knowledge Summaries. Available at: https://cks.nice.org.uk/topics/psychosis-schizophrenia/background-information/causes-risk-factors/ [Accessed 08/04/2023]

NICE. (2022a). Bipolar disorder: How common is it? Clinical Knowledge Summaries. Available at: https://cks.nice.org.uk/topics/bipolar-disorder/background-information/incidence-prevalence/ [Accessed 08/04/2023].

NICE. (2022b). Bipolar disorder: What causes it? Clinical Knowledge Summaries. Available at: https://cks.nice.org.uk/topics/bipolar-disorder/background-information/causes/ [Accessed 08/04/2023].

NICE. (2022c). Dementia: What are the risk factors? Clincial Knowledge Summaries Available at: https://cks.nice.org.uk/topics/dementia/background-information/risk-factors/ [Accessed 16/04/2023].

NICE. (2022d). Depression: How common is it? Clinical Knowledge Summaries. Available at: https://cks.nice.org.uk/topics/depression/background-information/prevalence/ [Accessed 08/04/2023].

NICE. (2022e). Peripheral arterial disease: How common is it? Available at: https://cks.nice.org.uk/topics/peripheral-arterial-disease/background-information/prevalence/ [Accessed 16/04/2023].

NICE. (2022f). Peripheral arterial disease: What are the risk factors? Available at: https://cks.nice.org.uk/topics/peripheral-arterial-disease/background-information/risk-factors/ [Accessed 16/04/2023].

NICE. (2022g). Prostate cancer: What are the risk factors for prostate cancer? Clinical Knowledge Summaries Available at: https://cks.nice.org.uk/topics/prostate-cancer/background-information/risk-factors/ [Accessed 08/04/2023].

NICE. (2022h). Stroke and TIA: What are the risk factors for stroke and TIA? Available at: https://cks.nice.org.uk/topics/stroke-tia/background-information/risk-factors/ [Accessed 16/04/2023].

NICE. (2023a). Diabetes - type 1: What are the causes and risk factors? Clincial Knowledge Summaries Available at: https://cks.nice.org.uk/topics/diabetes-type-1/background-information/causes-risk-factors/ [Accessed 16/04/2023].

NICE. (2023b). Diabetes - type 2: What are the risk factors? Clinical Knowledge Summaries. Available at: https://cks.nice.org.uk/topics/diabetes-type-2/background-information/risk-factors/ [Accessed 16/04/2023].

NICE. (2023c). Generalized anxiety disorder: How common is it? Clinical Knowledge Summaries. Available at: https://cks.nice.org.uk/topics/generalized-anxiety-disorder/background-information/prevalence/ [Accessed 08/04/2023].

NICE. (2023d). Generalized anxiety disorder: What are the risk factors for the development of generalized anxiety disorder? Clinical Knowledge Summaries. Available at: https://cks.nice.org.uk/topics/generalized-anxiety-disorder/background-information/risk-factors/ [Accessed 08/04/2023].

NICE. (2023e). Heart failure: Chronic: How common is it? Clinical Knowledge Summaries. Available at: https://cks.nice.org.uk/topics/heart-failure-chronic/background-information/prevalence/ [Accessed 16/04/2023].

NICE. (2023f). Hypertension: How common is it? Clinical Knowledge Summaries. Available at: https://cks.nice.org.uk/topics/hypertension/background-information/prevalence/ [Accessed 16/04/2023].

NICE. (2023g). Hypertension: What are the risk factors? Available at: https://cks.nice.org.uk/topics/hypertension/background-information/risk-factors/ [Accessed 16/04/2023].

NICE. (2023h). Obesity: How common is it? Clinical Knowledge Summaries. Available at: https://cks.nice.org.uk/topics/obesity/background-information/prevalence/ Accessed 19/04/2023].

NICE. (2023i). Obesity: What are the causes and risk factors? Available at: https://cks.nice.org.uk/topics/obesity/background-information/causes-risk-factors/ [Accessed 19/04/2023].

Office for Health Improvement & Disparities. (2022a). Atrial fibrillation: QOF prevalence (all ages). Available at: https://fingertips.phe.org.uk/search/atrial%20fibriliation#page/4/gid/1/pat/159/par/K02000001/ati/15/are/E92000001/iid/280/age/1/sex/4/cat/-1/ctp/-1/yrr/1/cid/4/tbm/1 [Accessed 16/04/2023].

Office for Health Improvement & Disparities. (2022b). CHD: QOF prevalence (all ages). Available at: https://fingertips.phe.org.uk/search/CHD#page/4/gid/1/pat/159/par/K02000001/ati/15/are/E92000001/iid/273/age/1/sex/4/cat/-1/ctp/-1/yrr/1/cid/4/tbm/1 [Accessed 16th April 2023].

Office for Health Improvement & Disparities. (2022c). Depression: QOF prevalence (18+ yrs). Available at: https://fingertips.phe.org.uk/search/depression#page/4/gid/1/pat/159/par/K02000001/ati/15/are/E92000001/iid/848/age/168/sex/4/cat/-1/ctp/-1/yrr/1/cid/4/tbm/1.

Office for Health Improvement & Disparities. (2022d). Diabetes: QOF preva;ence (17+ years) Available at: https://fingertips.phe.org.uk/search/diabetes#page/4/gid/1/pat/159/par/K02000001/ati/15/are/E92000001/iid/241/age/187/sex/4/cat/-1/ctp/-1/yrr/1/cid/4/tbm/1 [Accessed 16/04/2023]

Office for Health Improvement & Disparities. (2022e). Heart failure: QOF prevalence (all ages). Available at: https://fingertips.phe.org.uk/search/heart%20failure [Accessed 16/04/2023].

Office for Health Improvement & Disparities. (2022f). Mixed anxiety and depressive disorder: estimated % of population aged 16-74 Available at: https://fingertips.phe.org.uk/search/depression%20and%20anxiety%20prevalence#page/4/gid/1/pat/159/par/K02000001/ati/15/are/E92000001/iid/90419/age/240/sex/4/cat/-1/ctp/-1/yrr/1/cid/4/tbm/1 [Accessed 19/03/2023].

Office for Health Improvement & Disparities. (2022g). Obesity: QOF prevalence (18+ yrs) Available at: https://fingertips.phe.org.uk/search/obesity#page/4/gid/1/pat/159/par/K02000001/ati/15/are/E92000001/iid/92588/age/168/sex/4/cat/-1/ctp/-1/yrr/1/cid/4/tbm/1 [Accessed 19/04/2023].

Office for Health Improvement & Disparities. (2022h). PAD: QOF prevalence (all ages). Available at: https://fingertips.phe.org.uk/search/Peripheral%20arterial%20disease [Accessed 16/04/2023].

Office for Health Improvement & Disparities. (2022i). Smoking: QOF prevalence (15+ yrs). Available at: https://fingertips.phe.org.uk/search/smoking#page/4/gid/1/pat/159/par/K02000001/ati/15/are/E92000001/iid/91280/age/188/sex/4/cat/-1/ctp/-1/yrr/1/cid/4/tbm/1 [Accessed 19/04/2023].

Office for Health Improvement & Disparities. (2022j). Stroke: QOF prevalence (all ages). Available at: https://fingertips.phe.org.uk/search/stroke#page/4/gid/1/pat/159/par/K02000001/ati/15/are/E92000001/iid/212/age/1/sex/4/cat/-1/ctp/-1/yrr/1/cid/4/tbm/1 [Accessed 27/04/2023]

Office for National Statistics. (2022). Adult smoking habits in the UK: 2021: Cigarette smoking habits among adults in the UK, including the proportion of people who smoke, demographic breakdowns, changes over time and use of e-cigarettes. Available at: file:///C:/Users/wnsdje/Downloads/Adult%20smoking%20habits%20in%20the%20UK%202021.pdf [Accessed 27/04/2023].

Pashayan N, Antoniou AC, Ivanus U, et al. (2020). Personalized early detection and prevention of breast cancer: ENVISION consensus statement. Nature Reviews Clinical Oncology. 17(11): 687–705. https://doi.org/10.1038/s41571-020-0388-9

Pathirana, T. I., & Jackson, C. A. (2018). Socioeconomic status and multimorbidity: a systematic review and meta-analysis. Australian and New Zealand Journal of Public Health, 42(2), 186–194. https://doi.org/10.1111/1753-6405.12762

Pérez-Ferrer C, Jaccard A, Knuchel-Takano A, et al. (2018). Inequalities in smoking and obesity in Europe predicted to 2050: Findings from the EConDA project. Scandinavian Journal of Public Health. 46(5): 530–40. https://doi.org/10.1177/1403494818761416

Pineda E, Sanchez-Romero LM, Brown M, et al. (2018). Forecasting future trends in obesity across Europe: the value of improving surveillance. Obesity Facts. 11(5): 360–71. https://doi.org/10.1159/000492115

Prince M, Knapp M, Guerchet M, et al. (2014). Dementia UK: Update. Alzheimer’s Society.

Public Health England. (2017). Technical document for sub-national English atrial fibrillation prevalence estimates: Application of Age-sex rates in a Swedish region to the English population Available at: https://assets.publishing.service.gov.uk/government/uploads/system/uploads/attachment_data/file/644868/atrial_fibrillation_AF_prevalence_estimates_technical_document.pdf [Accessed 07/04/2023].

Public Health England. (2019). Atrial Fibrillation prevalence estimates. Available at: https://assets.publishing.service.gov.uk/government/uploads/system/uploads/attachment_data/file/879448/AF_prevalence_estimates.xlsx [Accessed 07/04/2023].

Public Health England. (2020). Hypertension prevalence estimates in England, 2017. Available at: https://assets.publishing.service.gov.uk/government/uploads/system/uploads/attachment_data/file/873605/Summary_of_hypertension_prevalence_estimates_in_England1_.pdf [Accessed 16/04/2023].

Public Health Wales Observatory. (2017a). Health and its determinants in Wales. Information strategic planning. Report. Public Health Wales Observatory. Available at: https://phw.nhs.wales/services-and-teams/observatory/data-and-analysis/publication-documents/health-and-its-determinants-in-wales-2018/health-and-determinants-in-wales-report-eng-pptx/ [Accessed 09/03/2023].

Public Health Wales Observatory. (2017b). Health and its determinants in Wales. Informing Public Health Wales strategic planning. Interim report technical guide Available at: https://phw.nhs.wales/services-and-teams/observatory/data-and-analysis/publication-documents/health-and-its-determinants-in-wales-2018/health-and-determinants-in-wales-interim-report-tech-guide-pdf/ [Accessed 21/04/2023].

Public Health Wales Observatory. (2018a). Health and its determinants in Wales. Available at: https://phw.nhs.wales/services-and-teams/observatory/data-and-analysis/publication-documents/health-and-its-determinants-in-wales-2018/health-and-its-determinants-in-wales-data-xlsx/ [Accessed 19/04/2023].

Public Health Wales Observatory. (2018b). Health and its determinants in Wales (2018). Available at: https://phw.nhs.wales/services-and-teams/observatory/data-and-analysis/health-and-its-determinants-in-wales-2018/ [Accessed 16/04/2023].

Public Health Wales Observatory. (2023). Cardiovascular disease. Available at: https://phw.nhs.wales/services-and-teams/observatory/data-and-analysis/cardiovascular-disease/ [Accessed 12/04/2023].

Pugliese M, Newson R, Su B, et al. (2016). Health Survey for England hypertension analysis and local prevalence models. Project for Public Health England Imperial College. Available at: https://fingertips.phe.org.uk/documents/Hypertension-model-2016-Technical-Document-v2.5%20(2).docx [Accessed 16/04/2023].

Raleigh V, Jefferies D, Wellings D. (2022). Cardiovascular disease in England: spporting leaders and taking action King’s Fund. Available at: https://www.kingsfund.org.uk/sites/default/files/2022-11/CVD_Report_Web.pdf [Accessed 16/04/2023].

Rijken M, Struckmann V, Dyakova M, et al. (2023). ICARE4EU: improving care for people with multiple chronic conditions in Europe. Eurohealth. 3: 29–31.

Savarese G, Becher PM, Lund LH, et al. (2023). Global burden of heart failure: a comprehensive and updated review of epidemiology. Cardiovascular Research. 118(17): 3272–87. https://doi.org/10.1093/cvr/cvac013

Scarborough P, Kaur A, Cobiac LJ. (2022). Forecast of myocardial infarction incidence, events and prevalence in England to 2035 using a microsimulation model with endogenous disease outcomes. PLoS ONE. 17(6): e0270189. https://doi.org/10.1371/journal.pone.0270189

Shu J, Santulli G. (2018). Update on peripheral artery disease: Epidemiology and evidence-based facts. Atherosclerosis. 275: 379–81. https://doi.org/10.1016/j.atherosclerosis.2018.05.033

Smittenaar CR, Petersen KA, Stewart K, et al. (2016). Cancer incidence and mortality projections in the UK until 2035. British Journal of Cancer. 115: 1147–55. https://doi.org//10.1038/bjc.2016.304

Social Care Wales. (2023). National social care data portal for Wales. Available at: https://www.socialcaredata.wales/dataset?c=175&p=171 [Accessed 8th April 2023].

Soley-Bori, M., Ashworth, M., Bisquera, A., Dodhia, H., Lynch, R., Wang, Y., & Fox-Rushby, J. (2021). Impact of multimorbidity on healthcare costs and utilisation: A systematic review of the UK literature. British Journal of General Practice, 71(702). https://doi.org/10.3399/bjgp20X713897

Stafford M, Steventon A, Thorlby R, et al. (2018). Understanding the health care needs of people with multiple health conditions. Health Foundation. Available at: https://www.health.org.uk/publications/understanding-the-health-care-needs-of-people-with-multiple-health-conditions [Accessed 27/04/2023].

Stansfeld S, Clark C, Bebbington P, et al. (2016). Chapter 2: Common mental disorders. NHS Digital Available at: https://files.digital.nhs.uk/pdf/t/6/adult_psychiatric_study_ch2_web.pdf [Accessed 19/04/2023].

Sun H, Saeedi P, Karuranga S, et al. (2022). IDF Diabetes Atlas: global, regional and country-level diabetes prevalence estimates for 2021 and projections for 2045. Diabetes research and clinical practice. 183: 109119. https://doi.org/10.1016/j.diabres.2021.109119

Sung H, Ferlay J, Siegel RL, et al. (2021). Global Cancer Statistics 2020: GLOBOCAN Estimates of Incidence and Mortality Worldwide for 36 Cancers in 185 Countries. CA: A Cancer Journal for Clinicians. 71(3): 209–49. https://doi.org/10.3322/caac.21660

Teoh JYC, Hirai HW, Ho JMW, et al. (2019). Global incidence of prostate cancer in developing and developed countries with changing age structures. PLoS ONE 14(10): e0221775. https://doi.org/10.1371/journal.pone.0221775

The Academy of Medical Sciences. (2018). Multimorbidity: a priority for global health research.

The Health Foundation. (2022). Life expectancy and healthy life expectancy at birth by deprivation [Accessed 08/06/2023].

Tsoi KKF, Hirai HW, Chan FCH, et al. (2017). Predicted increases in incidence of colorectal cancer in developed and developing regions, in association with ageing populations. Clinical Gastroenterology & Hepatology. 15(6): 892–900.e4. https://doi.org//10.1016/j.cgh.2016.09.155

Vaduganathan M, Mensah GA, Turco JV, et al. (2022). The global burden of cardiovascular diseases and risk: a compass for future health. Journal of the American College of Cardiology. 80(25): 2361–71. https://doi.org/10.1016/j.jacc.2022.11.005

WCISU. (2022). Cancer Incidence in Wales, 2002-2019. Available at: https://phw.nhs.wales/services-and-teams/welsh-cancer-intelligence-and-surveillance-unit-wcisu/cancer-incidence-in-wales-2002-2019/ [Accessed 30/03/2023].

Wittenberg R, Hu B, Barraza-Araiza L, et al. (2019). Projections of older people with dementia and costs of dementia care in the United Kingdom, 2019–2040, CPEC Working Paper 5. Care Policy and Evaluation Centre. Available at: https://www.lse.ac.uk/cpec/assets/documents/cpec-working-paper-5.pdf [Accessed 10/03/2023].

Wittenberg R, Hu B, Jagger C, et al. (2020). Projections of care for older people with dementia in England: 2015 to 2040. Age and Ageing. 49(2): 264–9. https://doi.org/10.1093/ageing/afz154

World Health Organization. (2016). Multimorbidity: technical series on safer primary care. Available at: https://www.who.int/publications-detail-redirect/9789241511650 [Accessed 27/04/2023].

World Health Organization. (2021a). Cardiovascular diseases (CVDs). Geneva: WHO. Available at: https://www.who.int/news-room/fact-sheets/detail/cardiovascular-diseases-(cvds) [Accessed 19/04/2023]

World Health Organization. (2021b). Obesity and overweight. Available at: https://www.who.int/news-room/fact-sheets/detail/obesity-and-overweight [Accessed 08/04/2023].

World Health Organization. (2022a). Mental disorders. Available at: https://www.who.int/news-room/fact-sheets/detail/mental-disorders [Accessed 08/04/2023].

World Health Organization. (2022b). Tobacco fact sheet. Available at: https://www.who.int/en/news-room/fact-sheets/detail/tobacco [Accessed 19/04/2023]

World Health Organization. (2022c). World Health Statistics 2022: Monitoring Health for the SDGs. Available at: https://www.who.int/data/gho/publications/world-health-statistics. [Accessed 19/04/2023]

World Obesity Federation. (2023). World Obesity Atlas. Available at: https://s3-eu-west-1.amazonaws.com/wof-files/World_Obesity_Atlas_2023_Report.pdf [Accessed 19/04/2023].

World Stroke Organisation. (2022). World Stroke Organisation (WSO): Global Stroke Fach Sheet 2022 Available at: https://www.world-stroke.org/assets/downloads/WSO_Global_Stroke_Fact_Sheet.pdf [Accessed 16/04/2023].

Yiin GS, Howard DP, Paul NL, et al. (2014). Age-specific incidence, outcome, cost, and projected future burden of atrial fibrillation-related embolic vascular events: a population-based study. Circulation. 130(15): 1236–44. https://doi.org/10.1161/CIRCULATIONAHA.114.010942

